# ECG foundation models share an age-related performance disparity that originates in the data, not the models

**DOI:** 10.64898/2026.10.30.26366943

**Authors:** David Lung, Marie-Kathrin Breyer, Francesco Moscato, Max Haberbusch

## Abstract

**Background:** Electrocardiogram (ECG) foundation models are proposed for clinical use and promoted as fair, yet such claims are rarely tested with subgroup-stratified evaluation. We measured demographic disparities across four ECG foundation models, tested whether mitigation removes them, and asked whether the dominant disparity originates in the models or in the data and the clinical task.

**Methods:** We probed frozen embeddings of four architecturally distinct ECG foundation models (ECGFounder, xECG, ECG-FM, DeepECG-SSL) in thirteen cohorts on four continents (5.4 million analysed ECGs, 31 endpoints, six held-out cohorts), stratified by age, sex and race. We examined embedding geometry, tested post-hoc correction, larger labelled training sets, comorbidity exclusion and low-rank fine-tuning, and compared each network with an untrained copy of itself and with logistic regression on classical ECG measurements.

**Findings:** Disparities concentrated on age, not sex or race (median worst-group AUROC gap 0.028 *vs* 0.004 and 0.016; age exceeded sex in 97% of results) and replicated externally (ED-BWH 30-day MACE AUROC 0.81–0.90 under 40 years, 0.64–0.71 at ≥75). Age formed a smooth gradient in the embeddings, whereas sex, though most decodable, carried no gap. No remedy closed it (low-rank adaptation: −0.007 to −0.004). Untrained copies (r=0.81, 102 pairs) and classical measurements (*ρ*=0.90) reproduced the endpoint-specific gaps, and pretraining raised AUROC (+0.067) without changing them. The largest race effect, higher potassium error in Black patients, vanished at matched true potassium.

**Interpretation:** ECG foundation models carry an age-structured disparity that replicates externally and survives mitigation. Because untrained networks and a classical model reproduce it, the results indicate that it is inherited from the age structure of the ECG and its labels rather than created by the models, so switching or adapting models will not remove it. Age-stratified reporting, untrained and classical baselines, and age-specific thresholds and recalibration are therefore needed.

**Funding:** Austrian Research Promotion Agency (FFG; CARDISENSE project, grant FO999922422).

**Graphical abstract:** 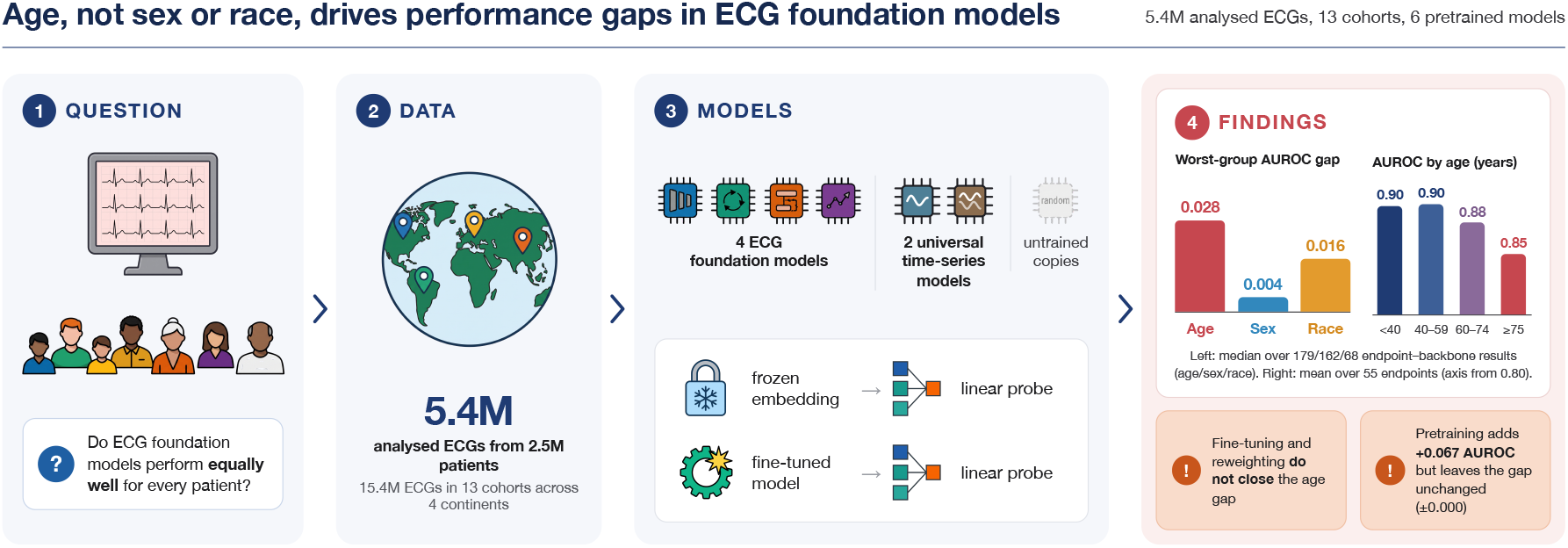

**Research in context:** *Evidence before this study:* We searched PubMed, Google Scholar, and arXiv from database inception to October, 2026, using combinations of the terms “ECG” or “electrocardiogram”, “foundation model” or “self-supervised”, and “fairness”, “bias”, “equity”, “subgroup”, or “demographic”, with no language restriction. Open-weight ECG foundation models have proliferated and are increasingly proposed for clinical use, and benchmarks compare them on aggregate accuracy without stratifying by subgroup. The fairness literature documents demographic performance gaps in task-specific ECG classifiers, most often for sex or race on single tasks in single cohorts, and the remedies it proposes act on the model (reweighting, adversarial debiasing, group-robust training, fine-tuning). The one subgroup analysis of ECG foundation models we found covered two models of one family on their own endpoints, stratified by age and sex only. To our knowledge, no study has evaluated several architecturally distinct ECG foundation models along age, sex and race across endpoint families and continents, and none has tested whether a disparity found in a foundation model is also present without any pretraining or without deep learning.

*Added value of this study:* Evaluating four architecturally distinct ECG foundation models across thirteen cohorts on four continents, including six held-out external cohorts, we show that strong average and external discrimination coexists with a disparity that concentrates on age, not sex or race, and that replicates externally. The pattern has a consistent substrate: age is encoded as a smooth gradient in the embeddings, while sex and race, both decodable, carry no comparable gap. The gap is shared across every architecture and pretraining objective, and neither post-hoc correction, larger labelled training sets, exclusion of comorbid ECGs nor low-rank fine-tuning closes it. Two controls explain why: untrained copies of the same networks and a logistic regression on classical ECG measurements reproduce the endpoint-specific age gaps, which indicates that the gap is inherited from the data and the clinical task rather than learned in pretraining.

*Implications of all the available evidence:* Fairness evaluation of ECG foundation models should prioritise age and report age-stratified performance as a primary result, because a threshold fixed on the pooled population operates at a worse point for older patients than the headline metric implies. Because the gap persists across architectures, vendors, fine-tuning and even untrained networks, switching models will not resolve it. Evaluations should include an untrained and a classical baseline before attributing a gap to a model, and services deploying these models should use age-specific thresholds and recalibration, or at minimum report and monitor performance by age band.

## 1 Introduction

Deep learning applied to the 12-lead electrocardiogram has progressed from arrhythmia classification [1] to the inference of conditions with no classical ECG signature, such as left-ventricular dysfunction [2], hyperkalaemia and latent atrial fibrillation [3], and recently to risk markers for sudden cardiac death. ECG foundation models [4–7] now generalise across many such tasks and are increasingly proposed for clinical deployment, with their scale and demographic breadth presented as answering the fairness concerns raised against earlier task-specific models [4], and with fairness analyses restricted to age and sex at a single operating point [7]. Such claims are rarely supported by discrimination evaluated separately by age, sex and race, so whether these models are equitable where they would be used remains largely uncharacterised. This matters because fairness failures in clinical machine learning are well documented, from a care-management algorithm that under-served Black patients [8] to under-diagnosis of under-served groups by imaging models [9] and the recovery of self-reported race from images clinicians cannot read [10].

Foundation models reach clinical tasks most often as *frozen feature extractors*, where a fixed pretrained backbone produces an embedding that a lightweight probe maps to a clinical label, and increasingly through parameter-efficient fine-tuning of the backbone. Benchmarks such as OpenECG [11] and BenchECG [6] compare aggregate cross-centre generalisation without stratifying by subgroup, and the one fairness analysis of an ECG foundation model family [7] is confined to two models on their own endpoints, micro-averaged over 77 classes and stratified by age and sex only. Prior work has shown that individual, task-trained ECG and cardiac-imaging models carry race, sex and age disparities [9, 12], and that model-side remedies often fail to generalise [13].

We evaluate four architecturally distinct ECG foundation models (ECGFounder [4], convolutional; xECG [6], recurrent (xLSTM); ECG-FM [5], supervised Transformer; DeepECG-SSL [7], self-supervised Transformer) under one frozen-probe protocol across thirteen cohorts on four continents, and ask whether low-rank fine-tuning removes any disparity the frozen-probe evaluation reveals. Three controls under the identical pipeline go one step further: universal time-series foundation models that have never seen an ECG, an untrained copy of every network, and a logistic regression on classical ECG measurements. A disparity that appears without ECG pretraining, without any training and without deep learning cannot be a property of the models and must come from the data and the clinical task. We therefore measure subgroup performance, including on held-out external cohorts, trace the dominant disparity to its substrate in the embedding geometry, test whether mitigation and fine-tuning remove it, and test whether it is present without pretraining or deep learning.

## 2 Methods

### Cohorts and endpoints

Thirteen de-identified cohorts span four continents (Fig. 1 and Supplementary Table S1): HEEDB-MGH and HEEDB-Emory [14], MIMIC-IV-ECG [15], EchoNext [16] and ED-BWH (United States), CODE [17] and CODE-II (Brazil), PTB-XL [18] (Germany), LEAD [19] (Austria), IKEM (Czechia), and Shaoxing, CPSC2018 and SPHDB (China). The analysis sets comprise 5,396,975 ECGs from 2,509,462 patients (curation flows in Supplementary Figs S1–S3). The 31 endpoints (Supplementary Table S2) cover rhythm and conduction, structural and ischaemic disease, laboratory values within *±*6 h of the ECG, outcomes and chronological age, and each classification endpoint is grouped by how its label is obtained (outcome, coded diagnosis, diagnosis read from the ECG, measurement). Six cohorts (CODE-II, EchoNext, IKEM, ED-BWH, SPHDB, LEAD) are in no model’s pretraining corpus (Supplementary Table S3), and no model had seen any endpoint label. ED-BWH is analysed on its three 10-s leads (12-lead sensitivity analysis in the Supplementary Methods). Cohorts are analysed separately and never pooled.

**Figure 1:**
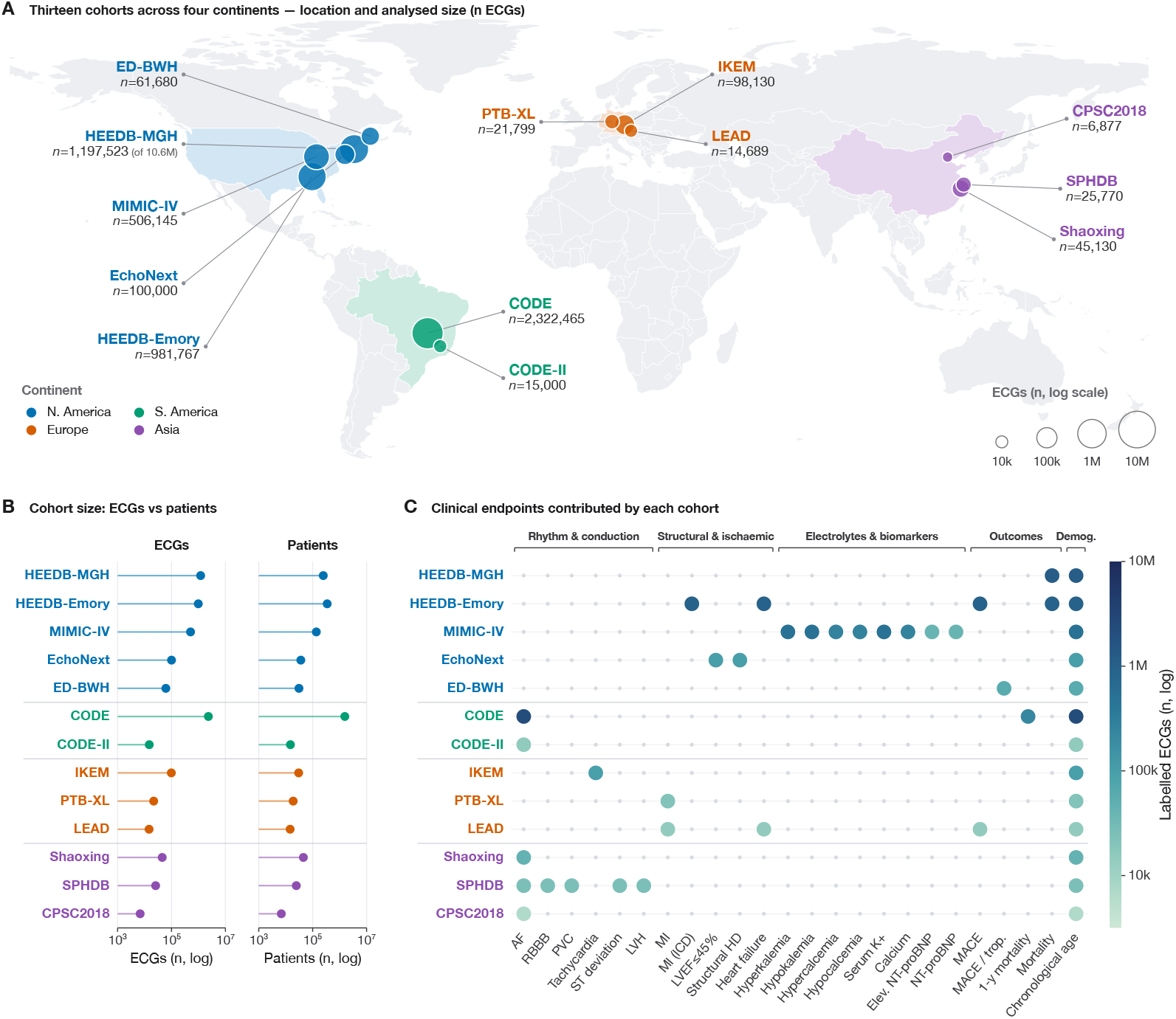
Thirteen cohorts, four continents and 31 endpoints. **(A)** Location and size of the thirteen cohorts; marker area is proportional to the number of analysed ECGs (log scale) and colour shows the continent. HEEDB-MGH is shown with its patient-grouped probe subsample (1,197,523 of 10,608,417 ECGs), CODE with all 2,322,465 ECGs and LEAD with 14,689 ECGs. **(B)** Number of ECGs and of unique patients per cohort (log scale). **(C)** Endpoints contributed by each cohort, coloured by the number of labelled ECGs; chronological age is available in every cohort. ED-BWH is shown with its analysis set (ECGs eligible for the MACE endpoint).

### Models and read-out

We used ECGFounder [4], xECG [6], ECG-FM [5] and DeepECG-SSL [7] with their public weights and native preprocessing (Supplementary Table S4), spanning supervised and self-supervised pretraining, and two universal time-series models never trained on ECGs: TiRex-2 [20], built like xECG on the xLSTM architecture, and MOMENT-1-large [21], a Transformer encoder that processes each lead as a separate series. Throughout, *pretrained* denotes a network with its publicly released weights, which we did not modify except in the low-rank adaptation experiments; *untrained* denotes the same architecture with random initialisation. For each backbone, cohort and endpoint we fitted an ℓ_2_-regularised logistic or ridge probe with patient-grouped five-fold out-of-fold prediction, so that every labelled ECG is scored held-out. We report AUROC (classification) and MAE (regression) overall and by age band (*<*40, 40–59, 60–74, ≥75 years), sex and race, with 1,000-fold bootstrap 95% CIs. Race is the category recorded in the source electronic health record (White, Black, Asian; US cohorts only). The *worst-group gap* is the best minus the worst subgroup on one axis. The *age gap* is AUROC in the youngest minus the oldest band (for MAE, the reverse), so that a positive value means worse performance in older patients. A secondary definition, AUROC at 40–59 minus ≥75 years, serves endpoints whose youngest band is sparse.

### Embedding geometry and attention

Embeddings were described by UMAP, by a silhouette index of how separated the groups of each attribute are in the embedding space (within-group against nearest-other-group distance; 0 means no structure), and by the decodability of each attribute, the gain in balanced accuracy over chance of a linear probe predicting age band, sex or race from the embedding. A detached cluster was characterised by HDBSCAN and acquisition metadata. To ask where the models attend and whether any wave carries the gap, SmoothGrad saliency was computed on delineated beats for the four ECG backbones on eight endpoints, and the P wave, QRS complex or T wave was set to zero and the probe refitted (Supplementary Methods).

### Mitigation strategies

Because AF, paced rhythm and bundle-branch block accumulate with age and could carry the gap, age gaps were re-scored after removing ECGs with these findings (never the target condition). Because a gap could reflect too few labels, probes were trained on 100 to 30 000 labelled ECGs for seven endpoints. Because it could reflect the capacity or objective of the read-out, on held-out patient-level splits we compared the standard probe with per-age-band read-outs, error-focused (focal) reweighting and a per-band gradient-boosted read-out (10 endpoints, four backbones). Because it could reside in the frozen weights, we compared frozen probes with low-rank adaptation (LoRA [22], rank 8) of each ECG backbone under empirical-risk, group-robust [23] and class-balanced objectives (114 age-stratified cells), each with a paired frozen probe on the identical split.

### Controls without learned weights or deep learning

To test whether the gap requires learned weights at all, each architecture was rebuilt with random initialisation and no loaded weights, and probed exactly as the pretrained model (117 endpoint–backbone pairs, 102 with AUROC endpoints). MOMENT-1-large, its untrained copy and two further random initialisations of the five networks were run on patient-grouped subsets of at most 100,000 ECGs per cohort, with paired probes on the same ECGs (Supplementary Methods). To test whether it requires deep learning, logistic regression and gradient-boosted trees were trained under the same folds on 20–48 standard measurements (intervals, durations, axes, amplitudes) and their age gaps compared with the median pretrained gap. To locate the gap, we computed the AUROC of cases from each age band against controls from each band and, for four endpoints, miss rate, specificity and calibration by age band at a single threshold versus age-specific thresholds chosen on separate patients. For the potassium race effect we compared errors within 0.25 mmol/L bins of true potassium, standardised the Black error to the White potassium distribution and adjusted for eGFR (Supplementary Methods).

### Statistics

Inference rests on effect sizes with bootstrap 95% CIs (1,000 resamples of ECGs, percentile method) and their consistency across backbones and cohorts. Hypothesis tests are limited to pre-defined paired comparisons: Wilcoxon signed-rank tests across endpoints for changes under each mitigation strategy, a Mann–Whitney test for the label-type contrast, and Pearson or Spearman correlations for the concordance of gaps between pretrained, untrained and classical models. All tests are two-sided and Holm-adjusted within each family (Supplementary Table S5). By a pre-specified rule, age-band results enter the summary statistics only where the youngest and the oldest band each contain at least 20 positive ECGs. The 10 endpoints that fail it (Supplementary Table S6) are shown but excluded from every summary statistic. LEAD MACE and MI, whose youngest band is sparse, are analysed on the secondary definition (Supplementary Methods). Lowering the threshold to 10 positives or removing the rule changed no conclusion (Supplementary Table S7). The study is reported following TRIPOD+AI [24] (checklist in Supplementary Table S12); it was not registered and no protocol was published, and patients and the public were not involved. Full details are in the Supplementary Methods.

## 3 Results

### 3.1 Strong discrimination, variable external generalisation, and a disparity that concentrates on age

Across thirteen cohorts on four continents (5.4 million analysed ECGs from 2.51 million patients; Fig. 1), the four backbones reach clinically useful discrimination and agree closely with one another (median range of AUROC across backbones per endpoint 0.015, interquartile range 0.006–0.035; Fig. 2A; Supplementary Tables S8 and S11): frozen-probe AUROC is high on structural endpoints (EchoNext LVEF ≤45% 0.824–0.879) and MI (PTB-XL 0.878–0.922), near-ceiling on atrial fibrillation (CODE 0.971–0.981, 2,322,465 ECGs), and moderate on physiological inference (hyperkalaemia 0.653–0.753). This competence carries onto the held-out cohorts, but not uniformly (Fig. 2B): MACE reaches AUROC 0.75–0.77 on Emory and 0.75–0.83 on ED-BWH, yet on the European LEAD cohort ECGFounder degrades sharply (heart failure 0.61 versus 0.81–0.83 for the other three). LEAD, whose youngest band holds too few events for the primary age gap, was analysed on the secondary 40–59 versus ≥75 definition (Supplementary Fig. S4). On it, LEAD MACE and MI showed no detectable decline (gaps −0.042 to 0.108 and −0.011 to 0.080, no CI excluding zero), whereas Emory MI, Emory heart failure and ED-BWH MACE lost 0.102–0.128 AUROC on the same definition. LEAD sex gaps were 0.02–0.14.

**Figure 2:**
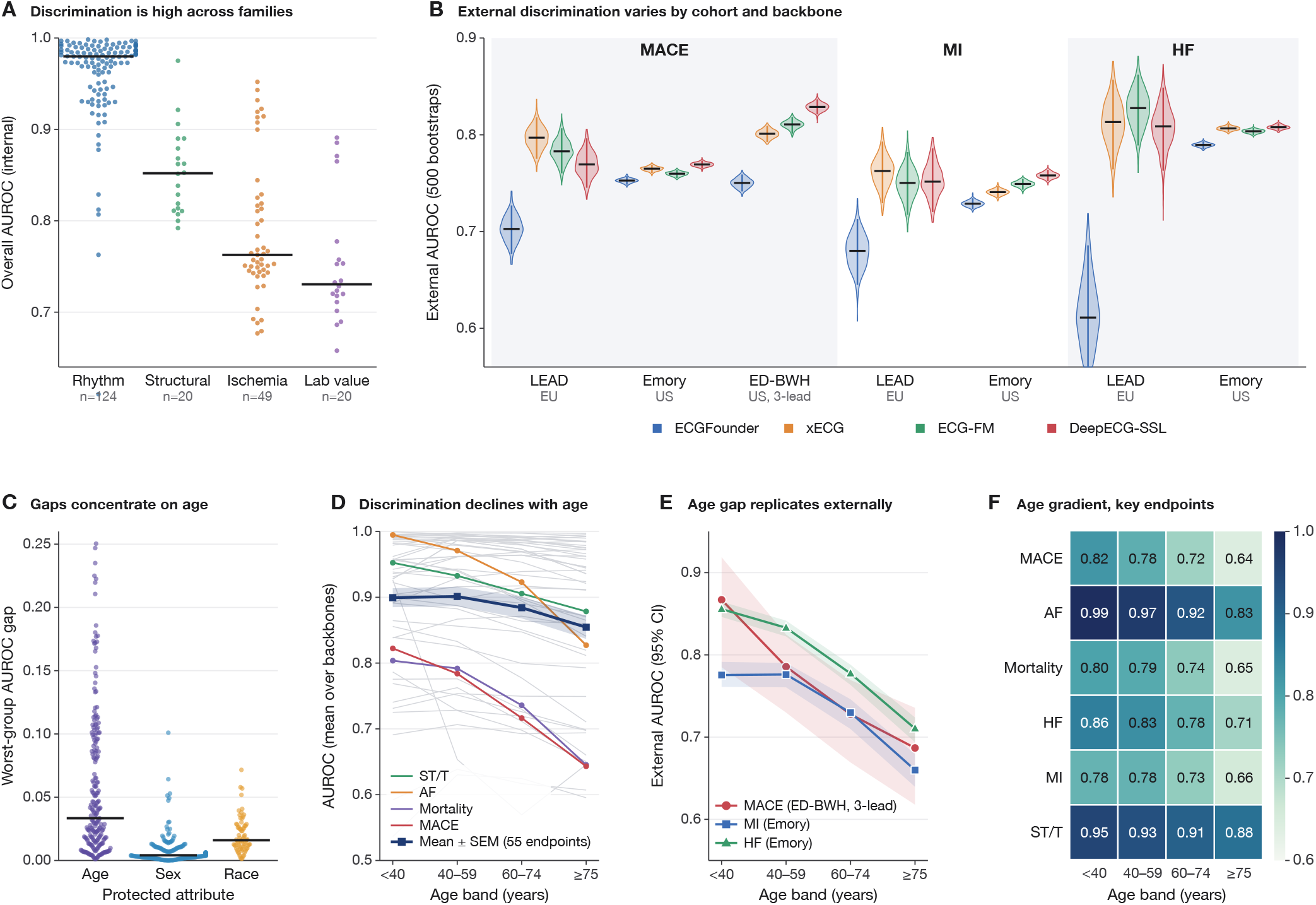
Capability and its age-structured disparity. **(A)** Frozen-probe discrimination by endpoint family for the four backbones; one point per endpoint–backbone result, bar = median. **(B)** External discrimination varies by cohort and backbone: overall AUROC for MACE, MI and heart failure on the held-out cohorts (LEAD, Emory, ED-BWH 3-lead input), bootstrap violins. **(C)** Worst-group AUROC gap by protected axis, each point an endpoint *×* backbone cell, bar = median. **(D)** AUROC by age band across endpoints (grey, all endpoints; mean*±* SEM over backbones; named endpoints highlighted). **(E)** The age gap replicates on the held-out cohorts (Emory MI and heart failure, ED-BWH MACE), mean over the four ECG backbones with bootstrap 95% CI ribbons. **(F)** Age gradient across key endpoints. Cohorts are analysed separately and never pooled; ED-BWH is shown with its primary 3-lead input (12-lead sensitivity analysis in Supplementary Table S8).

Beneath this competence a consistent disparity emerges, and it concentrates on **age** (Fig. 2C–F). The median worst-group gap was 0.028 for age, 0.004 for sex and 0.016 for race, and the age gap exceeded the sex gap in 97% of 309 results and the race gap in 57% of 177. Discrimination declined toward the oldest band on 12 of 16 classification endpoints, was near-identical between the sexes, and varied little by race, with the worst-served race group changing from endpoint to endpoint (White in 31, Asian in 24, Black in 16 of 102 results). Age gaps of different backbones were closely related across endpoints (pairwise Spearman *ρ* 0.85–0.95). The age gap *replicates on the held-out external sets* (Fig. 2E): Emory MI fell from AUROC 0.77–0.79 under 40 years to 0.64–0.68 at ≥75, Emory heart failure from 0.85–0.86 to 0.70–0.72, and ED-BWH 30-day MACE from 0.81–0.90 to 0.64–0.71, with bootstrap 95% CIs no wider than 0.04 for Emory (104,149–365,018 ECGs per band) and 0.13 for ED-BWH (3,899–24,576 ECGs per band).

The largest single race effect, higher serum-potassium error in Black than White patients (0.51– 0.55 versus 0.43–0.46 mmol/L, all five backbones), followed the label distribution: true potassium varies more in Black patients (SD 0.85 versus 0.69 mmol/L) and ridge probes shrink toward the mean, so that at matched true potassium the gap fell from +0.074–0.084 to 0.001–0.006 mmol/L (92–99% explained), unchanged by adjustment for kidney function, down-sampling White training data or Black-only training (Supplementary Fig. S5). It is a property of the label distribution under a shrinking read-out, the kind of hidden stratification that aggregate metrics miss [25].

### 3.2 Age is written into the embedding geometry, and decodability alone does not imply a disparity

Two geometric properties must be distinguished (Fig. 3). In UMAP projections age varies smoothly across the manifold while sex and race are intermixed (Fig. 3A–C), and the silhouette index confirms that age is by far the most geometrically structured attribute (0.28 versus 0.10 for sex and 0.11 for race). Yet a linear probe decodes **sex** at least as well as age (balanced-accuracy gain 0.19–0.25 versus 0.16–0.22, and 0.03–0.07 for race) although neither sex nor race carries a comparable performance gap (Fig. 3E,f). Decodability therefore does not imply disparity: age is singled out by the conjunction of a smooth gradient *and* declining discrimination along it (Fig. 3D). The detached cluster in the projection is an acquisition-device group and does not explain the gap (Supplementary Fig. S6). In saliency maps, older beats draw relatively more P-wave attention than young beats in two backbones (ECGFounder and xECG, not ECG-FM or DeepECG-SSL; Supplementary Fig. S7), tracking the atrial morphology that changes most with age: P-wave duration and PR interval lengthen steadily across adult life [26] and dominate ECG-based age estimation [27]. The shift is a correlate of the gap, not its cause. In wave-blanking experiments (delineated P wave, QRS complex or T wave set to zero before probing), blanking the P wave lowered AUROC by only 0.0031 and changed the age gap by −0.0001, whereas blanking the QRS cost 0.0307 AUROC in every age band without changing the gap either (Supplementary Fig. S8). The models discriminate from the QRS, and no single wave carries the gap.

**Figure 3:**
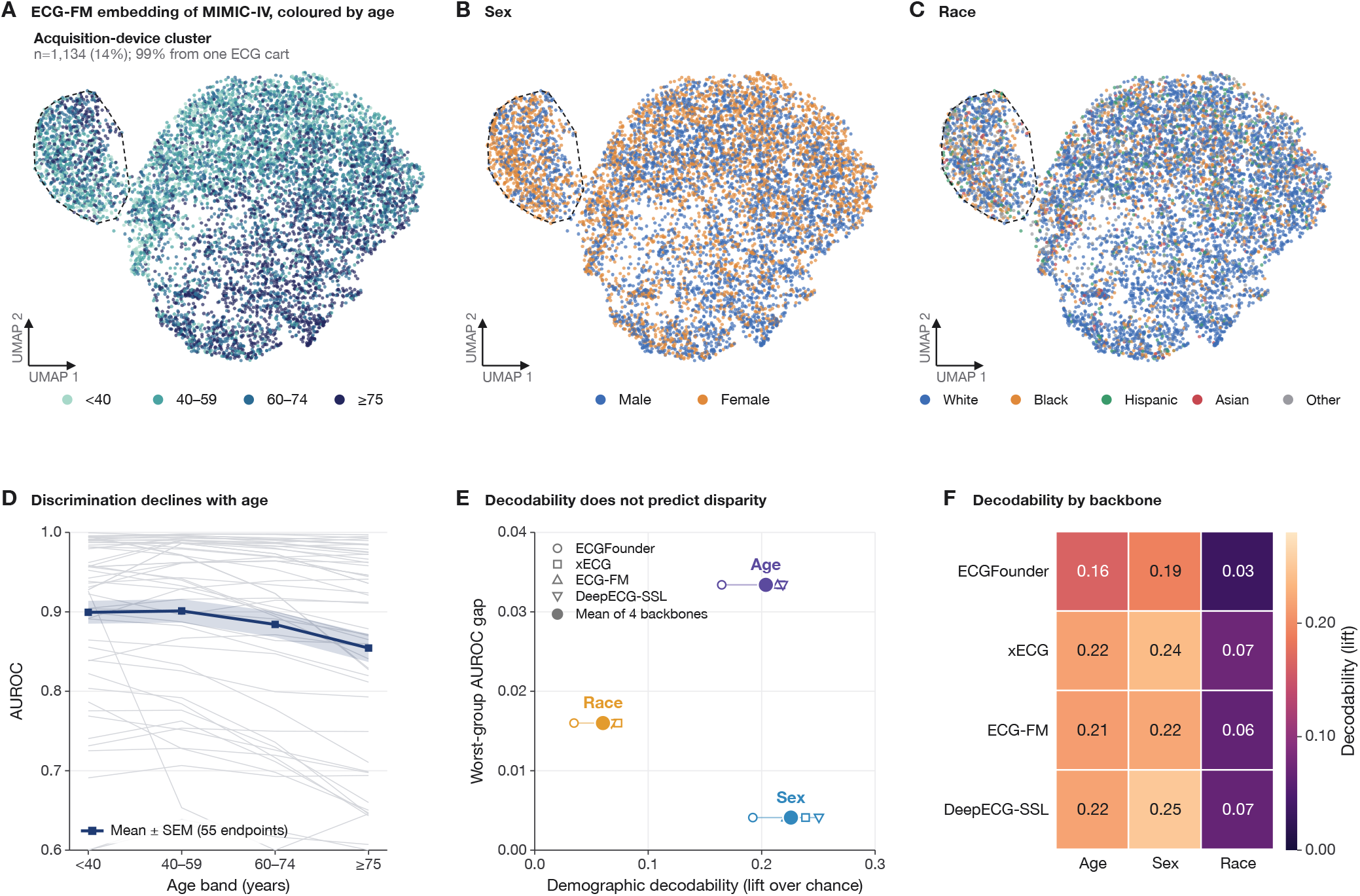
The embeddings encode age as a geometric gradient, and decodability does not track disparity. **(A–C)** UMAP projection of ECG-FM embeddings of 8,000 MIMIC-IV-ECG test ECGs coloured by age band, sex and race; the detached cluster (1,134 ECGs) is an acquisition-device group (Supplementary Fig. S6). **(D)** AUROC by age band for all classification endpoints (grey) and mean *±* SEM. **(E)** Linear decodability of each attribute (balanced-accuracy gain over chance, EchoNext) against its worst-group gap; small markers, backbones; large, mean. **(F)** Decodability by backbone and attribute.

### 3.3 Neither changing the data nor changing the model closes the age gap

If the gap were an artifact of the frozen read-out, of too few labels, or of the comorbid ECGs that accumulate with age, changing those should remove it. None does (Fig. 4). AF, pacing and bundle-branch block are far more common in older patients (AF in MIMIC-IV 1.7% under 40 versus 23.0% at ≥75), yet removing such ECGs changed the gap by at most 0.028 AUROC in 149 combinations (median −0.003; Fig. 4A), and measured comorbidity and severity explained 28–51% of the age trend in error, never all of it (Supplementary Fig. S21). Older patients are not scarce (Emory: 205,701 ECGs at ≥75), and increasing the labelled set from 3,000 ECGs to the full set raised AUROC by 0.016 (p=0.0001) but changed the gap by only − 0.010 (Holm-adjusted p=0.54; Fig. 4B). Model-side changes did no better. Group-robust optimisation [23], class balancing, per-age-band probes and focal reweighting changed the gap by medians of +0.000, +0.000, +0.003 and +0.002 (all adjusted p=1.0) while costing overall AUROC (−0.005 to −0.171; Fig. 4C), and a non-linear per-band read-out never recovered the oldest band (avoidable gain 0.025 to 0.001; Fig. 4D). Low-rank fine-tuning of the four backbones raised overall AUROC (+0.011 to +0.021) and AUROC at ≥75 in 97–100% of 114 cells, but changed the gap by only −0.007 to −0.004, because it raised accuracy in young patients too (Supplementary Fig. S9). Letting the backbone move its weights does not reallocate accuracy across age. The age-dominant, sex-null fingerprint survives every intervention and every architecture, so the disparity is not a tunable property of read-out capacity, label quantity or comorbid case mix. What can be changed is the operating point. With one threshold set for 90% overall sensitivity, 19.5% of Emory MACE cases were missed under 40 years against 2.9% at ≥75, while specificity fell from 0.71 to 0.08. Age-specific thresholds chosen on separate patients restored 90% sensitivity in every band and raised specificity in the oldest (Fig. 4E), and per-band recalibration brought the expected calibration error, which rose with age under pooled calibration (0.220 versus 0.317, Emory MI), to ≤0.024 in every band (Fig. 4F; Supplementary Figs S19 and S20, Table S9).

**Figure 4:**
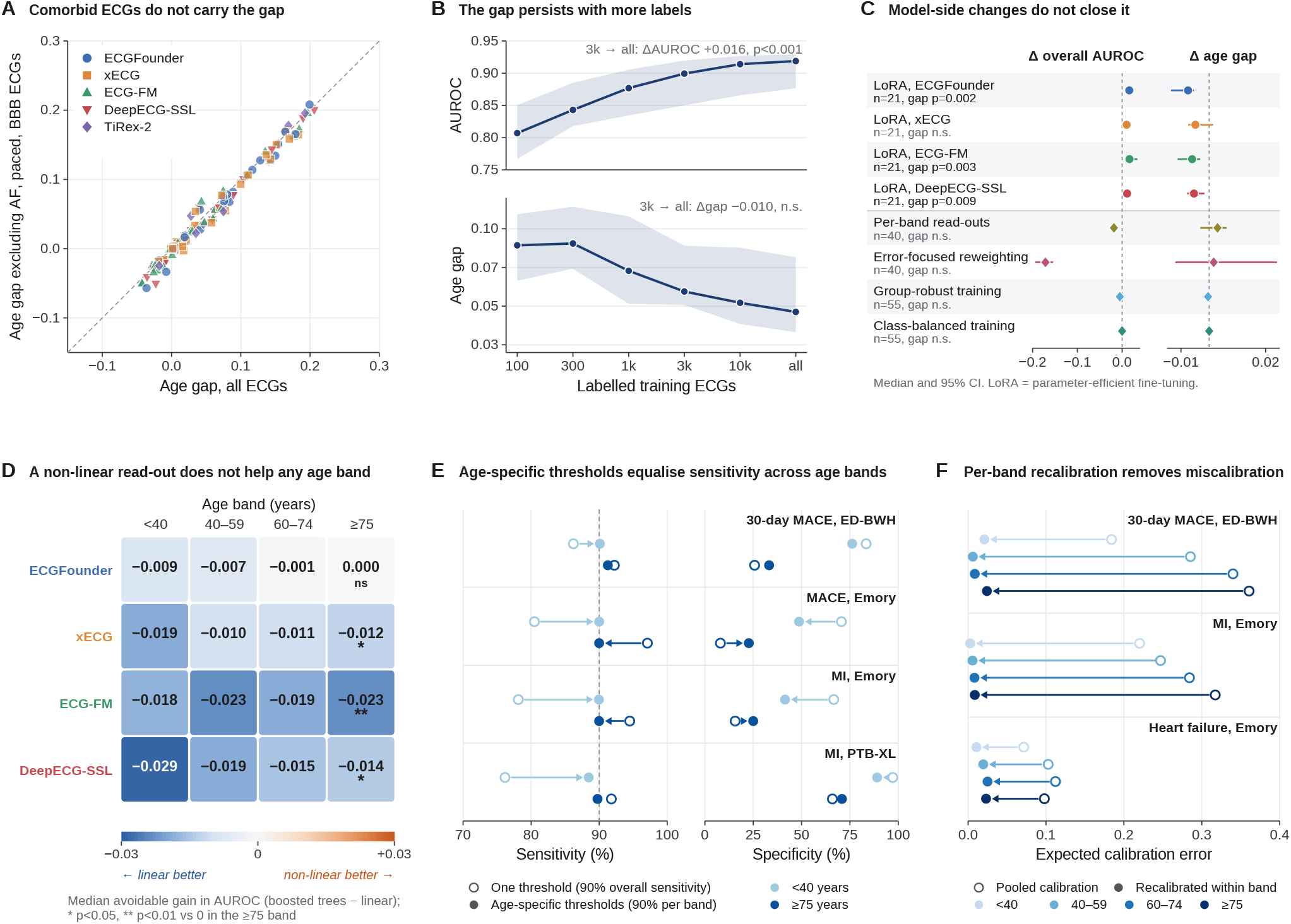
Changing the data or the model does not close the age gap; thresholds and calibration manage it. **(A)** Age gap before and after removing ECGs with AF, paced rhythm or bundle-branch block (never the target), 149 endpoint–backbone combinations; dashed, identity; inset, median change with 95% bootstrap CI. **(B)** Overall AUROC (top) and age gap (bottom) against the number of labelled training ECGs: median and interquartile range over 14 learning curves (seven endpoints, ECGFounder and xECG, three seeds); insets, change from 3,000 ECGs to the full set with Wilcoxon p (Supplementary Fig. S10). **(C)** Change in overall AUROC and in age gap after low-rank adaptation of the four ECG backbones (dots, 114 results; circles, median per backbone) and median changes for per-band read-outs, error-focused (focal) reweighting, group-robust and class-balanced objectives (diamonds); bars, 95% CIs; legend, raw Wilcoxon p for the change in gap (Holm-adjusted values in Supplementary Table S5). **(D)** Median avoidable gain of a per-band gradient-boosted read-out over the linear probe, by backbone and age band (10 endpoints per backbone); asterisks, Wilcoxon p*<*0.05 (*) and p*<*0.01 (**) against zero in the ≥75 band. **(E)** Sensitivity and specificity in the youngest and oldest band with one threshold set for 90% overall sensitivity (open) and with age-specific thresholds set for 90% sensitivity per band (filled, arrow), chosen on separate patients (two-fold patient cross-fitting); ensemble of the four ECG backbones, four externally validated endpoints. **(F)** Expected calibration error by age band with pooled calibration (open) and after recalibration within band (filled). Endpoints failing the small-*n* rule are excluded.

### 3.4 The gap is present without pretraining and without deep learning

A gap that every architecture shares and no adaptation removes could still be blamed on something all pretrained networks have in common. Two controls rule this out (Fig. 5). Untrained copies of the six networks, with random weights never exposed to an ECG, retain much of the discrimination (median AUROC 0.729 versus 0.793) and reproduce the endpoint-specific age gaps: across 102 classification pairs, untrained and pretrained gaps correlated at r=0.81, with near-identical medians (0.055 versus 0.058). Endpoints with a large gap kept it without any training (ED-BWH MACE 0.184 versus 0.168) and EchoNext LVEF had none either way (− 0.016 versus −0.021). Pretraining raised AUROC in 102 of 102 pairs (median +0.067) but changed the gap by 0.000 (Fig. 5D). Pretraining lifts the curve but does not tilt it. A logistic regression on classical ECG measurements, with no deep learning at all, reproduced the same gaps (*ρ*=0.90 over 16 endpoints; median 0.054 versus 0.053; Fig. 5C), as did boosted trees (*ρ*=0.91), also with equal numbers of ECGs per age band (Supplementary Figs S11–S15). Neither result depends on the random draw or on the xLSTM architecture TiRex-2 shares with xECG: across three random initialisations the across-seed SD of the untrained gap was 12–14% of its spread across endpoints, and MOMENT-1-large reproduced the endpoint-specific gaps with its released weights (r=0.94 over 17 endpoints) and less closely as an untrained copy (median AUROC 0.663; Supplementary Fig. S16).

**Figure 5:**
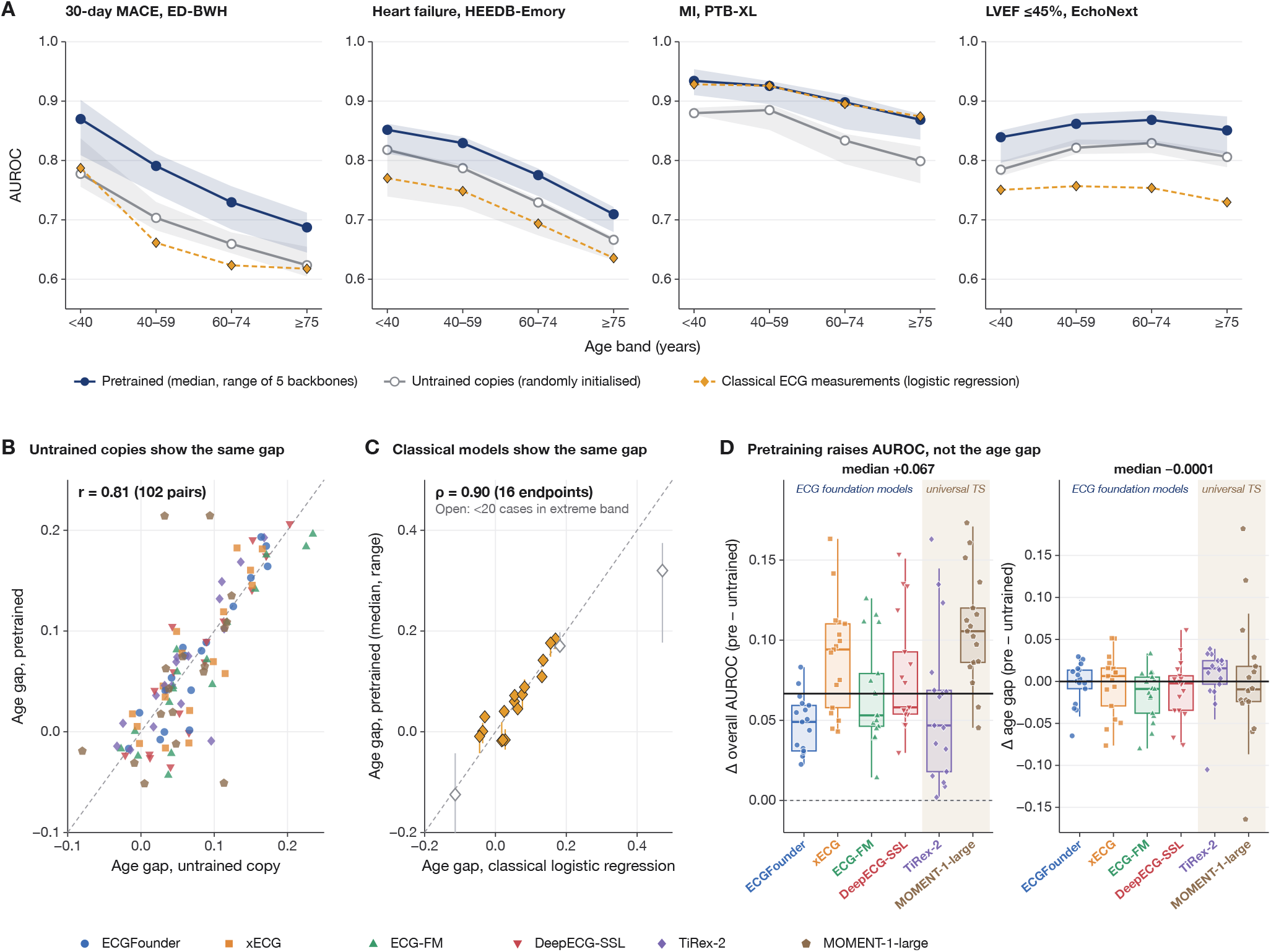
Untrained networks and classical measurements reproduce the age gap. **(A)** AUROC by age band for three endpoints with an age gap and one without: pretrained models (line, median; band, range over five backbones), untrained copies (grey) and logistic regression on classical ECG measurements (diamonds); ED-BWH, 3-lead input. **(B)** Age gap of each pretrained backbone against its untrained copy, 102 classification pairs (Pearson r=0.81; MOMENT-1-large pairs from the capped subsets; regression endpoints in Supplementary Fig. S12). **(C)** Age gap of the classical model against the median pretrained gap (bars, range over backbones), 16 endpoints (Spearman *ρ*=0.90); open markers, endpoints failing the small-*n* rule. **(D)** Change from untrained copy to pretrained backbone in overall AUROC and in age gap (102 pairs): points, pairs; boxes, median and interquartile range per backbone (shaded, universal time-series models); black line, overall median. Dashed diagonals, identity.

The gap is concentrated where labels depend on more than the ECG and where older non-cases resemble cases (Supplementary Fig. S17): the median age gap was 0.142 for outcome and coded-diagnosis endpoints and 0.030 for ECG-read or measured labels (p=0.0005), with the same contrast for untrained copies and classical models, and discrimination depended more on the age of the controls than of the cases (PTB-XL MI: cases ≥75 against controls *<*40, AUROC 0.99; against controls of their own age, 0.90; young cases against old controls, 0.78; Supplementary Fig. S18). Together with the invariance across architectures, objectives and adaptations, these controls indicate that the age gap lies in the age structure of the ECG and of its clinical labels rather than in the models.

## 4 Discussion

Across four ECG foundation models and thirteen cohorts, strong average and external discrimination coexists with a systematic disparity that concentrates on **age**, not sex or race. It replicates externally, has a representational substrate and resists every standard remedy. The same fingerprint is shared by every architecture and pretraining objective, reproduced by two time-series models that have never seen an ECG, by untrained copies of every network and by a logistic regression on classical measurements, and unchanged by larger labelled sets, comorbidity exclusion, alternative read-outs, group-robust objectives or low-rank fine-tuning. The results therefore indicate that the models *inherit* this disparity from the age structure of the ECG and of its clinical labels rather than create it. Three observations say where: the gap is largest for outcome and coded labels, which depend on information the ECG is unlikely to contain; discrimination depends more on the age of the controls than of the cases, because older people without the condition carry ECGs that resemble disease; and the comorbid ECGs we can flag carry only a small part of it. The second observation is physiology, not artefact: the ECG’s age signal is dominated by atrial features that strengthen with hypertension and smoking, and an ECG age ≥8 years above chronological age doubles mortality risk [27] — older-looking ECGs also look sicker. The gap is nonetheless real for patients and must be managed at deployment.

### Clinical implications

For a health system evaluating an ECG foundation model, the disparity to plan for is age, and it is invisible in the headline number. At a fixed threshold the *type* of error changes with age: a pooled threshold under-detects disease in the young and generates false alarms in the old, and calibration worsens with age. Both are manageable with age-specific thresholds and per-band recalibration (Fig. 4E, F). Three recommendations follow. Model cards [28] and prediction-model reporting [24] should present age-stratified discrimination, miss rate and calibration as primary results. Because the gap persists across every backbone, an untrained network and a classical model, switching vendors will not resolve it, and any fairness evaluation of a foundation model should include an untrained copy and a classical baseline before attributing a gap to the model. Deployments should use age-specific thresholds and recalibration, or at minimum report and monitor by age band.

### Toward mitigation

Because the gap is present before any ECG pretraining and unchanged by larger labelled sets, demographically balanced pretraining and more labels are unlikely to close it on their own. Remedies that act on the task are more promising, such as labels that depend less on non-ECG information (adjudicated rather than coded outcomes), decision thresholds defined within age strata, and explicit modelling of the age-dependent control distribution. Any model-side gain should be measured against the untrained and classical baselines reported here, on held-out data, with an explicit accuracy–fairness trade-off.

### Limitations

The non-ECG references are two models from different architecture families, run (with the additional seeds) on subsets of at most 100,000 ECGs per cohort, and the untrained MOMENT copy reproduced the gaps less closely than the untrained ECG networks; the concordance that carries the argument is between models that share nothing. Adaptation was evaluated as rank-8 LoRA on capped training sets rather than full fine-tuning, which is how frozen foundation models are adapted in practice, and the untrained and classical controls bound what any adaptation could reallocate. Race labels exist only for the three US cohorts and are EHR-recorded categories, so the race results describe those health systems, where the gaps were small and unstable in direction. LEAD has too few events under 40 years for the primary age-gap definition and enters the age analysis through the secondary 40–59 versus ≥75 gradient, on which it showed no decline with wide CIs and an oldest band limited to 75–80 years by its recruitment, while contributing fully to the overall external comparison. Bootstrap CIs resample ECGs while folds are patient-grouped and are therefore somewhat narrow in cohorts with repeated ECGs, but no conclusion rests on a single interval. The continental comparison is confounded with device and protocol [29], which is why every gap is estimated within a cohort; potassium is US-only; and ED-BWH’s print-format waveforms required a 3-lead primary analysis, with which the 12-lead sensitivity analysis agrees. Finally, the controls show that the gap is of similar size in untrained, classical and pretrained models, not that its avoidable component is zero.

## Data Availability

Primary ECG data are available from their source repositories under the respective licences (Supplementary Table S10). Figure source data and supporting tables are deposited on Zenodo (doi:10.5281/zenodo.23814933). Analysis code will be released on publication at https://github.com/CellularSyntax/ecg-fm-age-disparity (MIT licence).

https://doi.org/10.5281/zenodo.23814933

## Data sharing

### Data

All primary data derive from previously published, publicly released or credentialed-access cohorts, each obtained under its licence or data-use agreement (sources, references and licences in Supplementary Table S10): MIMIC-IV-ECG [15], PTB-XL [18], the Chapman–Shaoxing database and CPSC2018 (PhysioNet [30]), CODE-15% [17] (Zenodo) and the full CODE and CODE-II cohorts (Telehealth Network of Minas Gerais, on request), IKEM (Zenodo, CC BY 4.0), SPHDB (figshare), HEEDB-MGH and HEEDB-Emory [14] (Brain Data Science Platform, credentialed access), EchoNext [16] (PhysioNet, restricted access), ED-BWH (Nightingale Open Science), and LEAD [19] (data-use agreement with the LEAD study group). Raw waveforms cannot be redistributed by the authors. The derived analysis artifacts generated in this study (per-cohort subgroup results, figure source data and a table linking every reported number to its result file) are deposited on Zenodo (doi:10.5281/zenodo.23814933).

### Code

All analysis code will be released at https://github.com/CellularSyntax/ecg-fm-age-disparity under the MIT licence on publication, with the exact version used for this manuscript archived on Zenodo. The repository reproduces every number, table and figure from the deposited artifacts.

### Models

Pretrained backbone weights are used without modification from their public releases (Supplementary Table S4): ECGFounder [4], ECG-FM [5], xECG [6], DeepECG-SSL [7] and TiRex-2 [20].

## Ethics and data-use statement

This study is a secondary analysis of de-identified datasets. MIMIC-IV-ECG, PTB-XL, Shaoxing and CPSC2018 are available via PhysioNet [30] (MIMIC-IV-ECG under the credentialed-access data-use agreement), CODE-15% and IKEM via Zenodo, SPHDB via figshare, HEEDB-MGH and HEEDB-Emory via the Brain Data Science Platform under its data-use agreement, EchoNext via PhysioNet under its restricted-access data-use agreement, and ED-BWH via Nightingale Open Science under its data-use agreement. LEAD (Lung, hEart, sociAl, boDy) is a prospective observational population-based registry (ClinicalTrials.gov NCT01727518) approved by the Ethics Committee of the City of Vienna (EK-11-117-0711). The full CODE and CODE-II cohorts were accessed under institutional data-use agreements. All analyses used de-identified data, no protected health information was accessed and no patient contact occurred. Race/ethnicity categories are those recorded in each source dataset and are used solely to evaluate model performance for disparities.

## Contributors

M.H. conceived and designed the study, developed the software, curated the data, did the analyses, prepared the figures, supervised and administered the project, obtained funding and wrote the first draft. D.L. did analyses and validation, prepared figures and co-wrote the first draft. M.K.B. and F.M. provided and curated data and resources, did validation analyses and contributed to the investigation. All authors interpreted the results and critically revised the report. CRediT roles: conceptualisation, methodology, software, supervision, project administration and funding acquisition, M.H.; validation, formal analysis and investigation, all authors; resources, M.K.B., F.M. and M.H.; data curation, M.K.B. and M.H.; visualisation, D.L. and M.H.; writing (original draft), M.H. and D.L.; writing (review and editing), all authors. M.H. and D.L. directly accessed and verified the underlying data. All authors had full access to all the data in the study and had final responsibility for the decision to submit for publication.

## Acknowledgments

This study was supported by the Austrian Research Promotion Agency (FFG), CARDISENSE project (grant FO999922422). We thank the teams that released the datasets (PhysioNet, Nightingale Open Science, the Brain Data Science Platform, the Telehealth Network of Minas Gerais, and the figshare and Zenodo contributors), the developers of the foundation models evaluated here for releasing weights and code, the LEAD study investigators and participants, and the IKEM data providers.

