## Supplementary_appendix for "ECG foundation models share an age-related performance disparity that originates in the data, not the models"

October 30, 2026

### S1 Supplementary methods

**Cohorts.** ED-BWH (Brigham and Women's Hospital emergency department) is distributed by Nightingale Open Science and stores 2.5 s for each of twelve leads and 10 s for three leads (V1, II, V5). Positive ECGs per age band were counted for every classification endpoint from the out-of-fold prediction files. The 10 endpoints with fewer than 20 positives in the youngest or oldest band (all LEAD endpoints, and AF in Shaoxing, CPSC2018, CODE-II and SPHDB) are listed in Supplementary Table S6 and excluded from all age-gap aggregates; the rule also applies to the adaptation targets built on these cohorts. LEAD (Lung, hEart, sociAl, boDy) is a population-based cohort recruited at 6–80 years of age, so 53% of its ECGs are from participants under 40 and only 419 from participants aged 75 or older; it has 1, 13 and 11 events under 40 years for heart failure, MACE and MI. Its MACE and MI endpoints are therefore reported on the secondary age-gap definition (AUROC at 40–59 minus ≥75 years; 62/218/69 and 48/159/39 positives in the 40–59, 60–74 and ≥75 bands), which is computed for every endpoint in Supplementary Fig. S4. A sensitivity analysis with the threshold at 10 positives is reported in Supplementary Table S6. Curation flows for all thirteen cohorts are shown in Supplementary Figs S1–S3. Pretraining corpora overlap some cohorts at the waveform level (Supplementary Table S3); no model had seen any endpoint label.

**Label types.** Outcomes: ED-BWH 30-day MACE (death, MI or positive troponin), HEEDB-Emory MACE (ICD-based composite), LEAD MACE and CODE-15% 1-year mortality. Coded diagnoses: HEEDB-Emory ICD-coded heart failure and MI, LEAD heart failure. ECG-read diagnoses: PTB-XL MI, conduction disturbance and hypertrophy (SCP statements from ECG reports) and AF (CODE-15%, Shaoxing, CPSC2018). Measurements: EchoNext LVEF $\leq$45% and structural heart disease (echocardiography) and MIMIC-IV laboratory classes. The ED-BWH 12-lead input is a sensitivity analysis of the same endpoint and was not counted. The comparison uses the median pretrained gap per endpoint (Mann–Whitney test over endpoints).

**Data quantity.** For seven endpoints with age-band subgroups (PTB-XL normal ECG, MI, ST/T change, conduction disturbance and hypertrophy; EchoNext LVEF; CODE-15% mortality), patients were split 70/10/20 by a hash of the patient identifier; logistic probes were trained on 100, 300, 1000, 3000, 10 000 and 30 000 labelled ECGs (or the full training set) with the regularisation chosen on the validation split, three seeds per size, and evaluated on the fixed test split. The gap here is overall AUROC minus the worst age-band AUROC (bands with at least 50 ECGs and 5 events). This analysis used ECGFounder and xECG (14 curves).

**Read-out capacity and model changes.** These experiments used held-out patient-level train/test splits (held-out fractions 1.6–10.1% of each cohort). Per-age-band read-outs fitted a separate logistic probe in each band; error-focused reweighting used a focal loss; the non-linear ceiling was a gradient-boosted classifier fitted within each band, and its *avoidable gain* is its band AUROC minus that of the linear probe (10 endpoints per backbone). Low-rank adaptation inserted rank-8 adapters (ECGFounder squeeze-excitation layers, xECG projection layers, ECG-FM feed-forward layers, DeepECG-SSL attention and feed-forward layers), trained for three epochs on capped training sets (20 000–30 000 ECGs) with empirical-risk, group-robust (worst age band) or class-balanced objectives, each with a paired frozen probe on the identical split. The gap here is the maximum minus the minimum band AUROC; 114 adaptation cells over 21 targets were age-stratified, and 30 further runs used two additional seeds for a subset of cells, and group-robust adaptation of PTB-XL MI was run with 5 seeds. Calibration by age (Supplementary Fig. S19C) used the ensemble of the four ECG backbones: expected calibration error [S1] of the pooled calibration map within each band, and after Platt recalibration fitted within each band.

**Case-age by control-age discrimination and operating points.** From the stored out-of-fold predictions of the frozen probes (no refitting), we restricted each endpoint to ECGs scored by all ten networks, averaged predicted probabilities over the four ECG backbones (and, separately, their untrained copies) and computed the AUROC of cases from each age band against controls from each age band, with 200 bootstrap resamples of ECGs within each cell; cells with fewer than 20 cases or controls were not computed. Operating points used the same ensemble. The single threshold was the 10th percentile of case scores (90% sensitivity); band-specific thresholds were the 10th percentile within each band. In the comparison of the two, thresholds were chosen on one half of patients (hash of the patient identifier) and applied to the other, and vice versa.

**Potassium.** We compared Black and White patients' errors within 0.25 mmol/L bins of true potassium, standardised the Black error to the White potassium distribution, adjusted for kidney function (CKD-EPI 2021 eGFR [S2]) in regressions with patient-clustered errors, and refitted probes with White training data down-sampled to the size of the Black training data. These analyses used the stored out-of-fold predictions.

**Embedding geometry.** We described embeddings with UMAP projections [S3], a separability index (silhouette [S4]) and the balanced-accuracy gain of linear probes for age band, sex and race (EchoNext), and characterised a detached cluster in the MIMIC-IV-ECG projection by HDBSCAN clustering and linkage to acquisition metadata.

**Endpoint definitions.** AF is a same-recording rhythm label (CODE, CODE-II diagnostic statements; Shaoxing and CPSC2018 SNOMED codes 164889003 and 49436004). PTB-XL MI is the SCP-ECG MI superclass. LVEF $\leq$45% and structural heart disease (EchoNext) are echocardiography-derived. Potassium, calcium and NT-proBNP (MIMIC-IV) are the closest laboratory value within $\pm$6 h of the ECG; classes are hyperkalaemia $\geq$5.5 and hypokalaemia $<$3.5 mmol/L, hypercalcaemia $>$10.5 and hypocalcaemia $<$8.5 mg/dL, and NT-proBNP $>$900 pg/mL (a single cut-off, so that the label does not depend on age). ED-BWH MACE is the 30-day composite of death, MI or positive troponin; HEEDB-Emory MACE, MI and heart failure are ICD-based; mortality endpoints are all-cause.

**Implementation details.** ECG-FM and DeepECG-SSL are Transformer models pretrained with a masked-prediction objective in the wav2vec 2.0 framework and implemented in fairseq-signals, whose forward pass applies random time masking during training. All embeddings, untrained-copy embeddings, adapted models and saliency maps were computed with this masking disabled (`mask=False`; for DeepECG-SSL also `features_only=True`) and with deterministic GPU kernels; two independent extractions of 200 MIMIC-IV-ECG records gave bit-identical embeddings for all four ECG backbones.

**ED-BWH inputs and DeepECG-SSL tiling.** The primary ED-BWH input is the three leads recorded for 10 s (V1, II, V5), placed at their canonical positions with the other leads set to zero. The sensitivity analysis uses the 2.5 s segments of all twelve leads, tiled to 10 s, for all backbones.

**Large cohorts.** For cohorts with more than two million ECGs, probes were fitted on a patient-grouped random subset of about 1.2 million ECGs; this applies to HEEDB-MGH (1,197,523 of 10,608,417 ECGs). CODE was analysed at full size (2,322,465 ECGs); above 400,000 training rows the logistic probe was fitted by stochastic gradient descent.

**Untrained networks.** Each architecture was constructed from its configuration without loading weights; all parameters were re-initialised (PyTorch default initialisers, Xavier-uniform or $\mathcal{N}(0, 0.02)$ for remaining tensors) with one fixed seed per architecture, and the parameters were identical on all GPU types. Preprocessing, pooling and probes were identical to the pretrained models. The 20 endpoints are from PTB-XL, EchoNext, MIMIC-IV, ED-BWH (both inputs), Shaoxing, CPSC2018, CODE-15%, LEAD and HEEDB-Emory. The pair definition (endpoint, backbone, both age-band results present, finite gap) is used for every correlation and count; Pearson and Spearman correlations are reported for all five backbones (primary), the four ECG backbones and classification endpoints only.

**Second non-ECG reference and additional seeds.** MOMENT-1-large [S5] (T5 encoder, 24 layers, 1,024-d, 341 million parameters in the embedding configuration; Hugging Face `AutonLab/MOMENT-1-large`, commit `ca58581b`, MIT licence; pretrained on the Time Series Pile, no ECG pretraining) was run with the TiRex-2 preprocessing (canonical 12-lead signal at 500 Hz, fixed to 10 s, decimated to 100 Hz). MOMENT encodes each channel independently in fixed windows of 512 samples, so each lead was embedded in two windows (samples 1–512 and 513–1,000, the second zero-padded with the padding masked), the model's own embedding head (masked mean over patches) was applied and the embeddings were averaged over windows and leads to one 1,024-d vector; TiRex-2, by contrast, sees the 12 leads jointly as one multivariate series. The encoder ran in bf16 autocast (embedding correlation with fp32 0.999997 on 96 PTB-XL ECGs). Its untrained copy was built with the same re-initialisation rule as the other untrained networks (seed 0; all 341 million parameters redrawn, including the RMS-norm weights). Seeds 1 and 2 of the untrained ECGFounder, xECG, ECG-FM, DeepECG-SSL and TiRex-2 used the same rule with `torch.manual_seed(S)`; parameter checksums differ by seed and are identical across cohorts and GPU types. All these runs used patient-grouped subsets fixed before any result was seen: ECGs with a usable label for at least one control endpoint (ED-BWH restricted to the eligible visits used in the paper), and, where more than 100,000 remained, whole patients ordered by a fixed hash of the patient identifier until 100,000 ECGs were reached (CODE-15% 345,779 → 100,000 ECGs, MIMIC-IV 506,145 → 100,000, HEEDB-Emory 981,767 → 100,000; the other six cohorts were used in full). The pretrained backbones and the seed-0 untrained copies were re-probed on exactly the same ECGs with the unmodified probe code, so all 22 model variants of an endpoint share folds and bootstrap resamples; on the uncapped cohorts the re-run reproduces Table S8 (median absolute difference 0.0001 in overall AUROC and in age gap). The campaign used 19.4 GPU-hours. Of the 22 control endpoints, 17 pass the small-$n$ rule on these subsets (failing: CPSC2018 AF, Shaoxing AF, LEAD MACE, MI and heart failure).

**Classical-measurement models.** Features were the GE MUSE measurements in MIMIC-IV-ECG (20 features), the full GE 12SL measurement set in PTB-XL (782 features; 23-feature global set as sensitivity), and intervals, amplitudes, axes and heart-rate variability from an automatic delineator (47–48 features) in other cohorts; vendor sentinel values were treated as missing, with missingness indicators. Logistic regression used median imputation and standardisation within training folds; LightGBM [S6] used native missing-value handling with early stopping on an inner patient-grouped split. Folds were identical to those of the foundation-model probes. The balanced-band sensitivity analysis refitted each model on a random subsample with equal numbers of ECGs per age band.

**Ageing versus disease.** Flags came from SCP or SNOMED codes (PTB-XL, Shaoxing, CPSC2018), machine statements (MIMIC-IV, HEEDB-Emory), diagnostic labels (CODE) or the AF label (ED-BWH); EchoNext and LEAD have no rhythm labels, and a wide-QRS ($\geq$120 ms) delineator proxy was used for EchoNext. The target condition was never excluded. Changes in gap are paired bootstrap differences on the same resamples. Covariate adjustment regressed per-ECG absolute error of the backbone ensemble on age band with and without clinical covariates (LEAD, ED-BWH, MIMIC-IV).

**UMAP island.** UMAP projections were computed for the same 8,000 MIMIC-IV-ECG test ECGs for each backbone. HDBSCAN (minimum cluster size 100) identified the detached ECG-FM cluster (1,134 ECGs, 14% of the sample). Cluster ECGs were compared with the rest of the sample for acquisition metadata (cart identifier, filter settings), machine rhythm statements, demographics and labels (95% Wilson CIs). Isolation of the cluster in each backbone was summarised by 15-nearest-neighbour purity in the full embedding space and by the five-fold AUROC of a linear probe for cluster membership.

**Saliency.** Saliency was computed with SmoothGrad [S7] (16 noise samples, noise SD 0.1 times the signal SD) for the four ECG backbones; TiRex-2 has no input gradient because its patch tokeniser detaches the input. Eight endpoints were analysed (PTB-XL MI; EchoNext LVEF≤45% and structural heart disease; MIMIC-IV-ECG hyper- and hypokalaemia, hyper- and hypocalcaemia and elevated NT-proBNP) on 250 shared test ECGs per cohort, giving 30 endpoint–backbone cells (DeepECG-SSL does not run on EchoNext). Beats were delineated on lead II with NeuroKit2 (discrete wavelet method), and saliency was summarised in three windows: R-aligned −300 to +300 ms (standard window), R-aligned −450 to +350 ms (extended window, primary) and P-onset-aligned −100 to +300 ms. Relative density is the mean absolute saliency within a wave divided by the mean over the window. Old-minus-young differences were pooled over cells with 1,000 bootstrap resamples of beats and, for the reported CIs, of ECGs (cluster bootstrap); the per-beat interval for the extended window was 0.078–0.098. ED-BWH could not be delineated and is not included.

**Wave blanking.** The delineated P wave, QRS complex or T wave was set to zero in all leads, embeddings were extracted from the blanked ECGs and the logistic probe was refitted (five-fold, out-of-fold predictions). Seven endpoint cells were used per backbone: PTB-XL MI (up to 8,000 ECGs), CODE-15% right bundle-branch block and HEEDB-Emory heart failure and MACE (up to 4,000 ECGs each), with AF and paced ECGs removed; and AF in CODE-15%, CPSC2018 and Shaoxing (up to 4,000 ECGs, no exclusion). The age gap in this analysis is overall AUROC minus the AUROC of the worst age band (bands with at least 25 positive and 25 negative ECGs). Drops are unblanked minus blanked AUROC with paired bootstrap CIs, pooled over 28 cells for the four ECG backbones and over 35 cells with TiRex-2. For TiRex-2, records shorter than 10 s were tiled rather than zero-padded in this experiment.

### S2 Supplementary results

**Overall discrimination and race.** On HEEDB-Emory, AUROC was 0.75–0.77 for MACE, 0.73–0.76 for MI and 0.79–0.81 for heart failure across the four ECG backbones, and for 30-day MACE in ED-BWH 0.75–0.83 (12-lead input 0.74–0.77; below 40 years 0.82–0.85, at 75 years or older 0.63–0.66). On LEAD, ECGFounder was weaker than the other backbones (heart failure 0.61 *vs* 0.81–0.83). In HEEDB-Emory MI, AUROC fell from 0.77–0.79 below 40 years to 0.64–0.68 at 75 years or older. The worst-served race group changed between endpoints (White in 31, Asian in 24, Black in 16 of 102 pretrained results).

**Secondary age-gap definition, LEAD and the small-$n$ rule.** The secondary gap (AUROC at 40–59 minus ≥75 years) was computed for every classification endpoint and model from the stored out-of-fold predictions with 1,000 ECG-level bootstrap resamples within band. Over the 16 main-text endpoints, the medians over pretrained backbones of the two definitions correlated at Spearman $\rho$=0.94 (Pearson $r$=0.93; median primary gap 0.053, median secondary gap 0.040; all 16 secondary gaps positive). LEAD MACE and MI have 62/218/69 and 48/159/39 positive ECGs in the 40–59, 60–74 and ≥75 bands. Their secondary gaps were −0.042 to 0.108 (median 0.010) and −0.011 to 0.080 (median 0.013) across the five backbones, with no CI excluding zero (CIs span roughly −0.15 to 0.22), whereas on the same definition HEEDB-Emory MI lost 0.102–0.120, HEEDB-Emory heart failure 0.112–0.128 and ED-BWH 30-day MACE 0.090–0.104 AUROC, every CI excluding zero (Supplementary Fig. S4). LEAD heart failure (75 events, 9 in the 40–59 band) fails both definitions. LEAD sex gaps (|AUROC female − male|) were 0.02–0.14 (median 0.05) over 15 endpoint–backbone results, with the female AUROC lower in 13 of 15 and the CI excluding zero in 5 of 15. Relaxing the small-$n$ threshold from 20 to 10 positives admits CPSC2018 AF and LEAD MACE and MI and changes the median worst-group age gap from 0.028 to 0.031 (no rule: 0.033), the share of results with age gap larger than sex gap from 97% to 96%, the untrained–pretrained correlation (five original networks, classification pairs) from 0.89 to 0.83 (no rule: 0.85) and the label-type contrast p from 0.0005 to 0.007 (no rule: 0.006); the sex and race medians are unchanged (Supplementary Table S7). Holm adjustment within families changes the status of one test: low-rank adaptation narrowed the age gap with adjusted $p<0.05$ in two rather than three of the four backbones (DeepECG-SSL raw p=0.009, adjusted 0.063; Supplementary Table S5).

**Untrained networks.** Fig. 5B and the correlations quoted in the main text use classification (AUROC) pairs only, so that every gap is an AUROC difference on one scale: r=0.81 over 102 pairs of the six networks (Spearman $\rho$=0.80); five original networks on the full cohorts r=0.89 (85 pairs); four ECG backbones r=0.90 (68 pairs); adding the 17 MOMENT-1-large pairs from the capped subsets lowers the pooled value because MOMENT's own untrained–pretrained agreement is weaker (below). The 117 probed pairs also include 15 regression pairs (potassium, calcium and log NT-proBNP in MIMIC-IV; gap = MAE at ≥75 minus <40 years, in each endpoint's units; Supplementary Fig. S12). Untrained and pretrained copies agree closely on them too (potassium 0.035–0.059 *vs* 0.034–0.051; calcium −0.056 to −0.040 *vs* −0.043 to −0.013; log NT-proBNP −0.63 to −0.43 *vs* −0.44 to −0.21), but because the NT-proBNP differences are an order of magnitude larger than any AUROC gap they dominate a pooled Pearson r: over all 117 pairs r=0.90 (five networks: 0.93 over 100), and 0.83 without the five NT-proBNP pairs. We therefore report the classification-only value. The untrained gap was positive and significant in 68 of the 85 five-network classification pairs (pretrained: 60). Further examples: Shaoxing AF nan pretrained *vs* nan untrained; HEEDB-Emory heart failure 0.142 *vs* 0.151 (Supplementary Fig. S11).

**Second non-ECG reference and seeds.** On the capped subsets, pretrained MOMENT-1-large reached a median AUROC of 0.776 (four ECG backbones on the same ECGs 0.800, TiRex-2 0.768) and its age gaps correlated with the median pretrained ECG gap at Pearson r=0.94 (Spearman $\rho$=0.87; 17 endpoints passing the small-$n$ rule; r=0.97 against the four backbones re-probed on the same ECGs, r=0.94 over all 22 endpoints); TiRex-2 on the same subsets gave r=0.96. The untrained MOMENT copy had a median AUROC of 0.663 and median gap 0.048, and correlated with the median pretrained ECG gap at r=0.54 ($\rho$=0.50; r=0.77 over all 22 endpoints) and with MOMENT's own pretrained gaps at r=0.45 (17 endpoints; 0.80 over all 22); the departures are concentrated in ED-BWH MACE (untrained gap 0.03 versus 0.18–0.21 for every other model) and CODE-15% AF. Across seeds 0–2 of the five untrained networks (85 endpoint–architecture pairs), the pooled untrained–pretrained correlation was r=0.87, 0.88 and 0.89, the median untrained gap 0.056, 0.063 and 0.068, and the across-seed SD of the gap per endpoint had a median of 0.007 (ECGFounder), 0.007 (xECG), 0.009 (ECG-FM), 0.007 (DeepECG-SSL) and 0.008 (TiRex-2) against a between-endpoint SD of the seed-mean gap of 0.054–0.067; the largest seed SD (0.068) was MIMIC-IV hypercalcaemia for ECG-FM, the endpoint with the fewest positives under 40 years (41). The sign of the gap was the same for all three seeds in 74 of 85 pairs (Supplementary Fig. S16). Because MOMENT-1-large was evaluated only under this control protocol, it appears in the figures that compare models by age band (Figs 5, S4, S11, S12, S13, S14 and S16; capped subsets marked) but not in analyses that require quantities it does not have: saliency and wave blanking (not computed for MOMENT), low-rank adaptation and learning curves, potassium regression, the out-of-fold case–control matrices and the ensemble of the four ECG backbones.

**Classical measurements.** Over the 16 endpoints passing the small-n rule, logistic regression gave Spearman $\rho$=0.90 (p=2e-06) and gradient-boosted trees $\rho$=0.91 (p=7e-07; median gap 0.054). Including the endpoints that fail the rule gave $\rho$=0.94 and 0.65 (19 endpoints); the lower value for boosted trees came from LEAD heart failure, whose youngest band contains 1 event. Classical age gaps were unchanged with equal numbers of ECGs per band ($\rho$ between full and balanced gaps 0.99 and 0.97; 16 endpoints). With the full PTB-XL measurement set, boosted trees reached an MI AUROC of 0.948 (backbones 0.878–0.922) and still showed the age gap (Supplementary Figs S13, S14 and S15).

**Label types.** Median pretrained age gap by group: outcomes 0.176, coded diagnoses 0.126, ECG-read diagnoses 0.031, measurements 0.030.

**Co-existing disease and data quantity.** AF, pacing and bundle-branch block were much more common in older patients (AF in MIMIC-IV 1.7% under 40 years *vs* 23.0% at 75 years or older; Shaoxing 1.5% *vs* 51.3%). After their removal the median change in gap was −0.003; the largest consistent reduction, −0.015 to −0.014, was in HEEDB-Emory heart failure (Supplementary Fig. S21). Measured comorbidity and severity explained 28–51% of the age trend in per-ECG error, but never all of it. Older patients were not scarce: in HEEDB-Emory the oldest band contained 205,701 ECGs and the youngest 104,149. Across the label-efficiency curves, the largest change in gap between 3000 labels and the full set was 0.035.

**Model-side changes.** Per-band read-outs changed overall AUROC by −0.018, focal reweighting by −0.171, group-robust adaptation by −0.005 and class-balanced adaptation by +0.000 (medians, four ECG backbones). Mean avoidable gains of the non-linear read-out at 75 years or older were −0.001 (ECGFounder), −0.012 (xECG), −0.025 (ECG-FM) and −0.013 (DeepECG-SSL); the largest single gain was +0.049 (ECGFounder). Across the 114 age-stratified adaptation cells, the gap narrowed by more than 0.005 in 62 and widened in 20; AUROC at 75 years or older rose in 100% of ECGFounder, 97% of xECG, 100% of ECG-FM and 100% of DeepECG-SSL cells (31 ECG-FM and 21 DeepECG-SSL cells).

**Potassium.** The largest race difference followed the label distribution. Potassium MAE was higher in Black than in White patients in all five backbones (0.51–0.55 *vs* 0.43–0.46 mmol/L). True potassium varied more in Black patients (SD 0.85 *vs* 0.69 mmol/L), and the probes shrink predictions towards the mean. At matched true potassium the difference fell from +0.074–0.084 to 0.001–0.006 mmol/L (Supplementary Fig. S5B). True potassium ≥5.5 mmol/L occurred in 8.3% of Black and 5.0% of White patients; matched-true-potassium standardisation explained 92–99% of the gap. Adjusting for eGFR did not reduce the remaining difference (xECG: +0.079 unadjusted, +0.005 after adjusting for true potassium, +0.012 after adding eGFR). Down-sampling White training data to the Black sample size left the gap almost unchanged (xECG 0.074 to 0.071), and probes trained only on Black patients did not reduce Black patients' error (0.520 *vs* 0.511; Supplementary Fig. S5).

**Embeddings and acquisition device.** Age varied smoothly across the projection while sex and race were intermixed (Fig. 3; separability 0.28 for age, 0.10 for sex, 0.11 for race), yet sex was decoded at least as well as age (balanced-accuracy gain 0.19–0.25 *vs* 0.16–0.22; race 0.03–0.07) and produced almost no gap. The detached cluster in the ECG-FM projection (1,134 of 8,000 ECGs) is an acquisition-device group: 99.1% of its ECGs came from one ECG cart with a 0.0005–150 Hz bandwidth setting, compared with 6.3% elsewhere. It differed demographically (women 57.2% *vs* 45.6%; Black patients 20.6% *vs* 14.2%), was younger than the rest of the cohort and does not explain the age gap (Supplementary Fig. S6).

**Attention.** In saliency maps of the four ECG backbones (30 endpoint–backbone cells), the P wave received relatively more attention in older patients (old minus young relative P-wave density +0.088, 95% CI 0.055 to 0.099), driven by ECGFounder (+0.183) and xECG (+0.102); ECG-FM (−0.006) and DeepECG-SSL (+0.001) showed none (Fig. S7). The standard analysis window rarely cut the P wave (P onset before −300 ms in 0.4–0.7% of beats under 40 years and 2.1–2.5% at 75 years or older), and the difference was similar with that window (+0.089) and after alignment on P-wave onset (+0.072); per-ECG cluster-bootstrap CIs led to the same conclusion as the per-beat CIs. Attention did not move within the P wave (centroid 0.494 *vs* 0.490 of P duration), and the QRS complex showed no age difference (+0.008). Setting the P wave to zero changed the age gap (overall minus worst band) by −0.0001 (−0.0044 to 0.0037), whereas setting the QRS complex to zero lowered AUROC by 0.0307 (Supplementary Fig. S8). The P-wave shift is a correlate of age in two backbones, not a cause of the gap. Saliency was analysed for the four ECG backbones on eight endpoints (TiRex-2 has no input gradient); the causal test, wave blanking, includes all five networks.

### S3 Supplementary tables

**Table S1:** Study cohorts (thirteen cohorts on four continents). *Available*: ECGs and unique patients in the cohort manifest. *Analysed*: the largest endpoint analysis set after curation and the unique patients contributing it; HEEDB-MGH was probed on a patient-grouped subsample. CODE-15% is a subset of CODE and the ED-BWH 3-lead and 12-lead recordings come from the same visits, so each is counted once. All backbones are used as fixed feature extractors; no endpoint label was seen during pretraining (waveform-level overlap in Supplementary Table S3). *Race-lab.* is the fraction with a race label, present only in the US cohorts. Cohorts are analysed stratified and never pooled.

| | | Available | | Analysed | | | | |
|---|---|---|---|---|---|---|---|---|
| Cohort | Region | ECGs | Patients | ECGs | Patients | Female% | Median age | Race-lab. |
| HEEDB-MGH | US | 10,608,417 | 1,818,247 | 1,197,523 | 247,318 | 47.0 | 64 | 86% |
| CODE | S. America | 2,829,840 | 1,558,748 | 2,322,465 | 1,558,748 | 49.5 | 54 | – |
| HEEDB-Emory | US | 981,767 | 344,075 | 981,767 | 344,075 | 48.5 | 63 | 100% |
| MIMIC-IV | US | 506,145 | 136,663 | 506,145 | 136,663 | 48.9 | 66 | 92% |
| EchoNext | US | 100,000 | 36,286 | 100,000 | 36,286 | 46.4 | 63 | 88% |
| IKEM | Europe | 98,130 | 30,290 | 98,130 | 30,290 | 34.4 | – | – |
| ED-BWH | US | 103,952 | 44,713 | 61,680 | 30,933 | 55.6 | 52 | 100% |
| Shaoxing | Asia | 45,130 | 45,130 | 45,130 | 45,130 | 43.6 | 61 | – |
| SPHDB | Asia | 25,770 | 24,666 | 25,770 | 24,666 | 44.6 | – | – |
| PTB-XL | Europe | 21,799 | 18,869 | 21,799 | 18,869 | 47.9 | 62 | – |
| CODE-II | S. America | 15,000 | 15,000 | 15,000 | 15,000 | 52.9 | 56 | – |
| LEAD | Europe | 14,951 | 14,866 | 14,689 | 14,607 | 52.7 | – | – |
| CPSC2018 | Asia | 6,877 | 6,877 | 6,877 | 6,877 | 46.2 | 64 | – |
| Total | | 15,357,778 | 4,094,430 | 5,396,975 | 2,509,462 | | | |

**Table S2:** Clinical endpoints and prevalence per cohort (all thirteen cohorts; thirty-one endpoints). Type abbreviations: clf, binary classification; reg, continuous regression; surv, survival (time-to-event) analysis. Prevalence is positives over the labelled analysis set for that endpoint. †ED-BWH and ‡LEAD are held-out external cohorts not seen in any model's pretraining; LEAD (Europe) and Emory/ED-BWH (US) share the MACE/MI/HF endpoint family for external replication.

| **Cohort** | **Endpoint** | **Type** | **Positives / N** |
|---|---|---|---|
| HEEDB-MGH | 1-y mortality | surv | 2,035,671 / 10,608,417 (19.2%) |
| HEEDB-Emory | MACE (composite) | clf | 502,124 / 981,767 (51.1%) |
| | Myocardial infarction (ICD) | clf | 149,618 / 981,767 (15.2%) |
| | Heart failure (ICD) | clf | 361,665 / 981,767 (36.8%) |
| | Mortality | clf | 130,354 / 981,767 (13.3%) |
| PTB-XL | Myocardial infarction (superclass) | clf | 5,469 / 21,799 (25.1%) |
| EchoNext | LVEF ≤45% | clf | 23,892 / 100,000 (23.9%) |
| | Structural heart disease | clf | 52,188 / 100,000 (52.2%) |
| MIMIC-IV | Hyperkalemia (K≥5.5 mmol/L) | clf | 28,288 / 506,145 (5.6%) |
| | Hypokalaemia (K<3.5 mmol/L) | clf | 54,623 / 506,145 (10.8%) |
| | Hypercalcemia (Ca>10.5 mg/dL) | clf | 4,269 / 265,007 (1.6%) |
| | Hypocalcemia (Ca<8.5 mg/dL) | clf | 73,855 / 265,007 (27.9%) |
| | Elevated NT-proBNP (>900 pg/mL) | clf | 26,122 / 41,841 (62.4%) |
| | Serum $K^+$ (regression) | reg | n=506,145; mean 4.25 (SD 0.72) mmol/L |
| | Calcium (regression) | reg | n=265,007; mean 8.87 (SD 0.82) mg/dL |
| | NT-proBNP (log, regression) | reg | n=41,841; median 1,692 (IQR 393–5,529) pg/mL |
| CODE | AF (dx) | clf | 41,851 / 2,322,465 (1.8%) |
| CODE-15% | 1-y mortality | surv | 8,341 / 233,647 (3.6%) |
| CODE-II | AF (dx) | clf | 248 / 15,000 (1.7%) |
| Shaoxing | AF (SNOMED) | clf | 1,780 / 45,130 (3.9%) |
| CPSC2018 | AF (SNOMED) | clf | 1,221 / 6,877 (17.8%) |
| ED-BWH† | MACE / troponin+ (30 d) | clf | 3,422 / 61,680 (5.5%) |
| LEAD‡ | MACE | clf | 362 / 14,951 (2.4%) |
| | Myocardial infarction | clf | 257 / 14,951 (1.7%) |
| | Heart failure | clf | 75 / 14,951 (0.5%) |
| IKEM | Tachycardia | clf | 9,317 / 98,130 (9.5%) |
| SPHDB | AF | clf | 460 / 25,770 (1.8%) |
| | Left-ventricular hypertrophy | clf | 209 / 25,770 (0.8%) |
| | Premature ventricular contraction | clf | 425 / 25,770 (1.6%) |
| | Right bundle branch block | clf | 710 / 25,770 (2.8%) |
| | ST deviation | clf | 1,833 / 25,770 (7.1%) |

**Table S3:** Pretraining–evaluation overlap at the waveform level. ✓ marks a cohort whose waveforms are included in that model's pretraining corpus (a *label* was still never seen: every probe is trained only on frozen embeddings, so all evaluations are zero-shot on the target task). ECGFounder's proprietary HEEDB corpus is the source of the HEEDB-MGH and HEEDB-Emory cohorts, so those two overlap ECGFounder at the waveform level. †Six cohorts are *truly external* — in no model's pretraining — with LEAD, ED-BWH and Emory anchoring the external replication of the age gap. The non-ECG reference TiRex-2 has no ECG pretraining and therefore overlaps no cohort; the paradigm reading is anchored on the concordance across TiRex-2 and the truly-external cohorts, not on overlapping backbones alone.

| Cohort | ECGFounder | xECG | ECG-FM | DeepECG-SSL |
|---|---|---|---|---|
| HEEDB-MGH | ✓ | | | |
| HEEDB-Emory | ✓ | | | |
| CODE | | ✓ | | ✓ |
| MIMIC-IV | | | ✓ | ✓ |
| Shaoxing | | ✓ | ✓ | |
| PTB-XL | | | ✓ | |
| CPSC2018 | | | ✓ | |
| CODE-II† | | | | |
| EchoNext† | | | | |
| IKEM† | | | | |
| ED-BWH† | | | | |
| SPHDB† | | | | |
| LEAD† | | | | |

**Table S4: Backbones used as frozen feature extractors.** All weights were used as released, without modification. Embeddings were extracted once with deterministic inference and kept fixed; the input is the cohort ECG resampled to the model's native sampling rate with the model's native preprocessing. Parameter counts are those of the loaded models.

| Model | Reference | Architecture (parameters) | Pretraining objective and data | Native input | Embedding | Weights (file; source; licence) |
|---|---|---|---|---|---|---|
| ECGFounder | [S8] | 1-D convolutional network, Net1D (30.7 M) | Supervised, 150 diagnostic labels, >10 million HEEDB ECGs | 12 leads, 500 Hz, 10 s | 1,024-d, penultimate layer | `12_lead_ECGFounder.pth`; github.com/PKUDigitalHealth/ECGFounder, HF `PKUDigitalHealth/ECGFounder`; MIT |
| xECG | [S9] | xLSTM, patch size 25 (57.0 M) | Self-supervised (SimDINOv2), ∼8 million ECGs from 1.7 million patients | 12 leads, 100 Hz, 10 s | 1,024-d, class token | `model.safetensors`; HF `riccardolunelli/xECG_base_model_v1`; MIT |
| ECG-FM | [S10] | Transformer encoder, wav2vec 2.0 framework, 12 layers (90.9 M) | Self-supervised (W2V + CMSC + RLM), MIMIC-IV-ECG and PhysioNet 2021 (∼1.25 million ECGs) | 12 leads, 500 Hz, 5,000 samples | 768-d, masked mean over time | `mimic_iv_ecg_physionet_pretrained.pt`; HF `wanglab/ecg-fm`, github.com/bowang-lab/ecg-fm; MIT |
| DeepECG-SSL | [S11] | Transformer encoder, wav2vec 2.0 CMSC framework, 12 layers (90.4 M) | Self-supervised (masked prediction), MHI, MIMIC-IV and CODE-15% ECGs | 12 leads, 250 Hz, 10 s | 768-d, layer 10, masked mean over time | `base_ssl.pt`; HeartWise-AI (github.com/HeartWise-AI/DeepECG_Docker, HF `heartwise`); licence Apache-2.0 |
| TiRex-2 | [S12] | xLSTM universal time-series model with multivariate blocks (82.5 M) | Zero-shot forecasting pretraining on public time-series corpora; no ECG | 12-lead multivariate series, 100 Hz, 1,000 samples | 512-d, mean-pooled hidden state of block `stack.2` | HF `NX-AI/TiRex-2` (commit `30788a31`); licence Apache-2.0 |
| MOMENT-1-large | [S5] | Transformer encoder (T5, 24 layers, 1,024-d), channel-independent (341 M) | Masked patch reconstruction on the Time Series Pile; no ECG | univariate windows of 512 samples; 12 leads × 2 windows at 100 Hz | 1,024-d, mean over patches, windows and leads | HF `AutonLab/MOMENT-1-large` (commit `ca58581b`); MIT |

**Table S5: Hypothesis tests, with Holm adjustment within each pre-defined family.** Raw p-values are those printed in the main text and figure legends (recomputed from the result files); the comorbidity-exclusion test and the avoidable-gain tests are listed for completeness although the text reports only their point estimates and bootstrap CIs. Holm adjustment is within family (family size given by the number of rows sharing a family). One test changes status at $\alpha$=0.05: low-rank adaptation of DeepECG-SSL (raw p=0.009, adjusted 0.063), so two rather than three backbones show an individually significant change in age gap under LoRA.

| Family | Comparison | Test | $n$ | Raw p | Holm p | Significant |
|---|---|---|---|---|---|---|
| Mitigation strategies (gap change) | Per-band read-outs: change in age gap vs paired standard probe | Wilcoxon signed-rank, two-sided | 40 | 0.67 | 1.0 | no |
| Mitigation strategies (gap change) | Error-focused (focal) reweighting: change in age gap vs paired standard probe | Wilcoxon signed-rank, two-sided | 40 | 0.69 | 1.0 | no |
| Mitigation strategies (gap change) | Group-robust training: change in age gap vs paired standard probe | Wilcoxon signed-rank, two-sided | 54 | 0.20 | 1.0 | no |
| Mitigation strategies (gap change) | Class-balanced training: change in age gap vs paired standard probe | Wilcoxon signed-rank, two-sided | 44 | 0.59 | 1.0 | no |
| Mitigation strategies (gap change) | LoRA (ECGFounder): change in age gap vs frozen probe, per target | Wilcoxon signed-rank, two-sided | 21 | 0.0021 | 0.019 | yes |
| Mitigation strategies (gap change) | LoRA (xECG): change in age gap vs frozen probe, per target | Wilcoxon signed-rank, two-sided | 21 | 0.29 | 1.0 | no |
| Mitigation strategies (gap change) | LoRA (ECG-FM): change in age gap vs frozen probe, per target | Wilcoxon signed-rank, two-sided | 21 | 0.0026 | 0.021 | yes |
| Mitigation strategies (gap change) | LoRA (DeepECG-SSL): change in age gap vs frozen probe, per target | Wilcoxon signed-rank, two-sided | 21 | 0.009 | 0.063 | no (raw <0.05) |
| Mitigation strategies (gap change) | Label quantity (3,000 ECGs vs full set): change in age gap | Wilcoxon signed-rank, two-sided | 14 | 0.091 | 0.54 | no |
| Label quantity (AUROC change) | Label quantity (3,000 ECGs vs full set): change in overall AUROC | Wilcoxon signed-rank, two-sided | 14 | 0.0001 | 0.0001 | yes |
| Mitigation strategies (gap change) | Comorbidity exclusion (AF/paced/BBB removed): change in age gap | Wilcoxon signed-rank, two-sided (computed here; text reports median and bootstrap CI only) | 149 | <0.0001 | <0.0001 | yes |
| Label type | Label type: outcome/coded vs ECG-read/measured median pretrained age gap | Mann-Whitney U, two-sided | 16 | 0.0005 | 0.0005 | yes |
| Control concordances (correlations) | Untrained vs pretrained age gap, Pearson r (100 pairs) | Pearson correlation test | 100 | <0.0001 | <0.0001 | yes |
| Control concordances (correlations) | Untrained vs pretrained age gap, Spearman rho (100 pairs) | Spearman correlation test | 100 | <0.0001 | <0.0001 | yes |
| Control concordances (correlations) | Classical logistic gap vs median pretrained gap, Spearman rho (16 endpoints) | Spearman correlation test | 16 | <0.0001 | <0.0001 | yes |
| Control concordances (correlations) | Classical GBM gap vs median pretrained gap, Spearman rho (16 endpoints) | Spearman correlation test | 16 | <0.0001 | <0.0001 | yes |
| Avoidable gain at $\geq$75 (Fig. 4D; p not printed in text) | Per-band GBM read-out avoidable gain at $\geq$75 vs 0 (ECGFounder) | Wilcoxon signed-rank vs 0, two-sided | 10 | 0.92 | 0.92 | no |
| Avoidable gain at $\geq$75 (Fig. 4D; p not printed in text) | Per-band GBM read-out avoidable gain at $\geq$75 vs 0 (xECG) | Wilcoxon signed-rank vs 0, two-sided | 10 | 0.037 | 0.11 | no (raw <0.05) |
| Avoidable gain at $\geq$75 (Fig. 4D; p not printed in text) | Per-band GBM read-out avoidable gain at $\geq$75 vs 0 (ECG-FM) | Wilcoxon signed-rank vs 0, two-sided | 10 | 0.002 | 0.0078 | yes |
| Avoidable gain at $\geq$75 (Fig. 4D; p not printed in text) | Per-band GBM read-out avoidable gain at $\geq$75 vs 0 (DeepECG-SSL) | Wilcoxon signed-rank vs 0, two-sided | 10 | 0.049 | 0.11 | no (raw <0.05) |

**Table S6: Endpoints excluded from the age-gap aggregates by the pre-specified small-n rule.** Age-band results are interpreted only where the youngest (<40 years) and the oldest (≥75 years) band each contain at least 20 positive ECGs (smallest count over backbones, from the out-of-fold prediction files). The endpoints below fail the rule; they are shown greyed in Supplementary Fig. S17A and with open markers in Fig. 5C, and are excluded from every summary statistic, correlation and test of age gaps.

| | | Positive ECGs by age band | | | |
|---|---|---|---|---|---|
| Cohort | Endpoint | <40 | 40–59 | 60–74 | ≥75 |
| CODE-II | AF (diagnostic statement) | 9 | 38 | 100 | 101 |
| CODE-II | Atrial fibrillation | 9 | 38 | 100 | 101 |
| CPSC2018 | AF (SNOMED) | 15 | 165 | 458 | 582 |
| CPSC2018 | Atrial fibrillation | 15 | 136 | 360 | 464 |
| LEAD | Heart failure | 1 | 9 | 43 | 22 |
| LEAD | MACE | 13 | 62 | 218 | 69 |
| LEAD | Myocardial infarction | 11 | 48 | 159 | 39 |
| SPHDB | Atrial fibrillation | 6 | 112 | 217 | 125 |
| Shaoxing | AF (SNOMED) | 8 | 192 | 671 | 909 |
| Shaoxing | Atrial fibrillation | 8 | 192 | 671 | 909 |

**Table S7: Sensitivity of the main-text summary statistics to the small-$n$ rule.** Each statistic was recomputed with the positive-count threshold at 20 (pre-specified; reproduces every published number), at 10 (admits CPSC2018 AF and LEAD MACE and MI) and with no rule. No conclusion changes.

| Statistic | Threshold 20 | Threshold 10 | No rule |
|---|---|---|---|
| Median worst-group gap, age | 0.028 | 0.031 | 0.033 |
| Median worst-group gap, sex | 0.004 | 0.004 | 0.004 |
| Median worst-group gap, race | 0.016 | 0.016 | 0.016 |
| Results with age gap > sex gap (%) | 97 | 96 | 96 |
| of $n$ results | 309 | 337 | 374 |
| Endpoints with AUROC declining toward ≥75 | 12 | 12 | 14 |
| of $n$ classification endpoints | 16 | 18 | 20 |
| Untrained vs pretrained gap, Pearson $r$ (five networks, classification pairs) | 0.89 | 0.83 | 0.85 |
| pairs | 85 | 95 | 105 |
| Classical logistic vs pretrained gap, Spearman $\rho$ | 0.90 | 0.92 | 0.94 |
| Classical GBM vs pretrained gap, Spearman $\rho$ | 0.91 | 0.93 | 0.65 |
| Median age gap, outcome and coded labels | 0.142 | 0.126 | 0.142 |
| Median age gap, ECG-read and measured labels | 0.030 | 0.015 | 0.030 |
| Label-type contrast, Mann–Whitney p | 0.0005 | 0.0069 | 0.0063 |

**Table S8: Discrimination and age gap per endpoint for pretrained, untrained and classical models.** Frozen-embedding probes, patient-grouped five-fold out-of-fold prediction. *Label*: how the label is obtained (outcome; coded diagnosis; diagnosis read from the ECG; measurement). *Overall*: AUROC (MAE for the three regression rows), median (range) over the four ECG foundation models and TiRex-2. *Age gap*: AUROC at <40 years minus AUROC at ≥75 years (MAE rows: MAE at ≥75 minus MAE at <40), so that a positive value means worse performance in the oldest; median (range) over backbones. *Untrained*: median age gap of randomly initialised copies of the same backbones, probed identically. *Classical*: age gap of logistic regression on standard ECG measurements. — = not run. Per-backbone values are in Supplementary Table S11. [a]Primary 3-lead, 10 s recording. [b]Fewer than 20 positive ECGs at <40 or ≥75 years (Supplementary Table S6); shown for completeness and excluded from all age-gap aggregates. [c]Patient-grouped subsample of 1,197,523 ECGs.

| Endpoint | Cohort | Label | $n$ ECGs | Overall | Age gap, pretrained | Untrained | Classical |
|---|---|---|---|---|---|---|---|
| Atrial fibrillation | CODE | ECG-read | 2,322,465 | 0.974 (0.971–0.981) | −0.010 (−0.025 to 0.016) | — | — |
| Atrial fibrillation | CODE-15% | ECG-read | 345,779 | 0.987 (0.971–0.991) | −0.009 (−0.043 to 0.002) | 0.066 | −0.045 |
| Atrial fibrillation | CODE-II[b] | ECG-read | 15,000 | 0.962 (0.894–0.980) | 0.040 (−0.081 to 0.064) | — | — |
| Atrial fibrillation | Shaoxing[b] | ECG-read | 45,033 | 0.930 (0.911–0.933) | 0.170 (0.164 to 0.198) | 0.206 | 0.182 |
| Atrial fibrillation | CPSC2018[b] | ECG-read | 6,877 | 0.980 (0.963–0.982) | −0.125 (−0.200 to −0.043) | −0.064 | −0.114 |
| Myocardial infarction | PTB-XL | ECG-read | 21,799 | 0.914 (0.878–0.922) | 0.072 (0.058 to 0.076) | 0.087 | 0.054 |
| LVEF ≤45% | EchoNext | Measured | 100,000 | 0.862 (0.824–0.879) | −0.016 (−0.024 to −0.012) | −0.021 | 0.021 |
| Structural heart disease | EchoNext | Measured | 100,000 | 0.814 (0.779–0.829) | 0.045 (0.041 to 0.054) | 0.042 | 0.063 |
| 30-day MACE/troponin | ED-BWH[a] | Outcome | 61,680 | 0.800 (0.752–0.825) | 0.184 (0.164 to 0.193) | 0.168 | 0.170 |
| MACE | HEEDB-Emory | Outcome | 981,767 | 0.757 (0.722–0.768) | 0.176 (0.149 to 0.184) | 0.171 | 0.156 |
| Myocardial infarction (ICD) | HEEDB-Emory | Coded | 981,767 | 0.744 (0.690–0.758) | 0.109 (0.102 to 0.124) | 0.113 | 0.131 |
| Heart failure (ICD) | HEEDB-Emory | Coded | 981,767 | 0.800 (0.764–0.811) | 0.142 (0.132 to 0.153) | 0.151 | 0.135 |
| MACE | LEAD[b] | Outcome | 14,689 | 0.770 (0.704–0.797) | −0.015 (−0.114 to 0.103) | −0.004 | — |
| Heart failure | LEAD[b] | Coded | 14,689 | 0.807 (0.610–0.829) | 0.320 (0.177 to 0.375) | 0.427 | 0.471 |
| 1-year mortality | HEEDB-MGH[c] | Outcome | 1,197,523 | 0.688 (0.659–0.692) | 0.166 (0.120 to 0.174) | — | — |
| 1-year mortality | CODE-15% | Outcome | 233,647 | 0.817 (0.764–0.829) | 0.089 (0.069 to 0.104) | 0.049 | 0.082 |
| Hyperkalaemia | MIMIC-IV | Measured | 499,280 | 0.711 (0.653–0.753) | 0.030 (0.018 to 0.041) | 0.052 | −0.030 |
| Hypokalaemia | MIMIC-IV | Measured | 499,280 | 0.733 (0.682–0.777) | −0.016 (−0.035 to 0.005) | 0.033 | 0.028 |
| Hypocalcaemia | MIMIC-IV | Measured | 260,675 | 0.724 (0.671–0.753) | 0.060 (0.046 to 0.084) | 0.049 | 0.053 |
| Hypercalcaemia | MIMIC-IV | Measured | 260,675 | 0.690 (0.629–0.720) | 0.001 (−0.023 to 0.018) | −0.008 | −0.036 |
| Elevated NT-proBNP | MIMIC-IV | Measured | 41,350 | 0.871 (0.812–0.891) | 0.040 (0.028 to 0.043) | 0.033 | 0.025 |
| Serum potassium (MAE, mmol/L) | MIMIC-IV | Measured | 499,280 | 0.457 (0.443–0.479) | 0.048 (0.034 to 0.051) | 0.057 | — |
| Serum calcium (MAE, mg/dL) | MIMIC-IV | Measured | 260,675 | 0.564 (0.545–0.587) | −0.028 (−0.043 to −0.013) | −0.042 | — |
| log NT-proBNP (MAE) | MIMIC-IV | Measured | 41,350 | 1.029 (0.949–1.194) | −0.239 (−0.441 to −0.211) | −0.445 | — |

**Table S9: Errors by age band at one pooled operating point.** Four-backbone ensemble (mean predicted probability of ECGFounder, xECG, ECG-FM and DeepECG-SSL; ECGFounder, xECG and ECG-FM for EchoNext), threshold set to give 90% sensitivity in the whole cohort and applied unchanged to every age band. Miss rate = 1 − sensitivity.

| Endpoint | Age band | *n* | Prevalence (%) | Sensitivity | Miss rate (%) | Specificity | PPV | NPV |
|---|---|---|---|---|---|---|---|---|
| MACE, HEEDB-Emory | <40 | 104,149 | 36.3 | 0.80 | 19.5 | 0.71 | 0.61 | 0.864 |
| | ≥75 | 205,701 | 67.1 | 0.97 | 2.9 | 0.08 | 0.68 | 0.575 |
| MI (ICD), HEEDB-Emory | <40 | 104,149 | 6.3 | 0.78 | 21.9 | 0.67 | 0.14 | 0.978 |
| | ≥75 | 205,701 | 20.2 | 0.94 | 5.5 | 0.16 | 0.22 | 0.918 |
| Heart failure (ICD), HEEDB-Emory | <40 | 104,149 | 28.1 | 0.84 | 16.3 | 0.73 | 0.54 | 0.919 |
| | ≥75 | 205,701 | 51.1 | 0.95 | 4.9 | 0.19 | 0.55 | 0.791 |
| 30-d MACE, ED-BWH (3-lead) | <40 | 16,723 | 2.3 | 0.86 | 14.1 | 0.83 | 0.11 | 0.996 |
| | ≥75 | 3,899 | 13.2 | 0.92 | 7.6 | 0.25 | 0.16 | 0.957 |
| MI, PTB-XL | <40 | 2,847 | 4.6 | 0.77 | 23.1 | 0.97 | 0.56 | 0.989 |
| | ≥75 | 4,591 | 35.3 | 0.92 | 8.1 | 0.66 | 0.59 | 0.937 |
| LVEF ≤45%, EchoNext | <40 | 11,433 | 16.6 | 0.81 | 18.7 | 0.72 | 0.36 | 0.951 |
| | ≥75 | 21,372 | 28.7 | 0.93 | 7.0 | 0.55 | 0.46 | 0.952 |
| Structural HD, EchoNext | <40 | 11,433 | 39.0 | 0.84 | 16.3 | 0.66 | 0.62 | 0.865 |
| | ≥75 | 21,372 | 65.4 | 0.94 | 6.5 | 0.33 | 0.72 | 0.727 |

**Table S10: Data sources, access and licences.** All data were de-identified by the providers.

| Cohort | Reference | Source and access | Licence or agreement |
|---|---|---|---|
| HEEDB-MGH, HEEDB-Emory | [S13] | Harvard-Emory ECG Database, Brain Data Science Platform (credentialed) | BDSP data-use agreement |
| MIMIC-IV-ECG | [S14, S15] | PhysioNet (credentialed access) | PhysioNet Credentialed Health Data License 1.5.0 |
| EchoNext | [S16] | PhysioNet (restricted access; registered users under a data-use agreement) | PhysioNet Restricted Health Data License 1.5.0 |
| ED-BWH | — | Nightingale Open Science (registered access) | Nightingale data-use agreement |
| CODE | [S17] | Telehealth Network of Minas Gerais, on request | Data-use agreement |
| CODE-15% (subset of CODE) | [S18] | Zenodo (open) | CC BY 4.0 |
| CODE-II | [S17] | Telehealth Network of Minas Gerais, on request | Data-use agreement |
| PTB-XL | [S19] | PhysioNet (open) | CC BY 4.0 |
| Shaoxing (Chapman–Shaoxing) | [S20] | PhysioNet / figshare (open) | CC BY 4.0 |
| CPSC2018 | [S21] | China Physiological Signal Challenge; PhysioNet/CinC Challenge 2020 (open) | CC BY 4.0 |
| SPHDB | [S22] | Shandong Provincial Hospital database, figshare (open) | CC BY 4.0 |
| LEAD | [S23] | LEAD study, Ludwig Boltzmann Institute for Lung Health, Vienna (collaboration) | Prospective population-based registry (NCT01727518); Ethics Committee of the City of Vienna, EK-11-117-0711 |
| IKEM | [S24] | Institute for Clinical and Experimental Medicine, Prague; Zenodo record 8393007 (open) | CC BY 4.0 |

**Table S11: All pretrained frozen-probe results.** AUROC or MAE overall and in the youngest (<40 years) and oldest (≥75 years) bands; age gap as in Table S8 (positive = oldest worse); sex gap = |female−male|; race gap = best minus worst race/ethnicity group (US cohorts only). One row per pretrained endpoint–backbone result.

| Cohort | Endpoint | Backbone | Metric | Overall | <40 | ≥75 | Age gap | Sex gap | Race gap |
|---|---|---|---|---|---|---|---|---|---|
| CODE-15% | Atrial fibrillation | ECGFounder | AUROC | 0.991 | 0.981 | 0.979 | 0.002 | 0.003 | — |
| CODE-15% | Atrial fibrillation | xECG | AUROC | 0.984 | 0.958 | 0.969 | −0.011 | 0.000 | — |
| CODE-15% | Atrial fibrillation | ECG-FM | AUROC | 0.987 | 0.933 | 0.976 | −0.043 | 0.002 | — |
| CODE-15% | Atrial fibrillation | DeepECG-SSL | AUROC | 0.990 | 0.977 | 0.978 | −0.001 | 0.003 | — |
| CODE-15% | Atrial fibrillation | TiRex-2 | AUROC | 0.971 | 0.930 | 0.939 | −0.009 | 0.004 | — |
| CODE-15% | 1-y mortality | ECGFounder | AUROC | 0.813 | 0.730 | 0.642 | 0.089 | 0.020 | — |
| CODE-15% | 1-y mortality | xECG | AUROC | 0.817 | 0.739 | 0.639 | 0.100 | 0.014 | — |
| CODE-15% | 1-y mortality | ECG-FM | AUROC | 0.819 | 0.733 | 0.651 | 0.082 | 0.025 | — |
| CODE-15% | 1-y mortality | DeepECG-SSL | AUROC | 0.829 | 0.759 | 0.655 | 0.104 | 0.023 | — |
| CODE-15% | 1-y mortality | TiRex-2 | AUROC | 0.764 | 0.681 | 0.612 | 0.069 | 0.002 | — |
| CODE-15% | First-degree AV block | ECGFounder | AUROC | 0.978 | 0.992 | 0.960 | 0.032 | 0.007 | — |
| CODE-15% | First-degree AV block | xECG | AUROC | 0.973 | 0.989 | 0.950 | 0.039 | 0.004 | — |
| CODE-15% | First-degree AV block | ECG-FM | AUROC | 0.983 | 0.992 | 0.965 | 0.027 | 0.006 | — |
| CODE-15% | AF (diagnostic statement) | ECGFounder | AUROC | 0.991 | 0.981 | 0.979 | 0.002 | 0.003 | — |
| CODE-15% | AF (diagnostic statement) | xECG | AUROC | 0.984 | 0.958 | 0.969 | −0.011 | 0.000 | — |
| CODE-15% | AF (diagnostic statement) | ECG-FM | AUROC | 0.987 | 0.933 | 0.976 | −0.043 | 0.002 | — |
| CODE-15% | LBBB | ECGFounder | AUROC | 0.995 | 0.989 | 0.987 | 0.002 | 0.005 | — |
| CODE-15% | LBBB | xECG | AUROC | 0.992 | 0.985 | 0.981 | 0.004 | 0.005 | — |
| CODE-15% | LBBB | ECG-FM | AUROC | 0.995 | 0.997 | 0.987 | 0.010 | 0.004 | — |
| CODE-15% | RBBB | ECGFounder | AUROC | 0.993 | 0.994 | 0.987 | 0.008 | 0.005 | — |
| CODE-15% | RBBB | xECG | AUROC | 0.989 | 0.988 | 0.982 | 0.006 | 0.007 | — |

| Cohort | Endpoint | Backbone | Metric | Overall | <40 | ≥75 | Age gap | Sex gap | Race gap |
|---|---|---|---|---|---|---|---|---|---|
| CODE-15% | RBBB | ECG-FM | AUROC | 0.993 | 0.993 | 0.986 | 0.007 | 0.006 | — |
| CODE-15% | Sinus bradycardia | ECGFounder | AUROC | 0.990 | 0.993 | 0.988 | 0.004 | 0.008 | — |
| CODE-15% | Sinus bradycardia | xECG | AUROC | 0.984 | 0.989 | 0.981 | 0.008 | 0.006 | — |
| CODE-15% | Sinus bradycardia | ECG-FM | AUROC | 0.988 | 0.991 | 0.986 | 0.005 | 0.009 | — |
| CODE-15% | Sinus tachycardia | ECGFounder | AUROC | 0.993 | 0.995 | 0.987 | 0.008 | 0.000 | — |
| CODE-15% | Sinus tachycardia | xECG | AUROC | 0.990 | 0.993 | 0.980 | 0.013 | 0.000 | — |
| CODE-15% | Sinus tachycardia | ECG-FM | AUROC | 0.991 | 0.993 | 0.985 | 0.009 | 0.001 | — |
| CODE | Atrial fibrillation | ECGFounder | AUROC | 0.981 | 0.962 | 0.946 | 0.016 | 0.006 | — |
| CODE | Atrial fibrillation | xECG | AUROC | 0.974 | 0.908 | 0.933 | −0.025 | 0.003 | — |
| CODE | Atrial fibrillation | ECG-FM | AUROC | 0.973 | 0.914 | 0.927 | −0.012 | 0.005 | — |
| CODE | Atrial fibrillation | DeepECG-SSL | AUROC | 0.979 | 0.927 | 0.937 | −0.010 | 0.006 | — |
| CODE | Atrial fibrillation | TiRex-2 | AUROC | 0.971 | 0.940 | 0.931 | 0.009 | 0.005 | — |
| CODE | First-degree AV block | ECGFounder | AUROC | 0.969 | 0.986 | 0.927 | 0.059 | 0.012 | — |
| CODE | First-degree AV block | xECG | AUROC | 0.960 | 0.977 | 0.916 | 0.060 | 0.009 | — |
| CODE | First-degree AV block | ECG-FM | AUROC | 0.963 | 0.981 | 0.919 | 0.063 | 0.010 | — |
| CODE | First-degree AV block | DeepECG-SSL | AUROC | 0.965 | 0.984 | 0.922 | 0.061 | 0.012 | — |
| CODE | First-degree AV block | TiRex-2 | AUROC | 0.943 | 0.966 | 0.886 | 0.080 | 0.016 | — |
| CODE | AF (diagnostic statement) | ECGFounder | AUROC | 0.981 | 0.962 | 0.946 | 0.016 | 0.006 | — |
| CODE | AF (diagnostic statement) | xECG | AUROC | 0.974 | 0.908 | 0.933 | −0.025 | 0.003 | — |
| CODE | AF (diagnostic statement) | ECG-FM | AUROC | 0.973 | 0.914 | 0.927 | −0.012 | 0.005 | — |
| CODE | AF (diagnostic statement) | DeepECG-SSL | AUROC | 0.979 | 0.927 | 0.937 | −0.010 | 0.006 | — |
| CODE | AF (diagnostic statement) | TiRex-2 | AUROC | 0.971 | 0.940 | 0.931 | 0.009 | 0.005 | — |
| CODE | LBBB | ECGFounder | AUROC | 0.987 | 0.993 | 0.965 | 0.028 | — | — |
| CODE | LBBB | xECG | AUROC | 0.984 | 0.985 | 0.957 | 0.029 | — | — |
| CODE | LBBB | ECG-FM | AUROC | 0.986 | 0.987 | 0.961 | 0.026 | — | — |
| CODE | LBBB | DeepECG-SSL | AUROC | 0.988 | 0.993 | 0.965 | 0.028 | — | — |
| CODE | LBBB | TiRex-2 | AUROC | 0.980 | 0.985 | 0.957 | 0.028 | — | — |
| CODE | RBBB | ECGFounder | AUROC | 0.986 | 0.989 | 0.972 | 0.017 | — | — |
| CODE | RBBB | xECG | AUROC | 0.979 | 0.977 | 0.963 | 0.014 | — | — |
| CODE | RBBB | ECG-FM | AUROC | 0.982 | 0.984 | 0.964 | 0.020 | — | — |
| CODE | RBBB | DeepECG-SSL | AUROC | 0.984 | 0.986 | 0.967 | 0.019 | — | — |
| CODE | RBBB | TiRex-2 | AUROC | 0.985 | 0.981 | 0.970 | 0.011 | — | — |
| CODE | Sinus bradycardia | ECGFounder | AUROC | 0.980 | 0.985 | 0.978 | 0.007 | 0.013 | — |
| CODE | Sinus bradycardia | xECG | AUROC | 0.974 | 0.981 | 0.973 | 0.009 | 0.009 | — |
| CODE | Sinus bradycardia | ECG-FM | AUROC | 0.972 | 0.978 | 0.968 | 0.010 | 0.017 | — |
| CODE | Sinus bradycardia | DeepECG-SSL | AUROC | 0.977 | 0.983 | 0.973 | 0.011 | 0.016 | — |
| CODE | Sinus bradycardia | TiRex-2 | AUROC | 0.976 | 0.983 | 0.969 | 0.013 | 0.012 | — |
| CODE | Sinus tachycardia | ECGFounder | AUROC | 0.988 | 0.991 | 0.979 | 0.012 | — | — |
| CODE | Sinus tachycardia | xECG | AUROC | 0.983 | 0.987 | 0.969 | 0.018 | — | — |
| CODE | Sinus tachycardia | ECG-FM | AUROC | 0.982 | 0.986 | 0.969 | 0.017 | — | — |
| CODE | Sinus tachycardia | DeepECG-SSL | AUROC | 0.985 | 0.988 | 0.972 | 0.016 | — | — |
| CODE | Sinus tachycardia | TiRex-2 | AUROC | 0.982 | 0.986 | 0.969 | 0.017 | — | — |
| CODE-II | Atrial fibrillation | ECGFounder | AUROC | 0.947 | 0.860 | 0.941 | −0.081 | 0.003 | — |
| CODE-II | Atrial fibrillation | xECG | AUROC | 0.973 | 0.984 | 0.935 | 0.048 | 0.004 | — |
| CODE-II | Atrial fibrillation | ECG-FM | AUROC | 0.962 | 0.987 | 0.947 | 0.040 | 0.015 | — |
| CODE-II | Atrial fibrillation | DeepECG-SSL | AUROC | 0.980 | 0.995 | 0.961 | 0.034 | 0.006 | — |
| CODE-II | Atrial fibrillation | TiRex-2 | AUROC | 0.894 | 0.865 | 0.801 | 0.064 | 0.019 | — |
| CODE-II | AF (diagnostic statement) | ECGFounder | AUROC | 0.947 | 0.860 | 0.941 | −0.081 | 0.003 | — |
| CODE-II | AF (diagnostic statement) | xECG | AUROC | 0.973 | 0.984 | 0.935 | 0.048 | 0.004 | — |
| CODE-II | AF (diagnostic statement) | ECG-FM | AUROC | 0.962 | 0.987 | 0.947 | 0.040 | 0.015 | — |
| CODE-II | AF (diagnostic statement) | DeepECG-SSL | AUROC | 0.980 | 0.995 | 0.961 | 0.034 | 0.006 | — |
| CODE-II | AF (diagnostic statement) | TiRex-2 | AUROC | 0.894 | 0.865 | 0.801 | 0.064 | 0.019 | — |
| CPSC2018 | Atrial fibrillation | ECGFounder | AUROC | 0.980 | 0.926 | 0.969 | −0.043 | 0.004 | — |
| CPSC2018 | Atrial fibrillation | xECG | AUROC | 0.982 | 0.848 | 0.973 | −0.125 | 0.005 | — |
| CPSC2018 | Atrial fibrillation | ECG-FM | AUROC | 0.975 | 0.764 | 0.964 | −0.200 | 0.008 | — |
| CPSC2018 | Atrial fibrillation | DeepECG-SSL | AUROC | 0.980 | 0.815 | 0.974 | −0.159 | 0.003 | — |
| CPSC2018 | Atrial fibrillation | TiRex-2 | AUROC | 0.963 | 0.866 | 0.937 | −0.071 | 0.009 | — |
| CPSC2018 | AF (SNOMED) | ECGFounder | AUROC | 0.987 | 0.875 | 0.978 | −0.103 | 0.001 | — |
| CPSC2018 | AF (SNOMED) | xECG | AUROC | 0.987 | 0.834 | 0.979 | −0.145 | 0.004 | — |
| CPSC2018 | AF (SNOMED) | ECG-FM | AUROC | 0.986 | 0.813 | 0.978 | −0.165 | 0.008 | — |
| CPSC2018 | RBBB (SNOMED) | ECGFounder | AUROC | 0.980 | 0.981 | 0.980 | 0.001 | 0.007 | — |
| CPSC2018 | RBBB (SNOMED) | xECG | AUROC | 0.985 | 0.987 | 0.979 | 0.007 | 0.009 | — |
| CPSC2018 | RBBB (SNOMED) | ECG-FM | AUROC | 0.972 | 0.948 | 0.976 | −0.028 | 0.002 | — |
| EchoNext | LVEF ≤45% | ECGFounder | AUROC | 0.853 | 0.821 | 0.838 | −0.017 | 0.010 | 0.024 |
| EchoNext | LVEF ≤45% | xECG | AUROC | 0.862 | 0.839 | 0.851 | −0.012 | 0.009 | 0.022 |
| EchoNext | LVEF ≤45% | ECG-FM | AUROC | 0.868 | 0.843 | 0.859 | −0.016 | 0.006 | 0.024 |

| Cohort | Endpoint | Backbone | Metric | Overall | <40 | ≥75 | Age gap | Sex gap | Race gap |
|---|---|---|---|---|---|---|---|---|---|
| EchoNext | LVEF ≤45% | DeepECG-SSL | AUROC | 0.879 | 0.851 | 0.874 | −0.024 | 0.002 | 0.023 |
| EchoNext | LVEF ≤45% | TiRex-2 | AUROC | 0.824 | 0.796 | 0.810 | −0.014 | 0.009 | 0.022 |
| EchoNext | Structural heart disease | ECGFounder | AUROC | 0.811 | 0.837 | 0.783 | 0.054 | 0.003 | 0.036 |
| EchoNext | Structural heart disease | xECG | AUROC | 0.814 | 0.835 | 0.789 | 0.045 | 0.003 | 0.041 |
| EchoNext | Structural heart disease | ECG-FM | AUROC | 0.819 | 0.839 | 0.793 | 0.046 | 0.001 | 0.039 |
| EchoNext | Structural heart disease | DeepECG-SSL | AUROC | 0.829 | 0.846 | 0.805 | 0.041 | 0.003 | 0.040 |
| EchoNext | Structural heart disease | TiRex-2 | AUROC | 0.779 | 0.796 | 0.750 | 0.045 | 0.008 | 0.047 |
| ED-BWH (12-lead) | 30-d MACE/troponin | ECGFounder | AUROC | 0.743 | 0.827 | 0.634 | 0.193 | 0.003 | 0.056 |
| ED-BWH (12-lead) | 30-d MACE/troponin | xECG | AUROC | 0.740 | 0.816 | 0.655 | 0.160 | 0.015 | 0.035 |
| ED-BWH (12-lead) | 30-d MACE/troponin | ECG-FM | AUROC | 0.751 | 0.844 | 0.647 | 0.197 | 0.022 | 0.056 |
| ED-BWH (12-lead) | 30-d MACE/troponin | DeepECG-SSL | AUROC | 0.767 | 0.852 | 0.646 | 0.206 | 0.010 | 0.041 |
| ED-BWH (12-lead) | 30-d MACE/troponin | TiRex-2 | AUROC | 0.768 | 0.834 | 0.665 | 0.168 | 0.012 | 0.061 |
| ED-BWH (3-lead) | 30-d MACE/troponin | ECGFounder | AUROC | 0.752 | 0.809 | 0.645 | 0.164 | 0.017 | 0.050 |
| ED-BWH (3-lead) | 30-d MACE/troponin | xECG | AUROC | 0.800 | 0.870 | 0.687 | 0.182 | 0.013 | 0.051 |
| ED-BWH (3-lead) | 30-d MACE/troponin | ECG-FM | AUROC | 0.810 | 0.887 | 0.703 | 0.184 | 0.009 | 0.050 |
| ED-BWH (3-lead) | 30-d MACE/troponin | DeepECG-SSL | AUROC | 0.825 | 0.902 | 0.712 | 0.190 | 0.000 | 0.060 |
| ED-BWH (3-lead) | 30-d MACE/troponin | TiRex-2 | AUROC | 0.767 | 0.858 | 0.665 | 0.193 | 0.002 | 0.038 |
| HEEDB-Emory | Mortality | ECGFounder | AUROC | 0.739 | 0.800 | 0.640 | 0.159 | 0.019 | 0.049 |
| HEEDB-Emory | Mortality | xECG | AUROC | 0.753 | 0.811 | 0.650 | 0.161 | 0.021 | 0.042 |
| HEEDB-Emory | Mortality | ECG-FM | AUROC | 0.745 | 0.800 | 0.646 | 0.154 | 0.012 | 0.062 |
| HEEDB-Emory | Mortality | TiRex-2 | AUROC | 0.712 | 0.761 | 0.628 | 0.133 | 0.006 | 0.064 |
| HEEDB-Emory | Heart failure (ICD) | ECGFounder | AUROC | 0.792 | 0.849 | 0.696 | 0.153 | 0.002 | 0.036 |
| HEEDB-Emory | Heart failure (ICD) | xECG | AUROC | 0.807 | 0.860 | 0.715 | 0.145 | 0.001 | 0.035 |
| HEEDB-Emory | Heart failure (ICD) | ECG-FM | AUROC | 0.800 | 0.852 | 0.710 | 0.142 | 0.002 | 0.039 |
| HEEDB-Emory | Heart failure (ICD) | DeepECG-SSL | AUROC | 0.811 | 0.862 | 0.722 | 0.140 | 0.001 | — |
| HEEDB-Emory | Heart failure (ICD) | TiRex-2 | AUROC | 0.764 | 0.811 | 0.679 | 0.132 | 0.011 | 0.034 |
| HEEDB-Emory | MACE | ECGFounder | AUROC | 0.751 | 0.815 | 0.630 | 0.184 | 0.008 | 0.062 |
| HEEDB-Emory | MACE | xECG | AUROC | 0.764 | 0.828 | 0.647 | 0.181 | 0.009 | 0.058 |
| HEEDB-Emory | MACE | ECG-FM | AUROC | 0.757 | 0.818 | 0.642 | 0.176 | 0.004 | 0.058 |
| HEEDB-Emory | MACE | DeepECG-SSL | AUROC | 0.768 | 0.828 | 0.654 | 0.174 | 0.006 | — |
| HEEDB-Emory | MACE | TiRex-2 | AUROC | 0.722 | 0.774 | 0.625 | 0.149 | 0.006 | 0.055 |
| HEEDB-Emory | MI (ICD) | ECGFounder | AUROC | 0.729 | 0.767 | 0.643 | 0.124 | 0.027 | 0.078 |
| HEEDB-Emory | MI (ICD) | xECG | AUROC | 0.746 | 0.778 | 0.659 | 0.119 | 0.024 | 0.048 |
| HEEDB-Emory | MI (ICD) | ECG-FM | AUROC | 0.744 | 0.771 | 0.662 | 0.109 | 0.024 | 0.046 |
| HEEDB-Emory | MI (ICD) | DeepECG-SSL | AUROC | 0.758 | 0.786 | 0.676 | 0.109 | 0.021 | — |
| HEEDB-Emory | MI (ICD) | TiRex-2 | AUROC | 0.690 | 0.715 | 0.613 | 0.102 | 0.021 | 0.065 |
| HEEDB-Emory | AF (12SL statement) | ECGFounder | AUROC | 0.969 | 0.946 | 0.963 | −0.017 | 0.007 | 0.013 |
| HEEDB-Emory | AF (12SL statement) | xECG | AUROC | 0.969 | 0.945 | 0.962 | −0.018 | 0.007 | 0.013 |
| HEEDB-Emory | AF (12SL statement) | ECG-FM | AUROC | 0.968 | 0.936 | 0.962 | −0.026 | 0.007 | 0.021 |
| HEEDB-Emory | AF (12SL statement) | TiRex-2 | AUROC | 0.926 | 0.895 | 0.899 | −0.004 | 0.015 | 0.023 |
| HEEDB-Emory | Anterior MI (12SL) | ECGFounder | AUROC | 0.932 | 0.940 | 0.913 | 0.027 | 0.007 | 0.026 |
| HEEDB-Emory | Anterior MI (12SL) | xECG | AUROC | 0.900 | 0.917 | 0.876 | 0.042 | 0.013 | 0.041 |
| HEEDB-Emory | Anterior MI (12SL) | ECG-FM | AUROC | 0.844 | 0.854 | 0.826 | 0.029 | 0.021 | 0.052 |
| HEEDB-Emory | Anterior MI (12SL) | TiRex-2 | AUROC | 0.829 | 0.846 | 0.810 | 0.036 | 0.023 | 0.051 |
| HEEDB-Emory | Inferior MI (12SL) | ECGFounder | AUROC | 0.952 | 0.951 | 0.943 | 0.008 | 0.004 | 0.020 |
| HEEDB-Emory | Inferior MI (12SL) | xECG | AUROC | 0.943 | 0.942 | 0.931 | 0.011 | 0.005 | 0.021 |
| HEEDB-Emory | Inferior MI (12SL) | ECG-FM | AUROC | 0.913 | 0.916 | 0.898 | 0.018 | 0.005 | 0.027 |
| HEEDB-Emory | Inferior MI (12SL) | TiRex-2 | AUROC | 0.798 | 0.840 | 0.755 | 0.085 | 0.020 | 0.027 |
| HEEDB-Emory | RBBB (12SL) | ECGFounder | AUROC | 0.986 | 0.988 | 0.980 | 0.008 | 0.005 | 0.004 |
| HEEDB-Emory | RBBB (12SL) | xECG | AUROC | 0.984 | 0.986 | 0.977 | 0.009 | 0.005 | 0.005 |
| HEEDB-Emory | RBBB (12SL) | ECG-FM | AUROC | 0.985 | 0.988 | 0.979 | 0.009 | 0.006 | 0.005 |
| HEEDB-Emory | RBBB (12SL) | TiRex-2 | AUROC | 0.981 | 0.987 | 0.973 | 0.014 | 0.007 | 0.011 |
| IKEM | Tachycardia | ECGFounder | AUROC | 0.995 | 0.996 | 0.993 | 0.003 | 0.005 | — |
| IKEM | Tachycardia | xECG | AUROC | 0.994 | 0.995 | 0.991 | 0.004 | 0.006 | — |
| IKEM | Tachycardia | ECG-FM | AUROC | 0.992 | 0.994 | 0.986 | 0.008 | 0.006 | — |
| IKEM | Tachycardia | DeepECG-SSL | AUROC | 0.994 | 0.996 | 0.989 | 0.007 | 0.004 | — |
| LEAD | Heart failure | ECGFounder | AUROC | 0.610 | 0.764 | 0.587 | 0.177 | 0.057 | — |
| LEAD | Heart failure | xECG | AUROC | 0.812 | 0.999 | 0.679 | 0.320 | 0.049 | — |
| LEAD | Heart failure | ECG-FM | AUROC | 0.829 | 0.989 | 0.665 | 0.324 | 0.019 | — |
| LEAD | Heart failure | DeepECG-SSL | AUROC | 0.807 | 0.953 | 0.652 | 0.300 | 0.050 | — |
| LEAD | Heart failure | TiRex-2 | AUROC | 0.771 | 0.889 | 0.514 | 0.375 | 0.045 | — |
| LEAD | MACE | ECGFounder | AUROC | 0.704 | 0.547 | 0.523 | 0.024 | 0.042 | — |
| LEAD | MACE | xECG | AUROC | 0.797 | 0.727 | 0.625 | 0.103 | 0.028 | — |
| LEAD | MACE | ECG-FM | AUROC | 0.783 | 0.558 | 0.672 | −0.114 | 0.064 | — |
| LEAD | MACE | DeepECG-SSL | AUROC | 0.770 | 0.535 | 0.608 | −0.073 | 0.062 | — |
| LEAD | MACE | TiRex-2 | AUROC | 0.769 | 0.612 | 0.627 | −0.015 | 0.036 | — |

| Cohort | Endpoint | Backbone | Metric | Overall | <40 | ≥75 | Age gap | Sex gap | Race gap |
|---|---|---|---|---|---|---|---|---|---|
| LEAD | MI | ECGFounder | AUROC | 0.679 | 0.429 | 0.513 | −0.084 | 0.143 | — |
| LEAD | MI | xECG | AUROC | 0.763 | 0.617 | 0.633 | −0.016 | 0.068 | — |
| LEAD | MI | ECG-FM | AUROC | 0.750 | 0.564 | 0.581 | −0.017 | 0.081 | — |
| LEAD | MI | DeepECG-SSL | AUROC | 0.754 | 0.535 | 0.654 | −0.119 | 0.065 | — |
| LEAD | MI | TiRex-2 | AUROC | 0.759 | 0.676 | 0.636 | 0.040 | 0.033 | — |
| HEEDB-MGH | Mortality | ECGFounder | AUROC | 0.688 | 0.756 | 0.594 | 0.162 | 0.229 | 0.089 |
| HEEDB-MGH | Mortality | xECG | AUROC | 0.692 | 0.774 | 0.600 | 0.174 | 0.094 | 0.063 |
| HEEDB-MGH | Mortality | ECG-FM | AUROC | 0.677 | 0.758 | 0.592 | 0.166 | 0.020 | 0.049 |
| HEEDB-MGH | Mortality | DeepECG-SSL | AUROC | 0.691 | 0.772 | 0.601 | 0.171 | 0.209 | 0.058 |
| HEEDB-MGH | Mortality | TiRex-2 | AUROC | 0.659 | 0.701 | 0.581 | 0.120 | 0.150 | 0.069 |
| MIMIC-IV | Calcium (MAE) | ECGFounder | MAE | 0.574 | 0.581 | 0.561 | −0.021 | 0.018 | 0.034 |
| MIMIC-IV | Calcium (MAE) | xECG | MAE | 0.545 | 0.545 | 0.532 | −0.013 | 0.014 | 0.040 |
| MIMIC-IV | Calcium (MAE) | ECG-FM | MAE | 0.564 | 0.577 | 0.548 | −0.028 | 0.018 | 0.043 |
| MIMIC-IV | Calcium (MAE) | DeepECG-SSL | MAE | 0.559 | 0.573 | 0.544 | −0.029 | 0.019 | 0.036 |
| MIMIC-IV | Calcium (MAE) | TiRex-2 | MAE | 0.587 | 0.610 | 0.567 | −0.043 | 0.023 | 0.050 |
| MIMIC-IV | 1-y mortality | ECGFounder | AUROC | 0.749 | 0.781 | 0.653 | 0.128 | 0.022 | 0.161 |
| MIMIC-IV | 1-y mortality | xECG | AUROC | 0.764 | 0.805 | 0.669 | 0.137 | 0.020 | 0.163 |
| MIMIC-IV | 1-y mortality | ECG-FM | AUROC | 0.728 | 0.769 | 0.633 | 0.135 | 0.015 | 0.160 |
| MIMIC-IV | 1-y mortality | DeepECG-SSL | AUROC | 0.743 | 0.791 | 0.646 | 0.145 | 0.020 | 0.162 |
| MIMIC-IV | 1-y mortality | TiRex-2 | AUROC | 0.701 | 0.720 | 0.616 | 0.104 | 0.005 | 0.160 |
| MIMIC-IV | Hypercalcaemia | ECGFounder | AUROC | 0.658 | 0.645 | 0.652 | −0.008 | 0.008 | 0.093 |
| MIMIC-IV | Hypercalcaemia | xECG | AUROC | 0.718 | 0.731 | 0.713 | 0.018 | 0.001 | 0.071 |
| MIMIC-IV | Hypercalcaemia | ECG-FM | AUROC | 0.690 | 0.692 | 0.691 | 0.001 | 0.005 | 0.109 |
| MIMIC-IV | Hypercalcaemia | DeepECG-SSL | AUROC | 0.720 | 0.696 | 0.719 | −0.023 | 0.010 | 0.052 |
| MIMIC-IV | Hypercalcaemia | TiRex-2 | AUROC | 0.629 | 0.637 | 0.629 | 0.008 | 0.008 | 0.093 |
| MIMIC-IV | Hyperkalaemia | ECGFounder | AUROC | 0.711 | 0.722 | 0.681 | 0.041 | 0.015 | 0.072 |
| MIMIC-IV | Hyperkalaemia | xECG | AUROC | 0.753 | 0.753 | 0.732 | 0.021 | 0.022 | 0.059 |
| MIMIC-IV | Hyperkalaemia | ECG-FM | AUROC | 0.686 | 0.700 | 0.670 | 0.030 | 0.016 | 0.060 |
| MIMIC-IV | Hyperkalaemia | DeepECG-SSL | AUROC | 0.720 | 0.725 | 0.707 | 0.018 | 0.019 | 0.079 |
| MIMIC-IV | Hyperkalaemia | TiRex-2 | AUROC | 0.653 | 0.669 | 0.633 | 0.036 | 0.011 | 0.053 |
| MIMIC-IV | Hypocalcaemia | ECGFounder | AUROC | 0.702 | 0.749 | 0.665 | 0.084 | 0.020 | 0.086 |
| MIMIC-IV | Hypocalcaemia | xECG | AUROC | 0.753 | 0.802 | 0.724 | 0.078 | 0.021 | 0.085 |
| MIMIC-IV | Hypocalcaemia | ECG-FM | AUROC | 0.724 | 0.757 | 0.697 | 0.060 | 0.015 | 0.085 |
| MIMIC-IV | Hypocalcaemia | DeepECG-SSL | AUROC | 0.734 | 0.767 | 0.709 | 0.059 | 0.012 | 0.087 |
| MIMIC-IV | Hypocalcaemia | TiRex-2 | AUROC | 0.671 | 0.694 | 0.647 | 0.046 | 0.012 | 0.081 |
| MIMIC-IV | Hypokalaemia | ECGFounder | AUROC | 0.733 | 0.725 | 0.725 | −0.000 | 0.007 | 0.012 |
| MIMIC-IV | Hypokalaemia | xECG | AUROC | 0.777 | 0.761 | 0.777 | −0.016 | 0.009 | 0.022 |
| MIMIC-IV | Hypokalaemia | ECG-FM | AUROC | 0.729 | 0.710 | 0.731 | −0.021 | 0.005 | 0.033 |
| MIMIC-IV | Hypokalaemia | DeepECG-SSL | AUROC | 0.757 | 0.729 | 0.764 | −0.035 | 0.008 | 0.031 |
| MIMIC-IV | Hypokalaemia | TiRex-2 | AUROC | 0.682 | 0.681 | 0.676 | 0.005 | 0.006 | 0.011 |
| MIMIC-IV | Potassium (MAE) | ECGFounder | MAE | 0.471 | 0.430 | 0.480 | 0.050 | 0.010 | 0.105 |
| MIMIC-IV | Potassium (MAE) | xECG | MAE | 0.450 | 0.407 | 0.458 | 0.051 | 0.009 | 0.093 |
| MIMIC-IV | Potassium (MAE) | ECG-FM | MAE | 0.457 | 0.419 | 0.462 | 0.043 | 0.010 | 0.097 |
| MIMIC-IV | Potassium (MAE) | DeepECG-SSL | MAE | 0.443 | 0.402 | 0.450 | 0.048 | 0.009 | 0.093 |
| MIMIC-IV | Potassium (MAE) | TiRex-2 | MAE | 0.479 | 0.451 | 0.485 | 0.034 | 0.013 | 0.097 |
| MIMIC-IV | Elevated NT-proBNP | ECGFounder | AUROC | 0.865 | 0.886 | 0.845 | 0.041 | 0.004 | 0.056 |
| MIMIC-IV | Elevated NT-proBNP | xECG | AUROC | 0.885 | 0.900 | 0.866 | 0.034 | 0.001 | 0.059 |
| MIMIC-IV | Elevated NT-proBNP | ECG-FM | AUROC | 0.871 | 0.896 | 0.853 | 0.043 | 0.005 | 0.063 |
| MIMIC-IV | Elevated NT-proBNP | DeepECG-SSL | AUROC | 0.891 | 0.915 | 0.875 | 0.040 | 0.001 | 0.054 |
| MIMIC-IV | Elevated NT-proBNP | TiRex-2 | AUROC | 0.812 | 0.815 | 0.787 | 0.028 | 0.015 | 0.066 |
| MIMIC-IV | log NT-proBNP (MAE) | ECGFounder | MAE | 1.041 | 1.198 | 0.959 | −0.239 | 0.007 | 0.107 |
| MIMIC-IV | log NT-proBNP (MAE) | xECG | MAE | 0.969 | 1.119 | 0.897 | −0.222 | 0.017 | 0.103 |
| MIMIC-IV | log NT-proBNP (MAE) | ECG-FM | MAE | 1.029 | 1.193 | 0.948 | −0.245 | 0.015 | 0.145 |
| MIMIC-IV | log NT-proBNP (MAE) | DeepECG-SSL | MAE | 0.949 | 1.090 | 0.879 | −0.211 | 0.015 | 0.093 |
| MIMIC-IV | log NT-proBNP (MAE) | TiRex-2 | MAE | 1.194 | 1.518 | 1.077 | −0.441 | 0.051 | 0.243 |
| PTB-XL | Age (MAE, years) | ECGFounder | MAE | 8.494 | 11.452 | 10.743 | −0.710 | 0.204 | — |
| PTB-XL | Age (MAE, years) | xECG | MAE | 7.188 | 8.573 | 9.026 | 0.453 | 0.286 | — |
| PTB-XL | Age (MAE, years) | ECG-FM | MAE | 8.008 | 10.689 | 9.798 | −0.891 | 0.347 | — |
| PTB-XL | Conduction disturbance | ECGFounder | AUROC | 0.927 | 0.933 | 0.914 | 0.019 | 0.003 | — |
| PTB-XL | Conduction disturbance | xECG | AUROC | 0.926 | 0.921 | 0.915 | 0.005 | 0.002 | — |
| PTB-XL | Conduction disturbance | ECG-FM | AUROC | 0.907 | 0.878 | 0.901 | −0.024 | 0.006 | — |
| PTB-XL | Conduction disturbance | DeepECG-SSL | AUROC | 0.916 | 0.886 | 0.912 | −0.026 | 0.006 | — |
| PTB-XL | Conduction disturbance | TiRex-2 | AUROC | 0.868 | 0.834 | 0.852 | −0.018 | 0.017 | — |
| PTB-XL | Hypertrophy | ECGFounder | AUROC | 0.851 | 0.886 | 0.805 | 0.080 | 0.024 | — |
| PTB-XL | Hypertrophy | xECG | AUROC | 0.906 | 0.936 | 0.866 | 0.070 | 0.016 | — |
| PTB-XL | Hypertrophy | ECG-FM | AUROC | 0.838 | 0.845 | 0.798 | 0.047 | 0.015 | — |

| Cohort | Endpoint | Backbone | Metric | Overall | <40 | ≥75 | Age gap | Sex gap | Race gap |
|---|---|---|---|---|---|---|---|---|---|
| PTB-XL | Hypertrophy | DeepECG-SSL | AUROC | 0.863 | 0.898 | 0.809 | 0.089 | 0.013 | — |
| PTB-XL | Hypertrophy | TiRex-2 | AUROC | 0.814 | 0.832 | 0.757 | 0.074 | 0.024 | — |
| PTB-XL | Myocardial infarction | ECGFounder | AUROC | 0.919 | 0.954 | 0.878 | 0.076 | 0.008 | — |
| PTB-XL | Myocardial infarction | xECG | AUROC | 0.922 | 0.934 | 0.876 | 0.058 | 0.005 | — |
| PTB-XL | Myocardial infarction | ECG-FM | AUROC | 0.908 | 0.929 | 0.857 | 0.072 | 0.019 | — |
| PTB-XL | Myocardial infarction | DeepECG-SSL | AUROC | 0.914 | 0.937 | 0.869 | 0.069 | 0.014 | — |
| PTB-XL | Myocardial infarction | TiRex-2 | AUROC | 0.878 | 0.910 | 0.835 | 0.075 | 0.005 | — |
| PTB-XL | Normal ECG | ECGFounder | AUROC | 0.945 | 0.889 | 0.937 | −0.048 | 0.000 | — |
| PTB-XL | Normal ECG | xECG | AUROC | 0.950 | 0.910 | 0.948 | −0.039 | 0.003 | — |
| PTB-XL | Normal ECG | ECG-FM | AUROC | 0.938 | 0.876 | 0.938 | −0.061 | 0.003 | — |
| PTB-XL | ST/T change | ECGFounder | AUROC | 0.916 | 0.949 | 0.874 | 0.075 | 0.001 | — |
| PTB-XL | ST/T change | xECG | AUROC | 0.926 | 0.960 | 0.887 | 0.073 | 0.004 | — |
| PTB-XL | ST/T change | ECG-FM | AUROC | 0.918 | 0.949 | 0.874 | 0.075 | 0.001 | — |
| PTB-XL | Sex (female) | ECGFounder | AUROC | 0.890 | 0.940 | 0.806 | 0.134 | — | — |
| PTB-XL | Sex (female) | xECG | AUROC | 0.936 | 0.974 | 0.877 | 0.097 | — | — |
| PTB-XL | Sex (female) | ECG-FM | AUROC | 0.898 | 0.951 | 0.817 | 0.134 | — | — |
| PTB-XL | Sex (female) | DeepECG-SSL | AUROC | 0.925 | 0.970 | 0.864 | 0.106 | — | — |
| PTB-XL | Sex (female) | TiRex-2 | AUROC | 0.802 | 0.870 | 0.710 | 0.159 | — | — |
| Shaoxing | Atrial fibrillation | ECGFounder | AUROC | 0.930 | 0.995 | 0.825 | 0.170 | 0.003 | — |
| Shaoxing | Atrial fibrillation | xECG | AUROC | 0.933 | 0.996 | 0.832 | 0.164 | 0.005 | — |
| Shaoxing | Atrial fibrillation | ECG-FM | AUROC | 0.931 | 0.995 | 0.830 | 0.165 | 0.001 | — |
| Shaoxing | Atrial fibrillation | DeepECG-SSL | AUROC | 0.929 | 0.994 | 0.822 | 0.172 | 0.005 | — |
| Shaoxing | Atrial fibrillation | TiRex-2 | AUROC | 0.911 | 0.990 | 0.792 | 0.198 | 0.002 | — |
| Shaoxing | AF (SNOMED) | ECGFounder | AUROC | 0.930 | 0.995 | 0.825 | 0.170 | 0.003 | — |
| Shaoxing | AF (SNOMED) | xECG | AUROC | 0.933 | 0.996 | 0.832 | 0.164 | 0.005 | — |
| Shaoxing | AF (SNOMED) | ECG-FM | AUROC | 0.931 | 0.995 | 0.830 | 0.165 | 0.001 | — |
| Shaoxing | RBBB (SNOMED) | ECGFounder | AUROC | 0.952 | 0.897 | 0.926 | −0.030 | 0.028 | — |
| Shaoxing | RBBB (SNOMED) | xECG | AUROC | 0.971 | 0.968 | 0.953 | 0.015 | 0.012 | — |
| Shaoxing | RBBB (SNOMED) | ECG-FM | AUROC | 0.967 | 0.928 | 0.954 | −0.026 | 0.008 | — |
| Shaoxing | Sinus bradycardia | ECGFounder | AUROC | 0.998 | 0.996 | 0.996 | 0.000 | 0.001 | — |
| Shaoxing | Sinus bradycardia | xECG | AUROC | 0.998 | 0.997 | 0.998 | −0.001 | 0.000 | — |
| Shaoxing | Sinus bradycardia | ECG-FM | AUROC | 0.997 | 0.994 | 0.998 | −0.004 | 0.000 | — |
| Shaoxing | Sinus tachycardia | ECGFounder | AUROC | 0.996 | 0.994 | 0.992 | 0.002 | 0.002 | — |
| Shaoxing | Sinus tachycardia | xECG | AUROC | 0.997 | 0.995 | 0.993 | 0.002 | 0.000 | — |
| Shaoxing | Sinus tachycardia | ECG-FM | AUROC | 0.997 | 0.993 | 0.995 | −0.002 | 0.001 | — |
| SPHDB | AF | ECGFounder | AUROC | 0.983 | 0.999 | 0.985 | 0.014 | 0.005 | — |
| SPHDB | AF | xECG | AUROC | 0.993 | 1.000 | 0.995 | 0.004 | 0.003 | — |
| SPHDB | AF | ECG-FM | AUROC | 0.997 | 1.000 | 0.994 | 0.006 | 0.001 | — |
| SPHDB | AF | DeepECG-SSL | AUROC | 0.997 | 1.000 | 0.995 | 0.005 | 0.001 | — |
| SPHDB | LVH | ECGFounder | AUROC | 0.890 | 0.858 | 0.870 | −0.012 | 0.036 | — |
| SPHDB | LVH | xECG | AUROC | 0.975 | 0.977 | 0.970 | 0.008 | 0.004 | — |
| SPHDB | LVH | ECG-FM | AUROC | 0.890 | 0.903 | 0.908 | −0.005 | 0.029 | — |
| SPHDB | LVH | DeepECG-SSL | AUROC | 0.921 | 0.953 | 0.901 | 0.052 | 0.018 | — |
| SPHDB | PVC | ECGFounder | AUROC | 0.893 | 0.877 | 0.928 | −0.051 | 0.018 | — |
| SPHDB | PVC | xECG | AUROC | 0.878 | 0.900 | 0.800 | 0.100 | 0.005 | — |
| SPHDB | PVC | ECG-FM | AUROC | 0.763 | 0.774 | 0.716 | 0.058 | 0.012 | — |
| SPHDB | PVC | DeepECG-SSL | AUROC | 0.883 | 0.906 | 0.802 | 0.104 | 0.013 | — |
| SPHDB | RBBB | ECGFounder | AUROC | 0.990 | 0.985 | 0.997 | −0.012 | 0.006 | — |
| SPHDB | RBBB | xECG | AUROC | 0.993 | 0.973 | 0.994 | −0.021 | 0.009 | — |
| SPHDB | RBBB | ECG-FM | AUROC | 0.991 | 0.977 | 0.996 | −0.019 | 0.012 | — |
| SPHDB | RBBB | DeepECG-SSL | AUROC | 0.992 | 0.982 | 0.994 | −0.012 | 0.011 | — |
| SPHDB | ST deviation | ECGFounder | AUROC | 0.908 | 0.935 | 0.828 | 0.106 | 0.042 | — |
| SPHDB | ST deviation | xECG | AUROC | 0.943 | 0.965 | 0.874 | 0.090 | 0.025 | — |
| SPHDB | ST deviation | ECG-FM | AUROC | 0.938 | 0.961 | 0.875 | 0.085 | 0.038 | — |
| SPHDB | ST deviation | DeepECG-SSL | AUROC | 0.944 | 0.967 | 0.882 | 0.085 | 0.032 | — |

**Table S12: TRIPOD+AI checklist.** Items of the TRIPOD+AI statement [S25] (abridged wording) and where each is reported. The study evaluates released models under a fixed frozen-probe protocol; items that apply only to the development of a model for implementation are marked not applicable.

| Section/topic | Item | Checklist item (abridged) | Where reported |
|---|---|---|---|
| Title | 1 | Identify the study as developing or evaluating a multivariable prediction model, the target population and the outcome | Title page; title names the models evaluated, the population (thirteen cohorts) and the disparity studied |
| Abstract | 2 | TRIPOD+AI for Abstracts | Structured summary (Background, Methods, Findings, Interpretation, Funding) |
| Background | 3a | Healthcare context and rationale, including references to existing models | Introduction, paragraphs 1–2 |
| | 3b | Target population, intended purpose and intended users | Introduction, paragraph 1; Discussion, clinical implications |
| | 3c | Known health inequalities between sociodemographic groups | Introduction, paragraph 2 |
| Objectives | 4 | Study objectives; development or evaluation | Introduction, final paragraph (evaluation of released models under one frozen-probe protocol) |
| Data | 5a | Sources of data, rationale and representativeness | Methods, Cohorts and endpoints; Supplementary Tables S1 and S10; Fig. 1 |
| | 5b | Dates of participant data | Acquisition periods as published for each source dataset (references in Supplementary Table S10); not re-derived |
| Participants | 6a | Study setting, number and location of centres | Methods, Cohorts and endpoints; Fig. 1; Supplementary Table S1 |
| | 6b | Eligibility criteria | Supplementary Methods (Cohorts); Supplementary Figs S1–S3 (curation flows) |
| | 6c | Treatments received | Not applicable (no treatment data used) |
| Data preparation | 7 | Pre-processing and quality checking, similarity across sociodemographic groups | Methods, Models and read-out (native preprocessing of each model); Supplementary Methods; identical pipeline for all subgroups |
| Outcome | 8a | Definition of the outcome, time horizon, consistency across sociodemographic groups | Methods, Cohorts and endpoints; Supplementary Table S2; label-type analysis (Supplementary Fig. S17) |
| | 8b | Outcome assessors | Not applicable (labels from clinical records, device statements and laboratory values) |
| | 8c | Blinding of outcome assessment | Not applicable |
| Predictors | 9a | Choice of predictors | Methods, Models and read-out (ECG waveform embeddings only; no demographic input) |
| | 9b | Definition and measurement of predictors | Methods, Models and read-out; Supplementary Table S4 |
| | 9c | Predictor assessors | Not applicable |
| Sample size | 10 | How the study size was arrived at | Methods, Statistics (all eligible ECGs; small-$n$ rule, Supplementary Table S6); Supplementary Table S2 |
| Missing data | 11 | Handling of missing data | Supplementary Methods and Supplementary Figs S1–S3 (ECGs with unknown age or sex kept for overall metrics, excluded from the corresponding subgroup axis) |
| Analytical methods | 12a | Use of data for development and evaluation, partitioning | Methods, Models and read-out (patient-grouped five-fold out-of-fold prediction); external cohorts (Fig. 2) |
| | 12b | Handling of predictors | Methods, Models and read-out; Supplementary Methods |
| | 12c | Type of model, model-building steps, hyperparameter tuning, internal validation | Methods, Models and read-out, Mitigation strategies; Supplementary Methods |

| Section/topic | Item | Checklist item (abridged) | Where reported |
|---|---|---|---|
| | 12d | Heterogeneity across clusters | Cohorts analysed separately and never pooled (Methods); Fig. 2B; Supplementary Table S8 |
| | 12e | Performance measures and plots | Methods, Models and read-out, Statistics (AUROC, MAE, worst-group gap, calibration, net benefit) |
| | 12f | Model updating or recalibration | Fig. 4E–F; Supplementary Methods; Supplementary Table S9 |
| | 12g | How predictions were calculated | Code repository (Data sharing) |
| Class imbalance | 13 | Class-imbalance methods | Class-balanced training evaluated as a mitigation strategy (Methods; Fig. 4C) |
| Fairness | 14 | Approaches used to address fairness | Entire study: subgroup evaluation by age, sex and race; mitigation strategies (Methods; Fig. 4) |
| Model output | 15 | Model output and thresholds | Probabilities from linear probes; thresholds by sensitivity target (Methods; Fig. 4E; Supplementary Table S9) |
| Training vs evaluation | 16 | Differences between development and evaluation data | Methods, Cohorts and endpoints; Supplementary Table S3 (pretraining–evaluation overlap) |
| Ethical approval | 17 | Ethics approval and consent | Ethics and data-use statement |
| Funding | 18a | Funding and role of funders | Funding; Role of the funding source |
| Conflicts of interest | 18b | Conflicts of interest | Declaration of interests |
| Protocol | 18c | Protocol availability | Methods, Statistics (no protocol was published) |
| Registration | 18d | Registration | Methods, Statistics (not registered) |
| Data sharing | 18e | Availability of data | Data sharing; Supplementary Table S10 |
| Code sharing | 18f | Availability of code | Data sharing |
| Patient and public involvement | 19 | Patient and public involvement | Methods, Statistics (none) |
| Participants | 20a | Flow of participants, numbers with and without outcome | Supplementary Figs S1–S3; Supplementary Table S2 |
| | 20b | Characteristics per data source, differences across demographic groups | Supplementary Tables S1 and S2 |
| | 20c | Comparison of evaluation with development data | Supplementary Tables S1 and S3 |
| Model development | 21 | Numbers of participants and events in each analysis | Supplementary Tables S2, S6, S8 |
| Model specification | 22 | Full model specification and access | Supplementary Table S4 (weights, sources, licences); code repository |
| Model performance | 23a | Performance with confidence intervals, including key subgroups | Figs 2, 5; Supplementary Tables S8, S11 |
| | 23b | Heterogeneity across clusters | Fig. 2B, E; Supplementary Table S8 |
| Model updating | 24 | Results of model updating | Fig. 4E–F; Supplementary Table S9 |
| Interpretation | 25 | Overall interpretation including fairness | Discussion |
| Limitations | 26 | Limitations | Discussion, Limitations |
| Usability | 27a | Handling of poor-quality or unavailable input | Not applicable (no model is proposed for implementation); Discussion, clinical implications |
| | 27b | User interaction and expertise | Not applicable |
| | 27c | Next steps for research | Discussion, Toward mitigation |

### S4 Supplementary figures

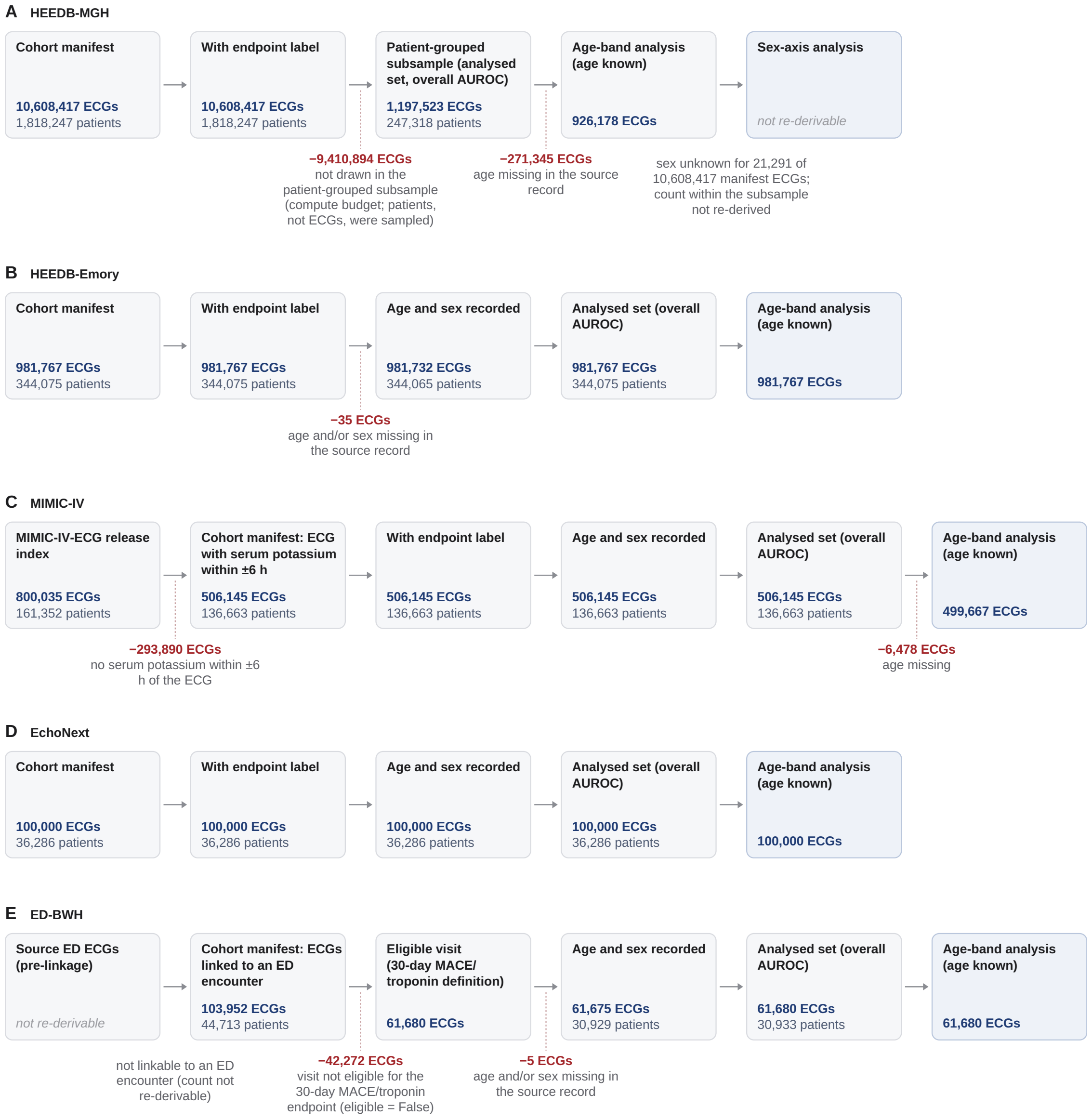

**Figure S1: Data curation flow, hospital/EHR-linked cohorts.** One flow per row, read from left to right: from the cohort manifest to the ECGs with an endpoint label, with recorded age and sex, the analysed set (overall AUROC) and the age-band analysis set; blue, ECGs and patients retained at each step; red numbers, ECGs removed at the preceding arrow, with the reason in grey. Counts were re-derived from the cohort manifests and the stored out-of-fold prediction files; age-band counts from the per-band tables. **(A)** HEEDB-MGH was probed on a patient-grouped subsample (patients, not ECGs, were sampled); the third box is this subsample, not a data-quality exclusion. **(B)** HEEDB-Emory. **(C)** MIMIC-IV-ECG starts from the release index and keeps ECGs with a serum potassium within ±6 h. **(D)** EchoNext. **(E)** ED-BWH starts from ECGs linked to an emergency-department encounter and keeps visits eligible for the 30-day MACE/troponin endpoint; the 3-lead and 12-lead inputs are the same 61,680 visits. ECGs with unknown sex or age were kept for the overall AUROC and dropped only from the corresponding subgroup axis (analysed set larger than the “age and sex recorded” box in HEEDB-Emory and ED-BWH). Two counts could not be re-derived and are left empty. Other cohorts: Supplementary Figs S2 and S3.

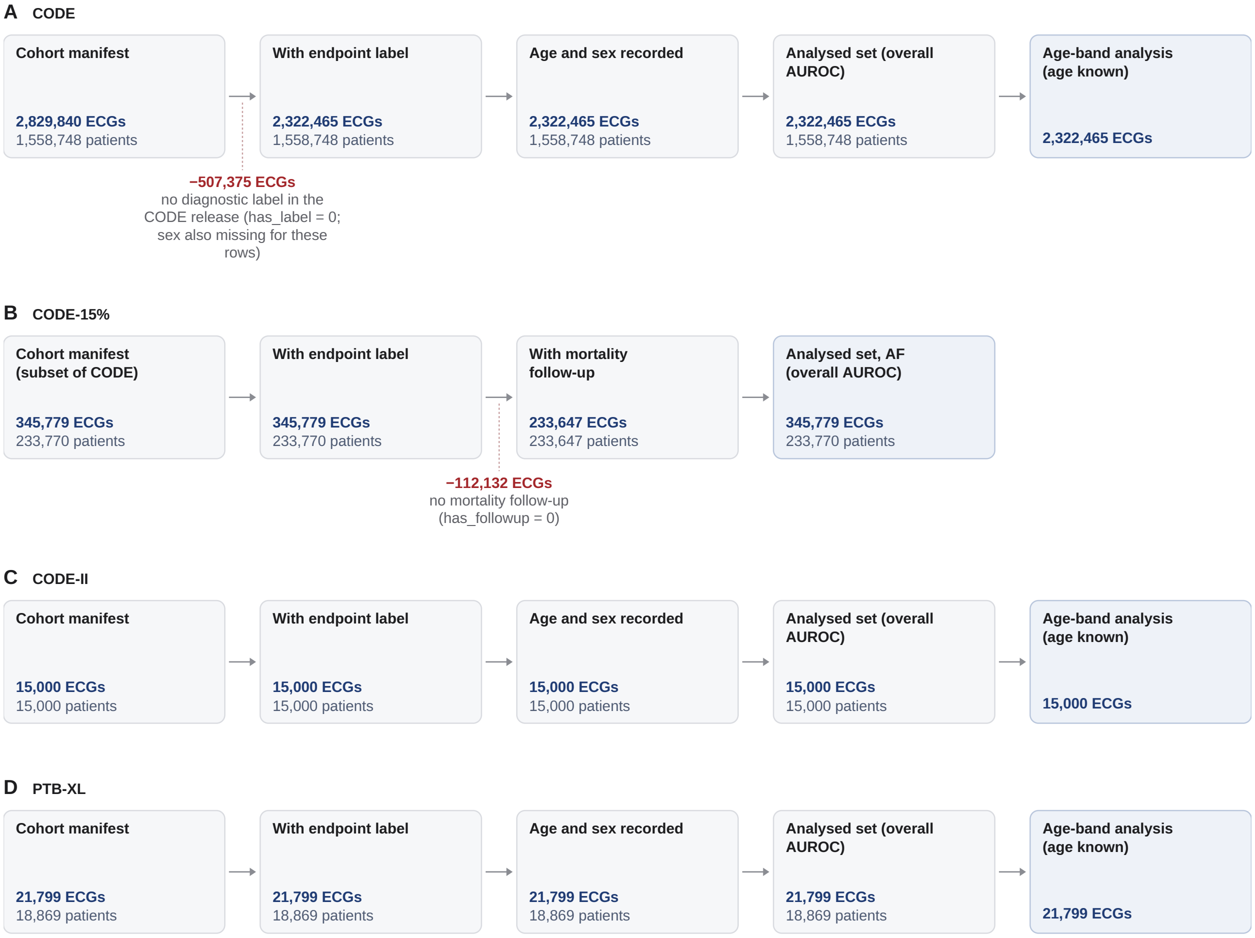

**Figure S2: Data curation flow, telehealth/benchmark cohorts.** Layout and colours as in Supplementary Fig. S1: one flow per row from the cohort manifest to the age-band analysis set; red numbers, ECGs removed at the preceding arrow, with the reason in grey. **(A)** CODE; the 507,375 ECGs without a diagnostic label in the release also lack a sex record. **(B)** CODE-15% is a subset of CODE and is counted once in the totals; its AF analysis uses the whole subset and the one-year mortality analysis the ECGs with follow-up, so the AF analysed set (345,779 ECGs) is larger than the preceding "with mortality follow-up" box (233,647 ECGs). **(C)** CODE-II. **(D)** PTB-XL. ECGs with unknown sex or age were kept for the overall AUROC and dropped only from the corresponding subgroup axis.

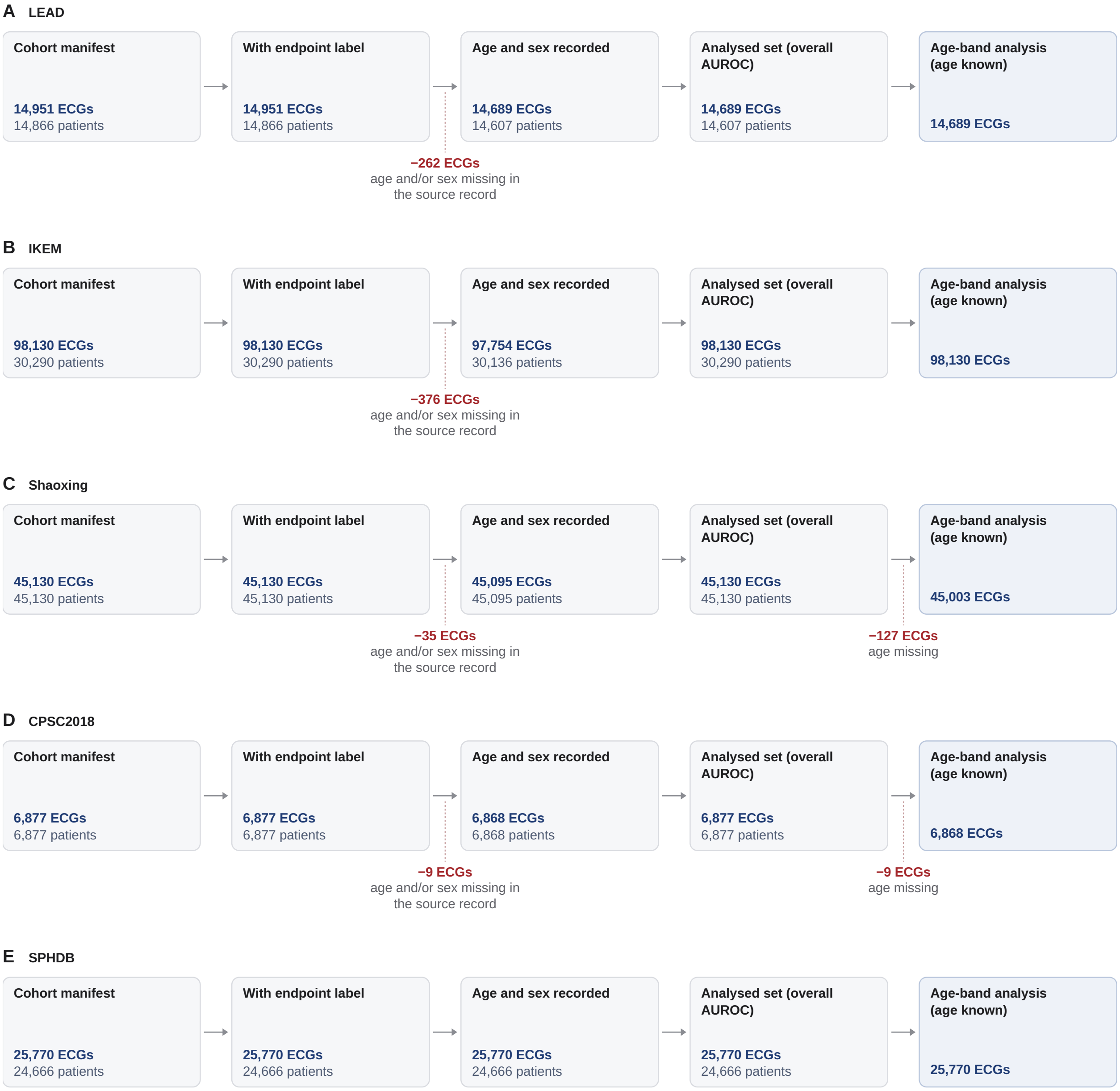

**Figure S3: Data curation flow, external/public cohorts.** Layout and colours as in Supplementary Fig. S1: one flow per row from the cohort manifest to the age-band analysis set; red numbers, ECGs removed at the preceding arrow, with the reason in grey. **(A)** LEAD. **(B)** IKEM. **(C)** Shaoxing. **(D)** CPSC2018. **(E)** SPHDB. ECGs with unknown sex or age were kept for the overall AUROC and dropped only from the corresponding subgroup axis, so the analysed set can be larger than the preceding "age and sex recorded" box (IKEM, Shaoxing, CPSC2018); the age-band set then drops the ECGs with unknown age again (Shaoxing, CPSC2018).

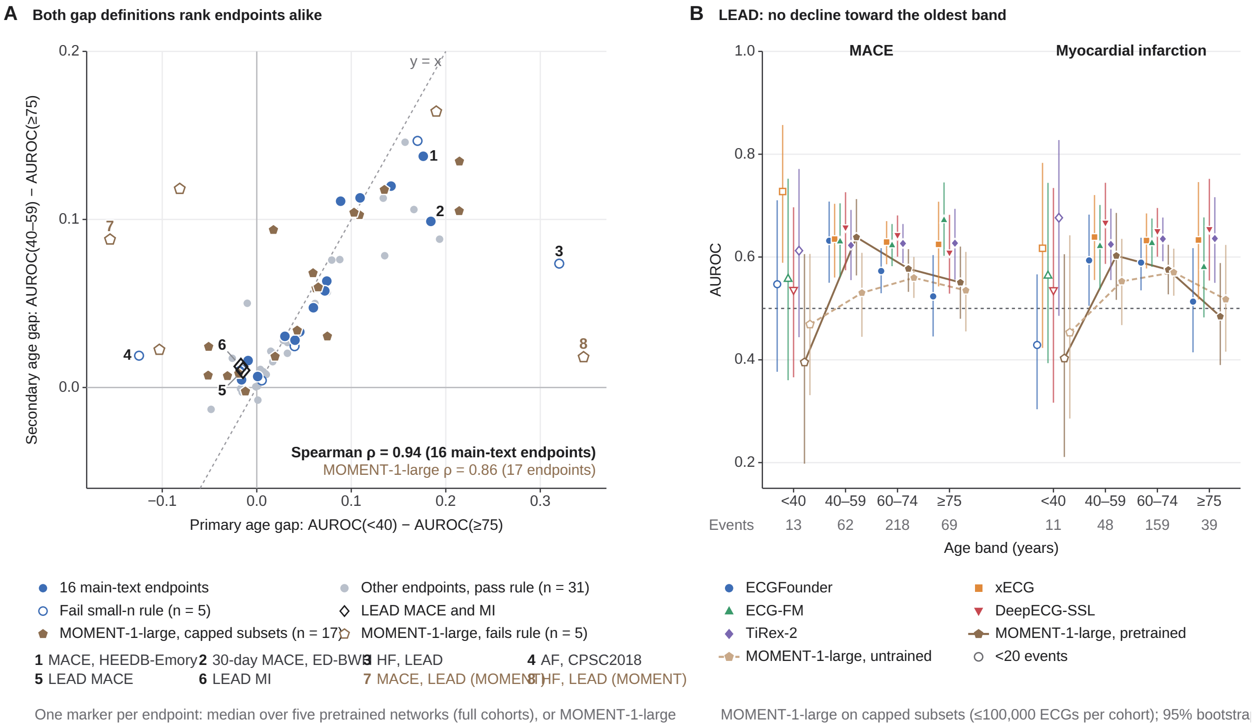

**Figure S4: The two age-gap definitions agree, and LEAD enters on the secondary definition.** **(A)** Primary age gap (AUROC <40 minus ≥75 years) against the secondary adjacent-band gap (40–59 minus ≥75), one marker per endpoint. Circles: median over the five pretrained ECG/time-series networks on the full cohorts (filled blue, the 16 main-text endpoints; small grey, 31 other classification endpoints passing the small-$n$ rule; open, 5 endpoints failing it); black diamonds, LEAD MACE and MI, for which only the secondary definition is valid; brown pentagons, MOMENT-1-large (capped subsets, ≤100,000 ECGs per cohort; 22 endpoints, open where the endpoint fails the small-$n$ rule on the capped subset). Spearman $\rho$ = 0.94 over the 16 main-text endpoints and 0.86 over the 17 MOMENT-1-large endpoints passing the rule. Dotted line, identity; each endpoint appears once (duplicate task keys removed), and the axes cover every marker. **(B)** AUROC by age band for LEAD MACE and MI per network with 95% bootstrap CIs (1,000 resamples): the five pretrained networks (full cohort; markers only) and MOMENT-1-large pretrained (brown, solid line) and its untrained copy (light brown, dashed line), both on the capped subsets (≤100,000 ECGs per cohort; LEAD is complete at this cap). Open markers, bands with fewer than 20 events (the <40 band, 13 and 11 events; counts below the axis). LEAD recruited participants aged 6–80 years, so its oldest band spans 75–80 years.

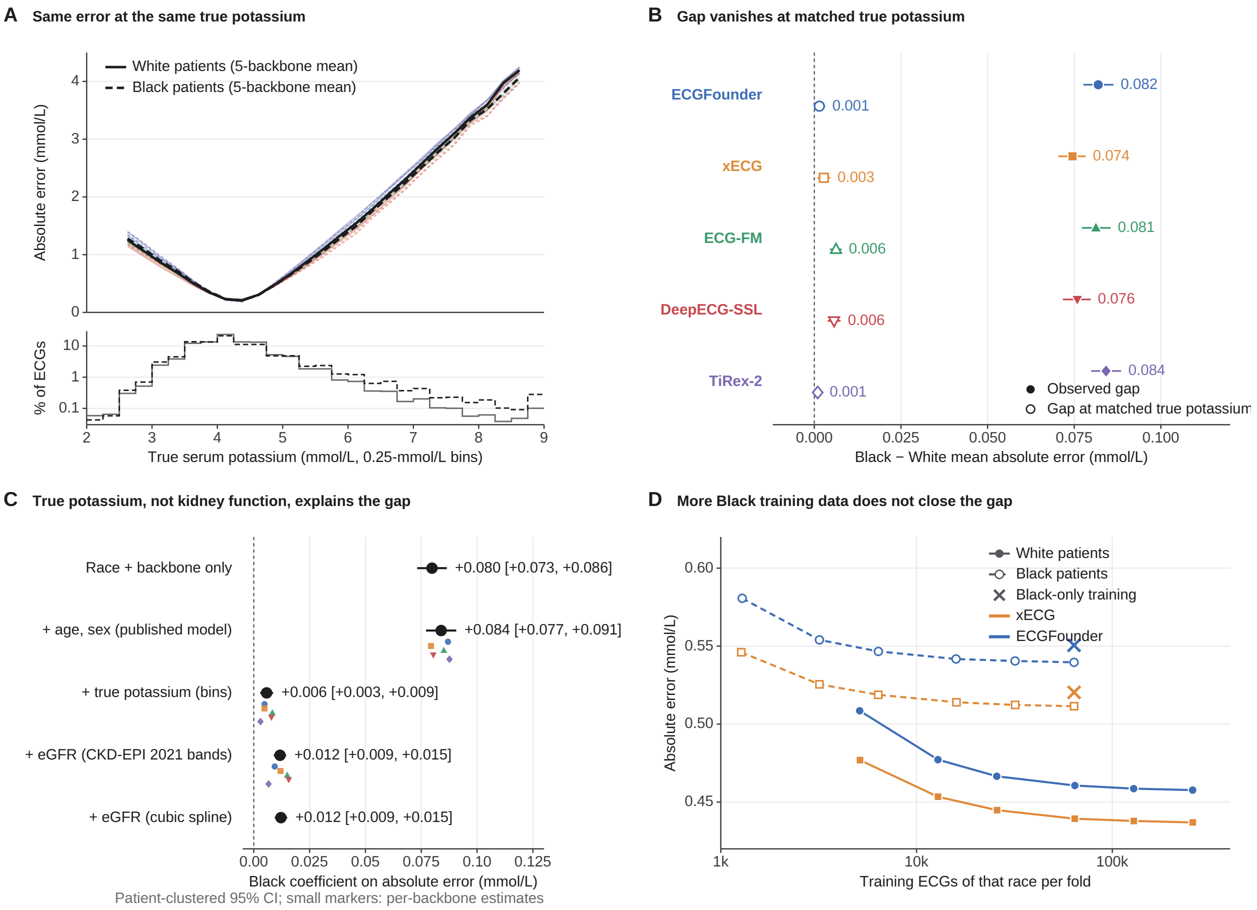

**Figure S5: The Black–White potassium error gap at matched true potassium.** (A) Absolute error against true potassium (0.25 mmol/L bins) for White and Black patients, with the potassium distributions below. (B) Observed gap and gap after standardising Black patients' error to the White potassium distribution, per backbone. (C) Black coefficient on absolute error in regressions with successive adjustment (patient-clustered 95% CIs). (D) Error by number of training ECGs of each race per fold, and with Black-only training (ECGFounder and xECG). Standardised gaps for the five backbones are 0.001–0.006 mmol/L.

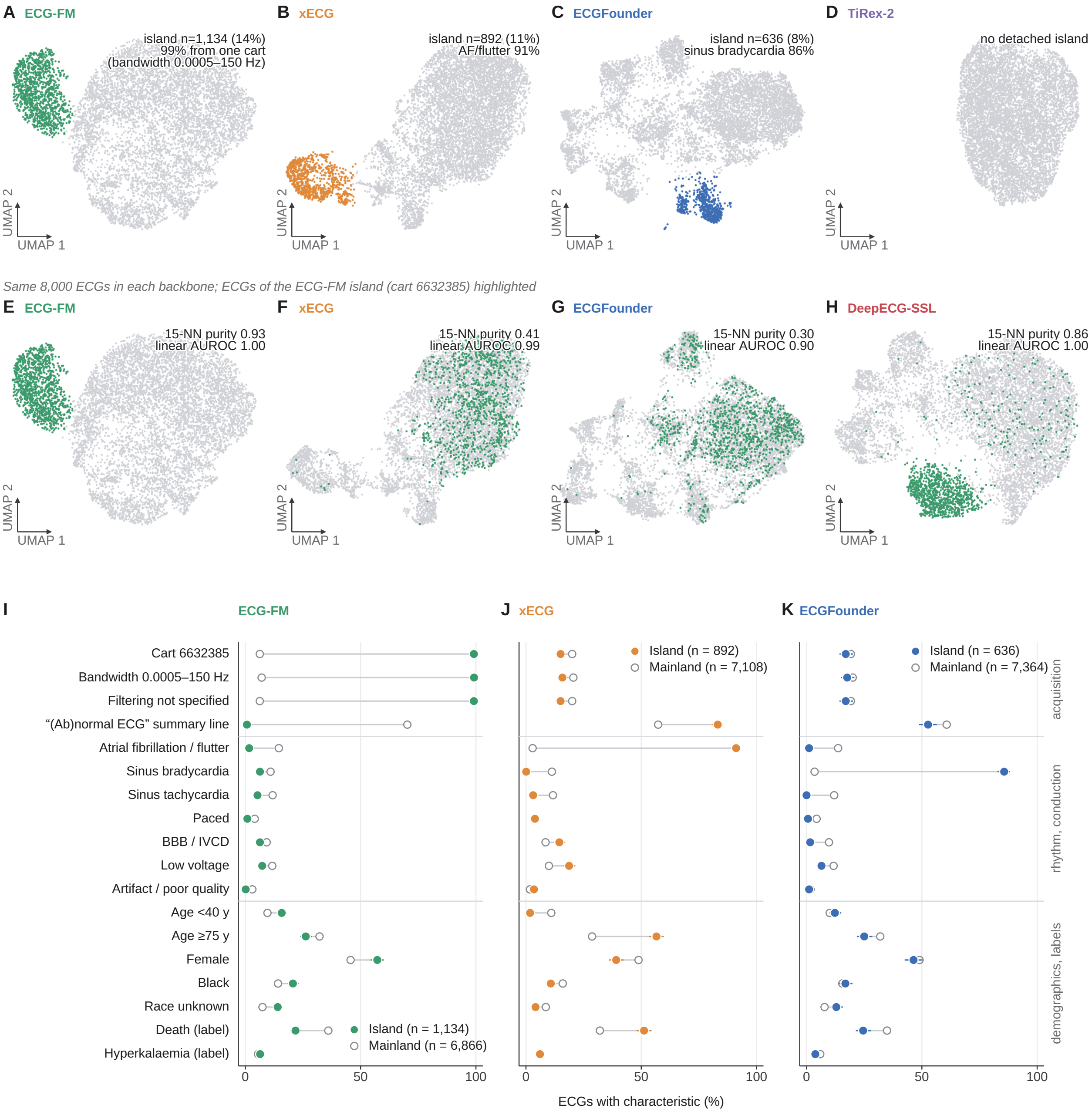

**Figure S6: The detached cluster in the MIMIC-IV embedding is an acquisition-device group and is backbone-specific.** (A–D) Each backbone's projection of the same 8,000 MIMIC-IV-ECG test ECGs with its own detached cluster highlighted, if any: ECG-FM, one ECG cart (1,134 ECGs); xECG, AF and flutter; ECGFounder, sinus bradycardia; TiRex-2, none. (E–H) The ECG-FM cluster ECGs (green) in each backbone's projection, with 15-nearest-neighbour purity in the full embedding space and linear-probe AUROC for cluster membership (ECG-FM 0.93 and 1.00; DeepECG-SSL 0.86 and 1.00; xECG 0.41 and 0.99; ECGFounder 0.30 and 0.90). (I–K) Cluster versus rest of the sample for acquisition, rhythm and demographic characteristics (95% Wilson CIs); in ECG-FM, 99.1% of cluster ECGs came from cart 6632385 (6.3% of the rest).

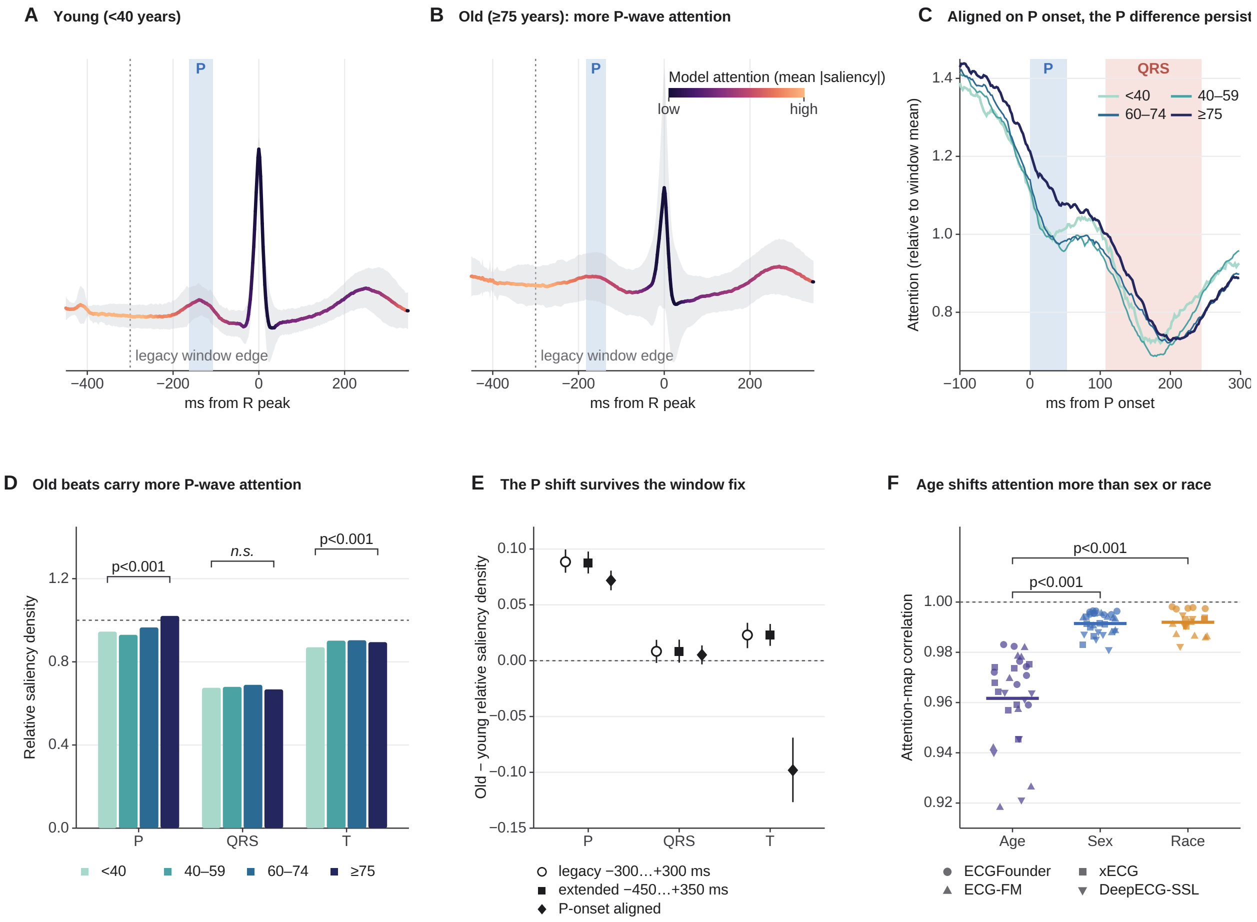

**Figure S7: Attention shifts toward the P wave with age in two backbones, and the shift survives the window definition.** SmoothGrad saliency of the four ECG backbones, eight endpoints (PTB-XL, EchoNext, MIMIC-IV-ECG; 30 cells); beats R-aligned, $-450$ to $+350$ ms; dotted line, edge of the standard $-300$ ms window. **(A, B)** Mean lead-II beat of patients $<40$ (6,941 beats) and $\geq 75$ years (19,906), coloured by mean absolute saliency, with P-wave detail. **(C)** Saliency aligned on P-wave onset, by age band. **(D)** Relative saliency density by wave and age band. **(E)** Old-minus-young difference by wave for the three windows; bars, 95% per-beat bootstrap CIs. **(F)** Correlation of mean saliency maps between subgroups, one marker per endpoint and backbone. Wave blanking: Supplementary Fig. S8.

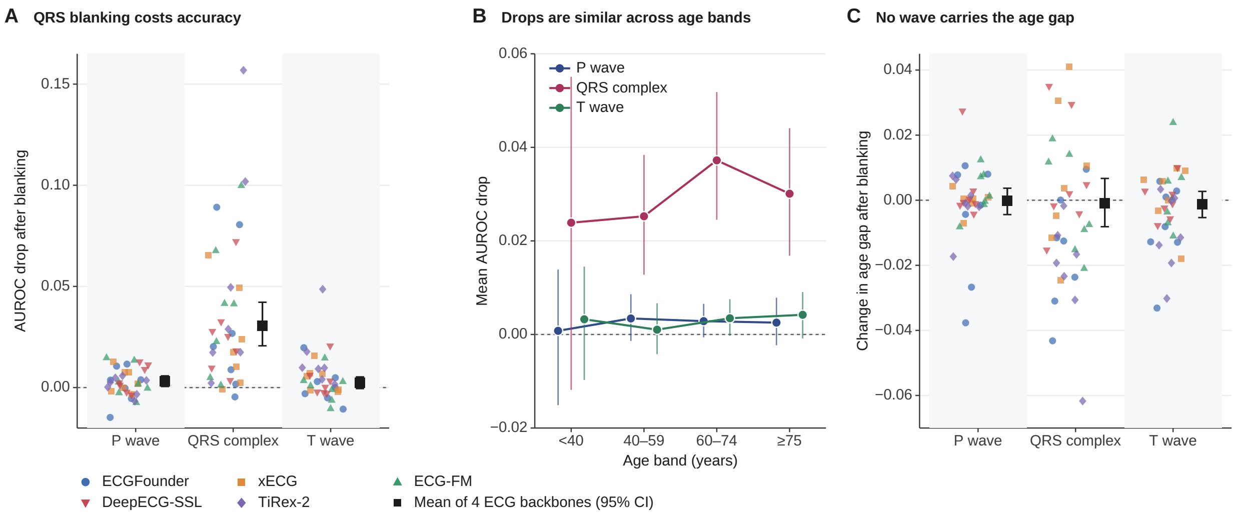

**Figure S8: Wave blanking.** (A) AUROC drop after setting the delineated P wave, QRS complex or T wave to zero (unblanked minus blanked; one marker per endpoint–backbone result, 35 results including TiRex-2, diamonds; black, mean over the 28 results of the four ECG backbones with 95% bootstrap CI). Four ECG backbones: P wave 0.0031 (0.0005 to 0.0058), QRS complex 0.0307 (0.0206 to 0.0422), T wave 0.0024 (−0.0005 to 0.0053); with TiRex-2: P wave 0.0027, QRS complex 0.0352, T wave 0.0048. (B) Mean drop by age band, four ECG backbones. (C) Change in the age gap (overall minus worst-band AUROC); P wave −0.0001 (−0.0044 to 0.0037) for the four ECG backbones and −0.0003 with TiRex-2.

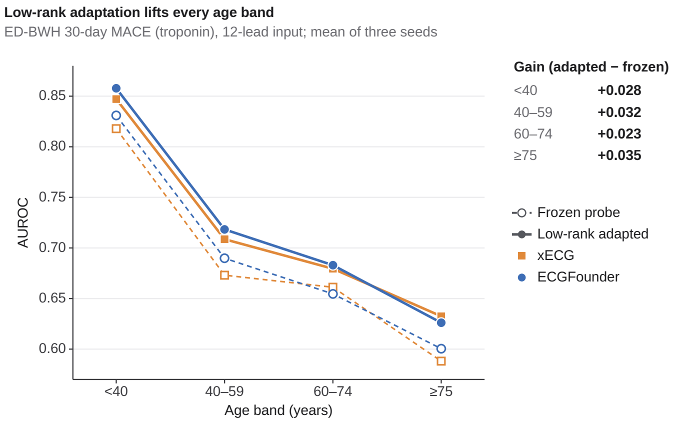

**Figure S9: Low-rank adaptation raises AUROC in every age band.** AUROC by age band for 30-day MACE in ED-BWH (12-lead input) with frozen probes (dashed, open) and after low-rank adaptation (solid), ECGFounder and xECG, mean of three seeds (6 runs); the gain at 75 years or older (+0.035) is matched by the gain below 40 years (+0.028).

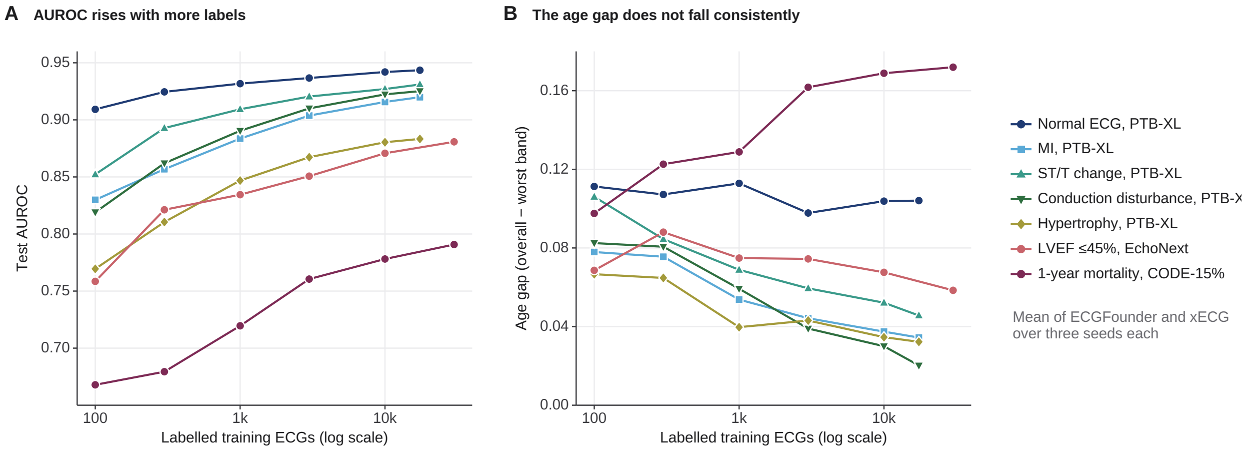

**Figure S10: Learning curves per endpoint.** (A) Test AUROC and (B) age gap (overall minus worst band) against the number of labelled training ECGs for the seven endpoints of Fig. 4B, mean of ECGFounder and xECG and of three seeds. The largest training size is 30,000 ECGs or the full training set of the endpoint, whichever is smaller (PTB-XL endpoints: 17,418).

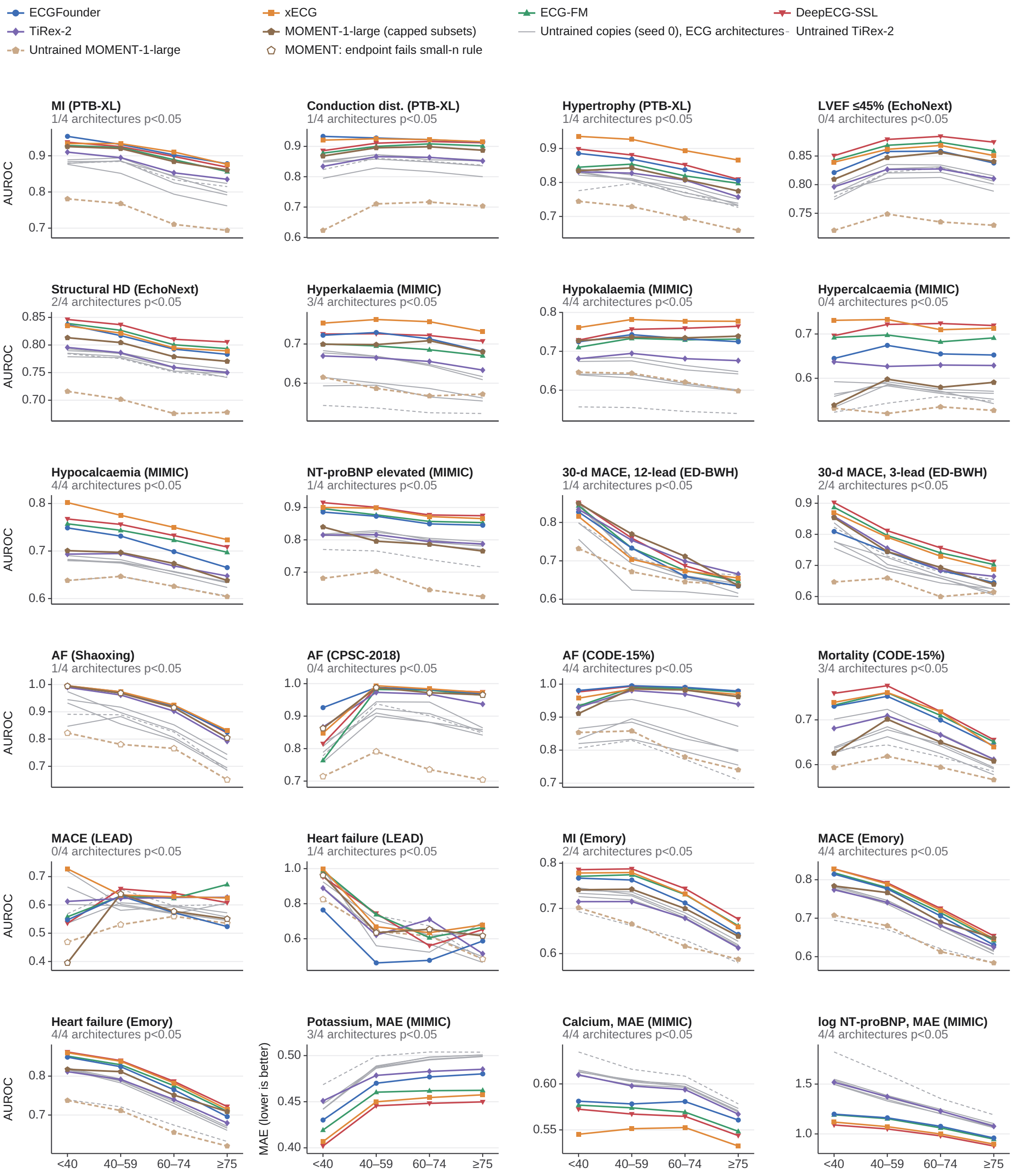

**Figure S11: Untrained networks reproduce the age pattern: performance by age band.** AUROC (or MAE for the three regression endpoints, lower = better) by age band for each pretrained backbone (colour) and its untrained copy (one random initialisation, seed 0; grey, dashed for TiRex-2), for the 24 null-control endpoints (21 classification, 3 regression), on the full cohorts. The 21 classification sub-panels additionally show MOMENT-1-large (capped subsets, ≤100,000 ECGs per cohort; brown, solid) and its untrained copy (light brown, dashed), evaluated under the control protocol on patient-grouped subsets; open MOMENT markers denote endpoints failing the small-$n$ rule (<20 positives in an age band: AF in Shaoxing and CPSC-2018, MACE and heart failure in LEAD). Sub-panel titles give the number of the four ECG architectures with a significant paired difference in age gap between pretrained and untrained copy (paired $p < 0.05$); this count excludes MOMENT-1-large. Age bands: <40, 40–59, 60–74, ≥75 years.

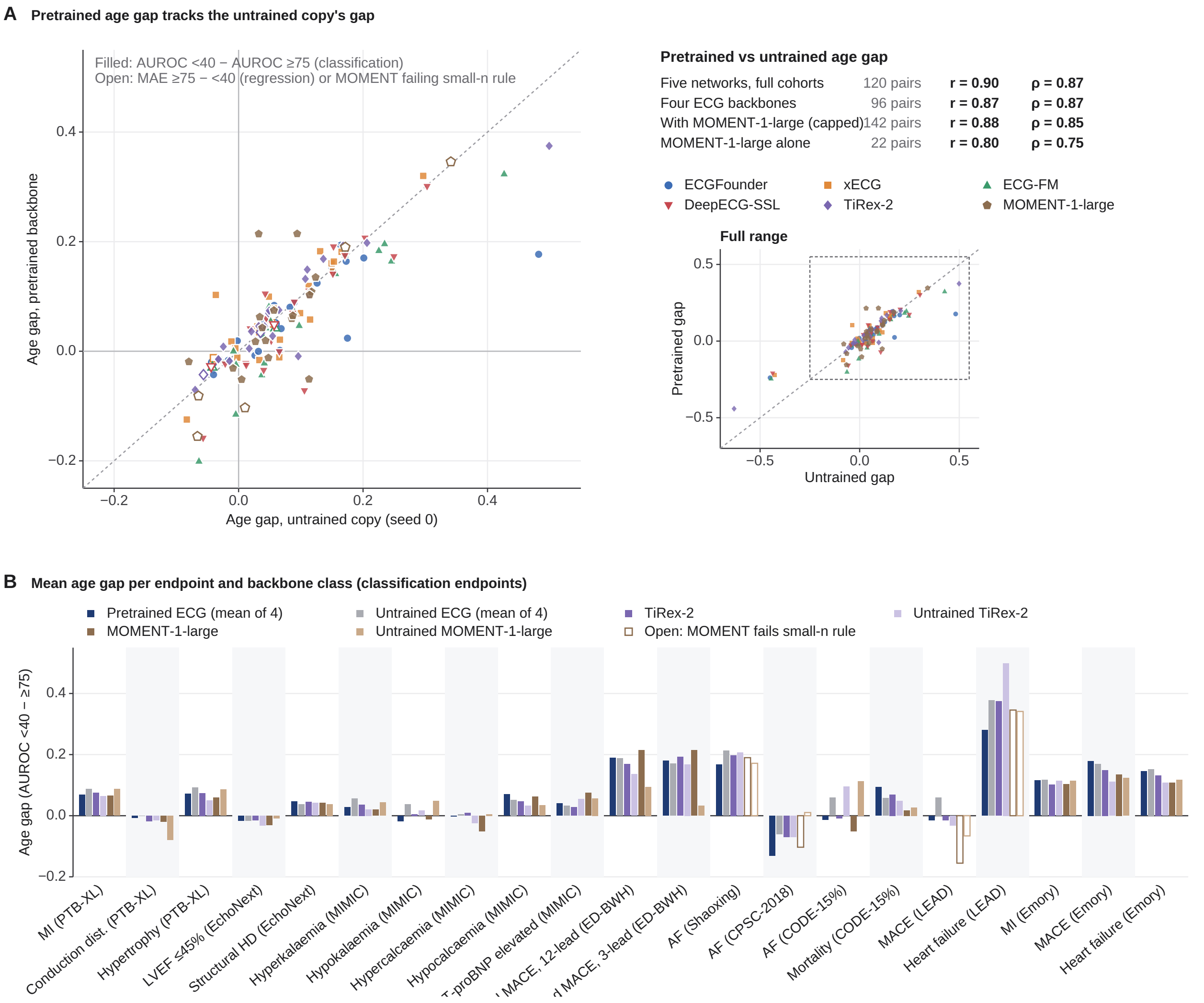

**Figure S12: Untrained networks reproduce the age pattern: age gaps.** (A) Age gap of each pretrained backbone against that of its untrained copy, all 24 endpoints without the small-$n$ rule (five networks on the full cohorts: 120 pairs, Pearson r=0.90, Spearman $\rho$=0.87; four ECG backbones r=0.87, 96 pairs), plus MOMENT-1-large (capped subsets, ≤100,000 ECGs per cohort; brown pentagons, 22 classification endpoints; with these 142 pairs r=0.88, $\rho$=0.85; MOMENT-1-large alone r=0.80, $\rho$=0.75). Filled markers, classification endpoints (gap = AUROC under 40 minus AUROC at 75 years or older); open markers of the five networks, regression endpoints (MAE at 75 years or older minus under 40 years, in each endpoint's units), which Fig. 5B omits; open MOMENT markers, endpoints failing the small-$n$ rule. The main axes show the 137 pairs within −0.25 to 0.55; the five log NT-proBNP (MAE) pairs (untrained gap −0.43 to −0.63) are shown in the full-range axes at the right together with all other pairs (dashed rectangle, extent of the main axes). Dotted line, identity. (B) Mean age gap per classification endpoint for pretrained and untrained ECG backbones (mean of four), TiRex-2 and its untrained copy (full cohorts), and MOMENT-1-large and its untrained copy (capped subsets; light brown for the untrained copy; open bars, endpoints failing the small-$n$ rule).

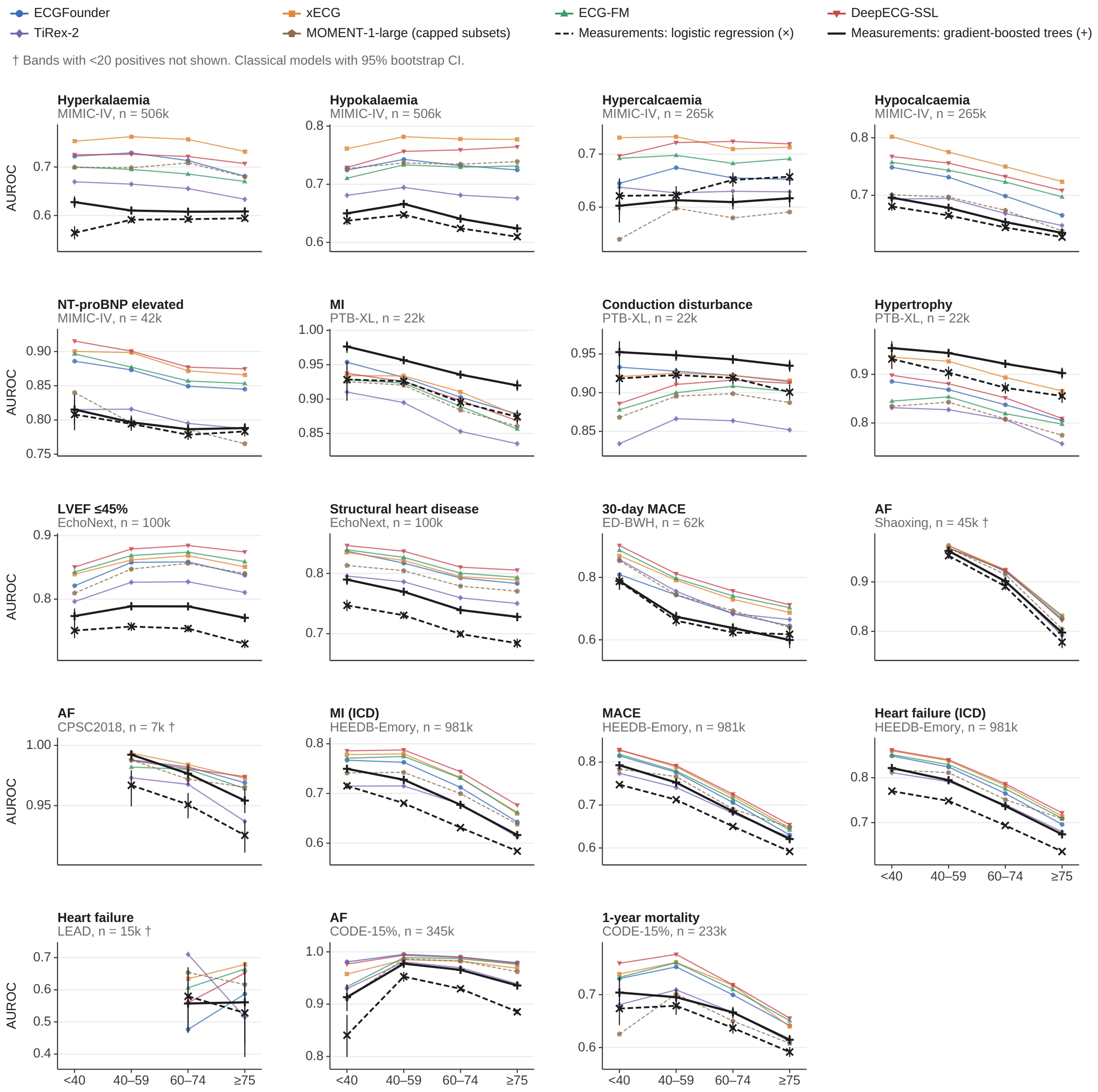

**Figure S13: Age-band AUROC of classical-measurement models and of the pretrained probes.** Logistic regression (dashed, crosses) and gradient-boosted trees (solid black) on standard ECG measurements, the five pretrained ECG/time-series networks (colour, full cohorts) and MOMENT-1-large pretrained (brown, dashed; capped subsets, ≤100,000 ECGs per cohort) for the 19 endpoints of Fig. 5C, 16 of which pass the small-$n$ rule; the three marked with a dagger (AF in Shaoxing and CPSC2018, heart failure in LEAD) fail it. Bands with fewer than 20 positive ECGs are not shown. Each sub-panel has its own AUROC range; $n$ is the number of ECGs of the endpoint.

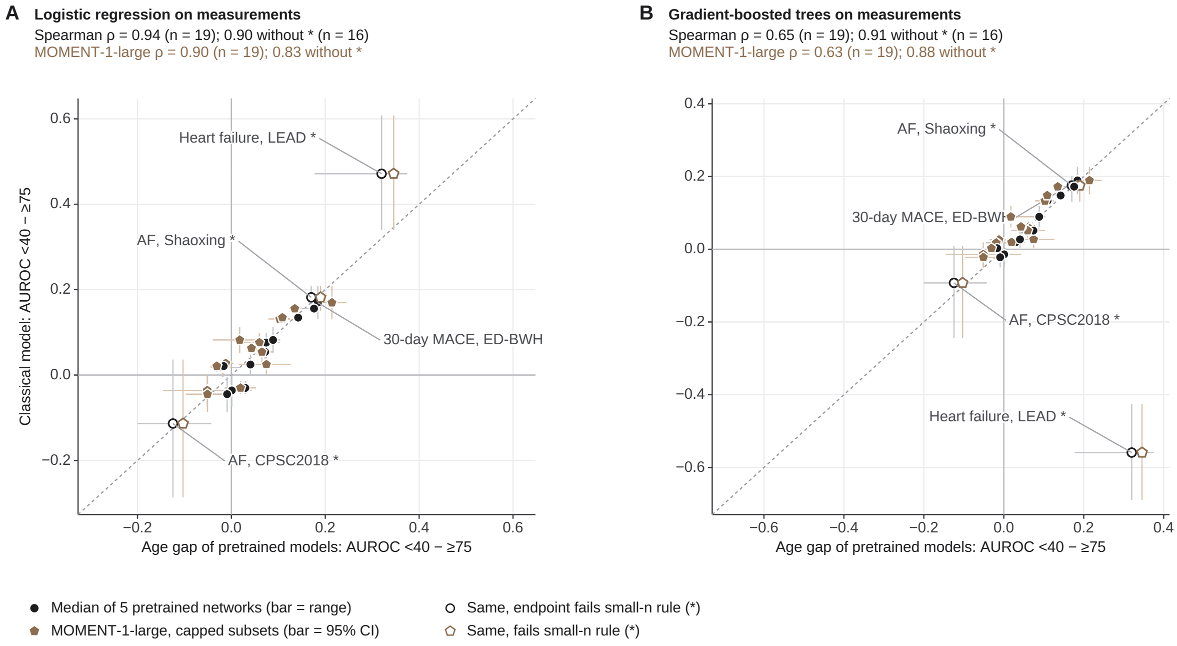

**Figure S14: Age gaps of classical-measurement models against the pretrained probes.** Age gap (AUROC under 40 years minus AUROC at 75 years or older) of (A) logistic regression and (B) gradient-boosted trees on ECG measurements against the median gap of the five pretrained ECG/time-series networks on the full cohorts (black circles; bars: 95% bootstrap CI and range over networks) and against the gap of MOMENT-1-large (brown circles; capped subsets, $\leq$100,000 ECGs per cohort; bar: 95% bootstrap CI). Open markers: the endpoints failing the small-$n$ rule (CPSC2018 AF, LEAD heart failure, Shaoxing AF). Classical gap versus the five-network median: Spearman $\rho$ 0.90 (p=2e-06) and 0.91 (p=7e-07) for the 16 endpoints passing the rule, and 0.94 and 0.65 including all 19. Classical gap versus the MOMENT-1-large gap: $\rho$ 0.83 and 0.88 for the 16 endpoints passing the rule, and 0.90 and 0.63 including all 19. Dotted line: identity.

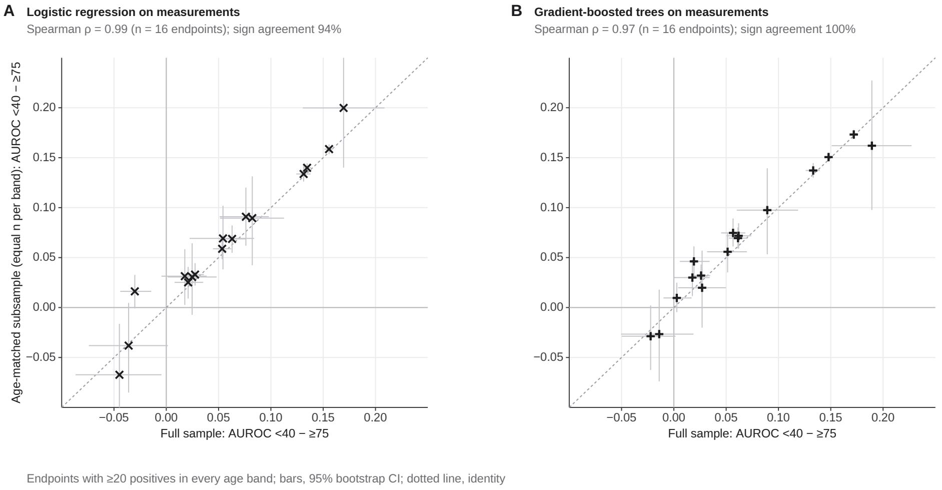

**Figure S15: Classical-model age gaps with equal numbers of ECGs per age band.** Age gap in the full sample against the gap after refitting on a subsample with equal numbers of ECGs in each band (Spearman $\rho$ 0.99 logistic regression, 0.97 gradient-boosted trees; 16 endpoints passing the small-n rule).

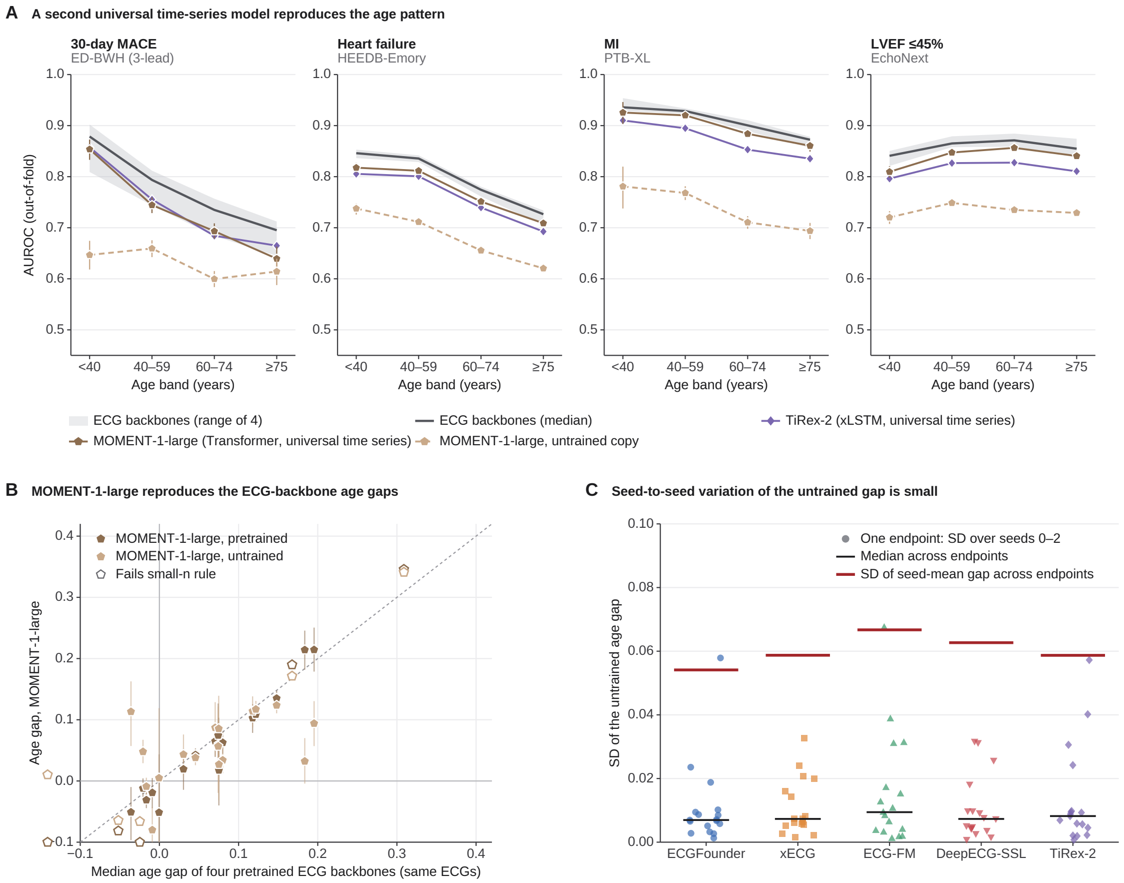

**Figure S16: A second non-ECG time-series model reproduces the age gap, and the untrained gap is stable across random initialisations.** **(A)** AUROC by age band for three endpoints with an age gap and one without: four pretrained ECG backbones (grey band, range; dark grey line, median), TiRex-2 (xLSTM [S26], purple) and MOMENT-1-large (Transformer; pretrained, brown; untrained copy, light brown dashed; 95% bootstrap CIs shown for the MOMENT series), all on the same patient-grouped subsets (≤100,000 ECGs per cohort). **(B)** Age gap of MOMENT-1-large (pretrained and untrained) against the median gap of the four pretrained ECG backbones on the same ECGs, 22 endpoints; open markers fail the small-$n$ rule; dashed line, identity. **(C)** Across-seed SD of the untrained age gap (seeds 0–2) per endpoint and architecture, 17 endpoints passing the small-$n$ rule; black, median across endpoints; red, SD of the seed-mean gap across endpoints.

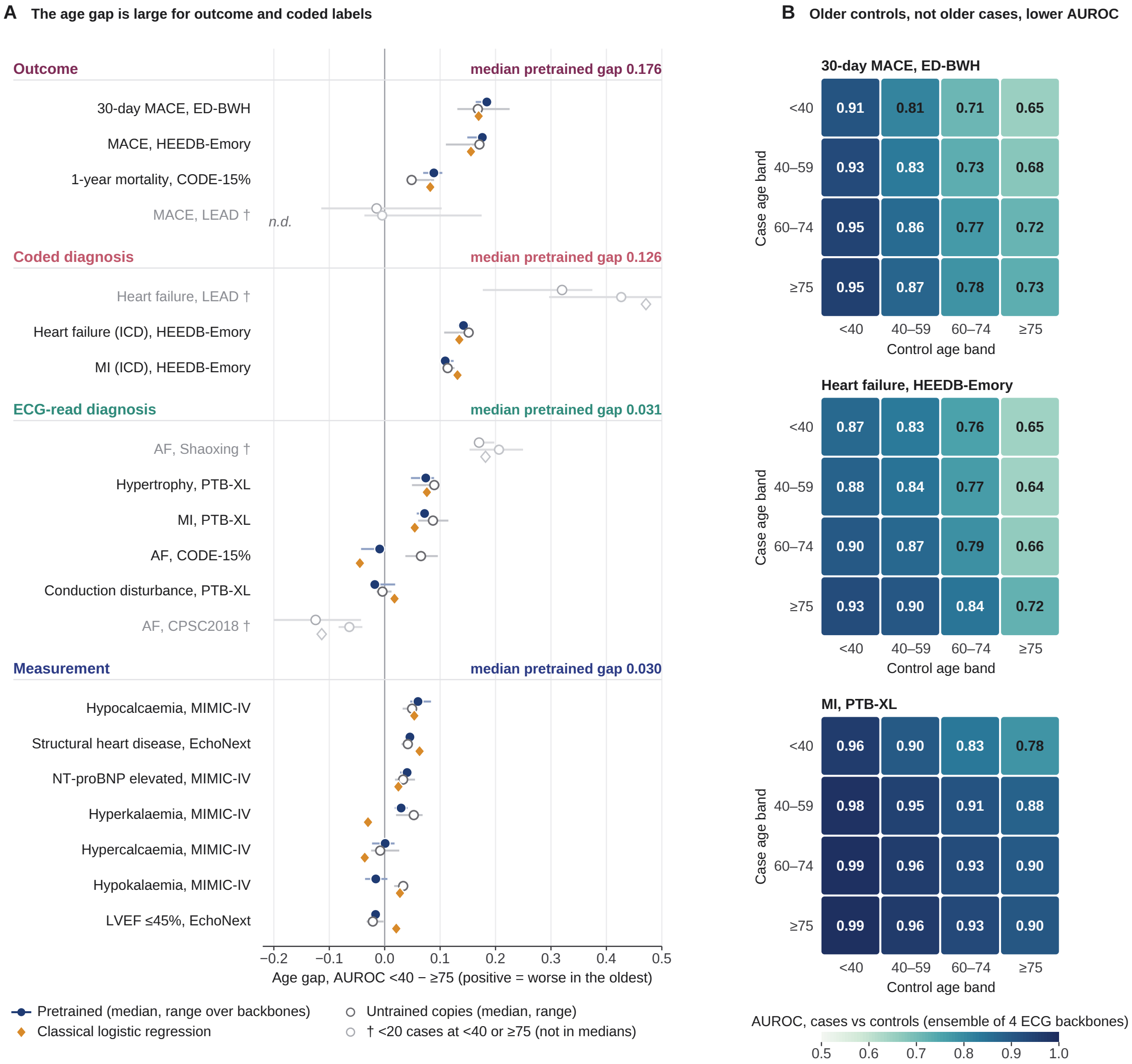

**Figure S17: The age gap is concentrated where labels depend on more than the ECG and where older non-cases resemble cases.** (A) Age gap per classification endpoint for the pretrained models (median and range), their untrained copies and classical logistic regression (n.d., not done), grouped by how the label is obtained; group medians in the headers. Grey (†), fewer than 20 positive ECGs in the youngest or oldest band: shown for completeness and excluded from the group medians. (B) AUROC of cases from each age band against controls from each age band, ensemble of the four ECG backbones, from stored out-of-fold predictions; AUROC depends more on the age of the controls (columns) than on that of the cases (rows).

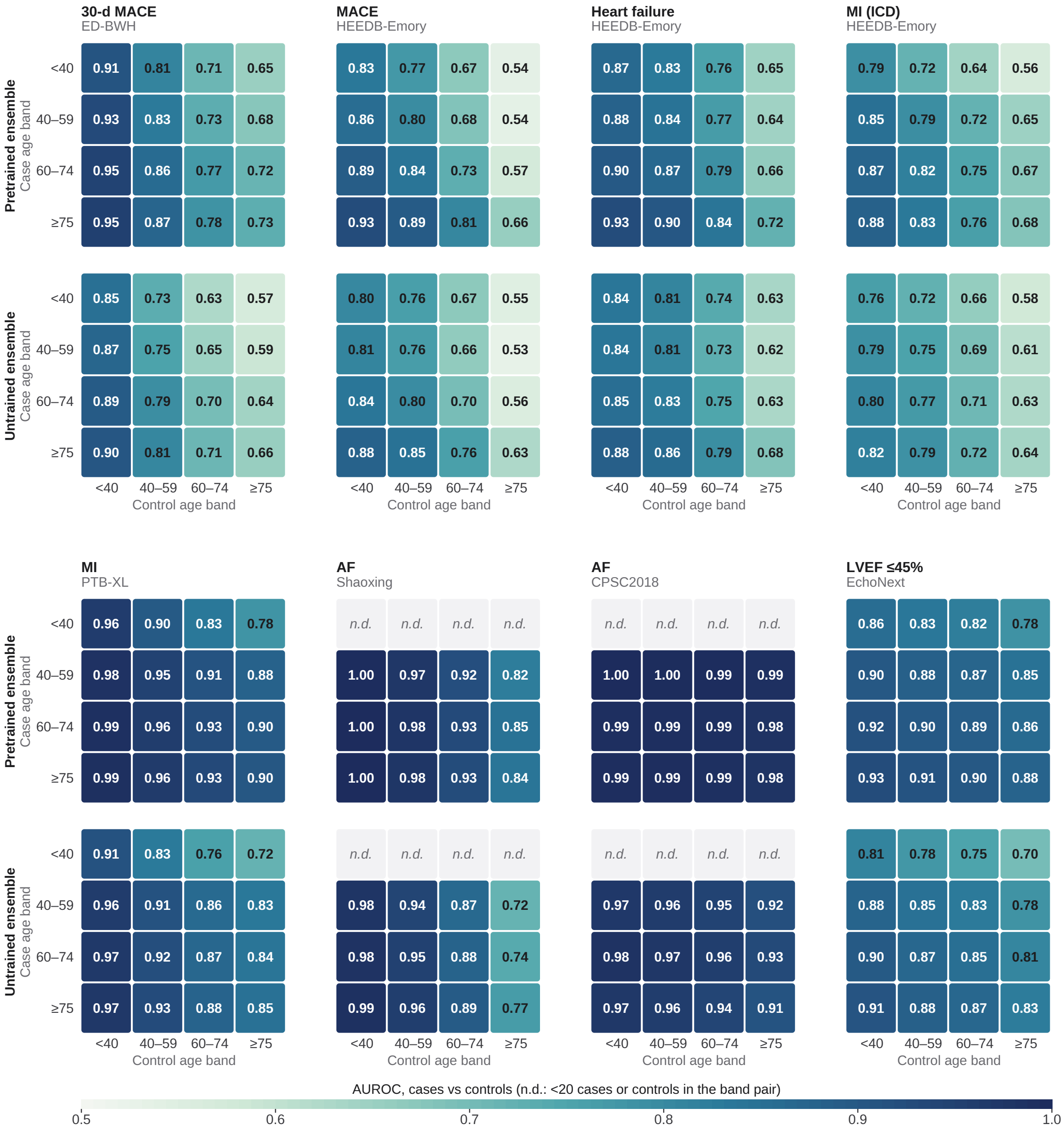

**Figure S18: Case-age by control-age AUROC for pretrained and untrained ensembles.** AUROC of cases from each age band against controls from each age band, for the ensemble of the four pretrained ECG backbones (top) and of their untrained copies (bottom), from stored out-of-fold predictions on the ECGs scored by all networks. n.d., fewer than 20 cases or controls.

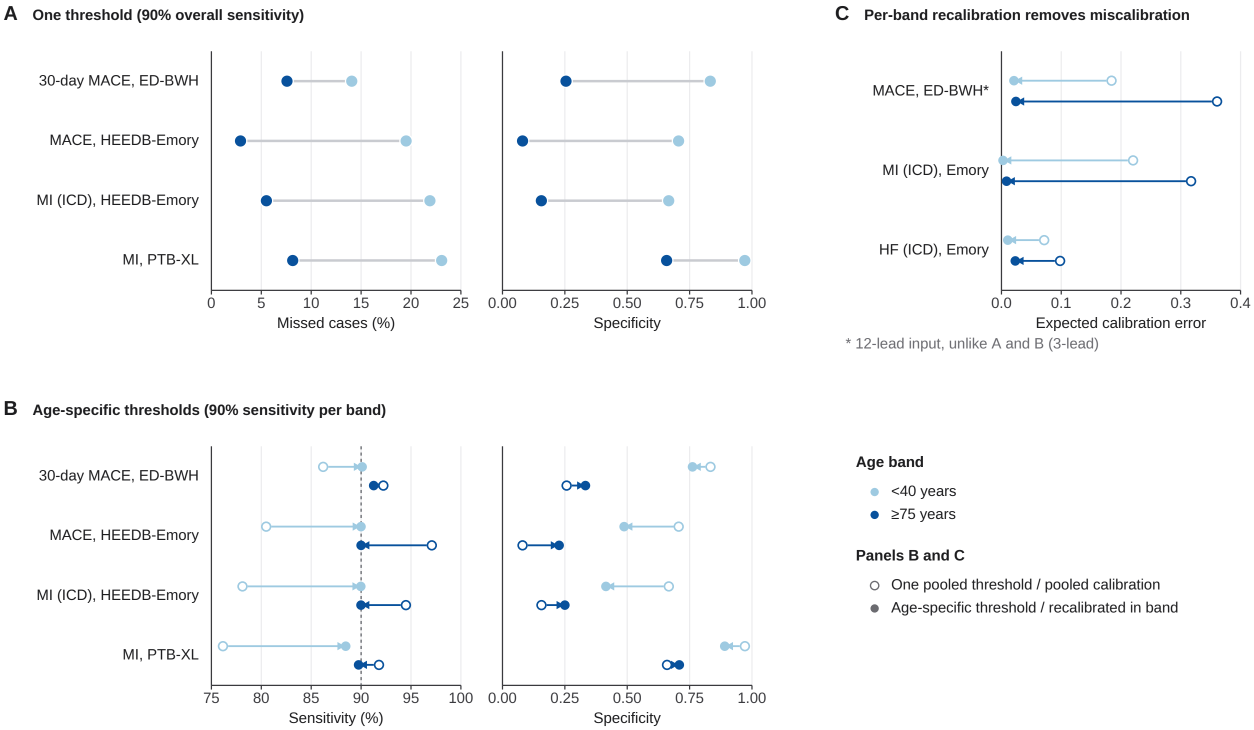

**Figure S19: At one threshold the type of error changes with age; age-specific thresholds and per-band recalibration manage it.** (A) Missed cases and specificity below 40 and at 75 years or older at one threshold giving 90% overall sensitivity (ensemble of the four ECG backbones; values and CIs in Supplementary Table S9). (B) Sensitivity and specificity in the same bands with one threshold (open circles) and with age-specific thresholds giving 90% sensitivity within each band (arrow heads); both chosen on one half of patients and evaluated on the other. (C) Expected calibration error below 40 and at 75 years or older with the pooled calibration (open) and after recalibration within each band (filled), ensemble of the four ECG backbones (*ED-BWH 12-lead input).

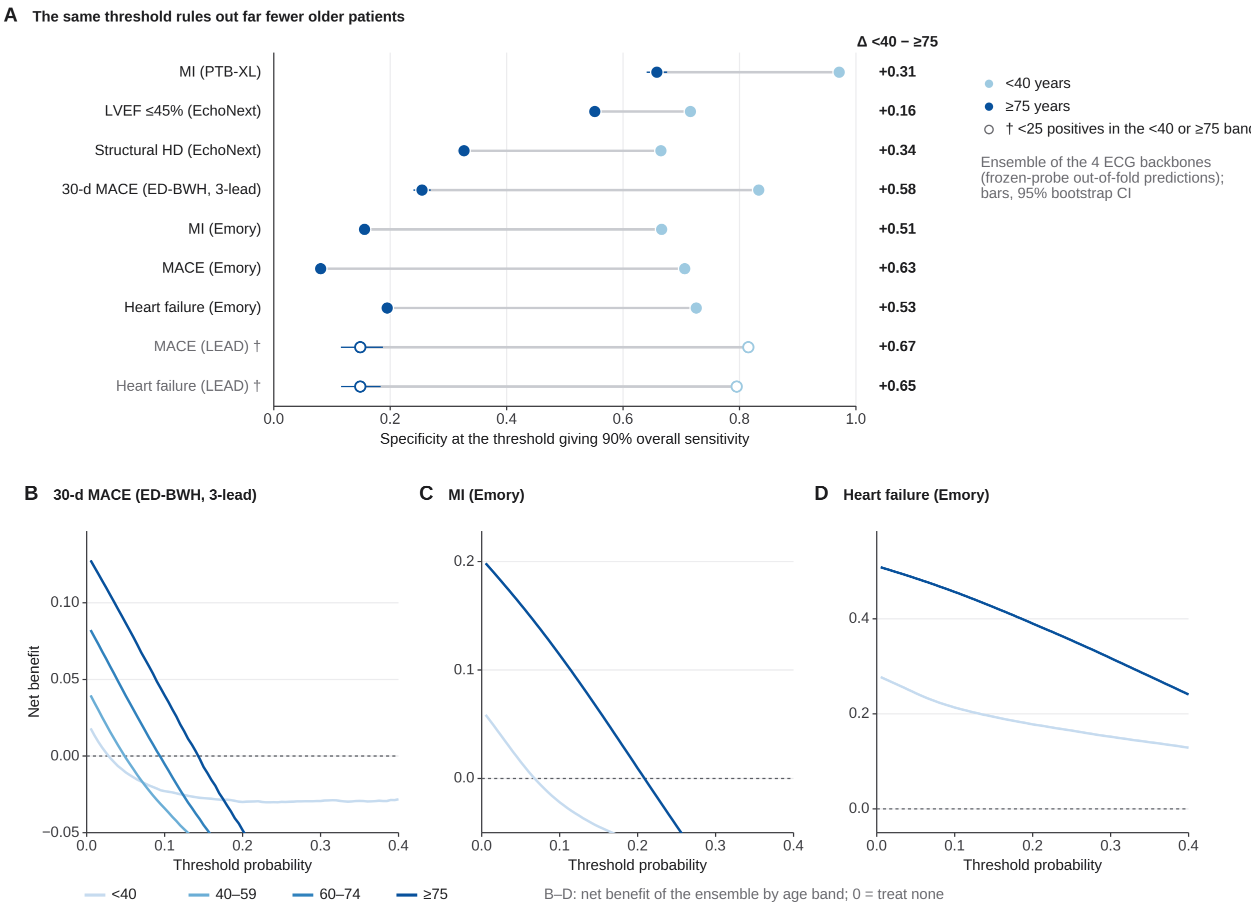

**Figure S20: The same threshold rules out fewer older patients.** (A) Specificity under 40 years and at 75 years or older at the threshold giving 90% overall sensitivity, for the ensemble of the four ECG backbones (out-of-fold frozen-probe predictions; bars, 95% bootstrap CIs; open markers, fewer than 25 positive ECGs in an extreme band). (B–D) Net benefit by age band for ED-BWH 30-day MACE (3-lead), HEEDB-Emory MI and HEEDB-Emory heart failure; 0 corresponds to treating none. Higher net benefit in older patients reflects their higher event rate, not better discrimination. Values in Supplementary Table S9.

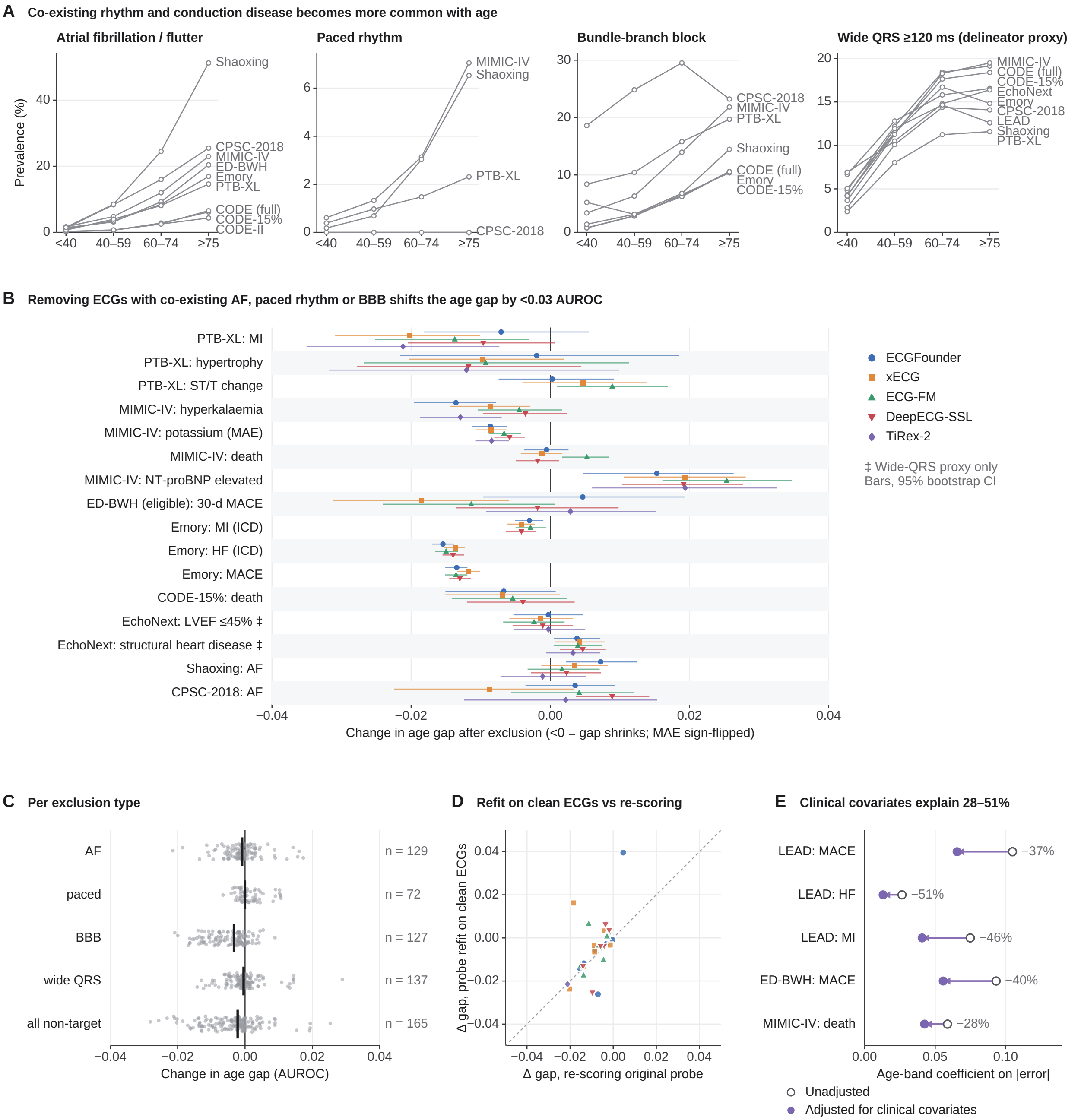

**Figure S21: Ageing versus co-existing disease.** (A) Prevalence of AF, paced rhythm, bundle-branch block and wide QRS by age band. (B) Change in age gap after removing ECGs with co-existing AF, paced rhythm or bundle-branch block (95% paired bootstrap CI). (C) Change by exclusion type. (D) Refitting on the remaining ECGs against re-scoring. (E) Age-band coefficient on per-ECG absolute error with and without clinical covariates. This analysis re-scored stored out-of-fold predictions of the five backbones (largest change 0.028 AUROC in the 149 endpoint–backbone combinations passing the small-n rule). Panel B also shows AF in Shaoxing and CPSC2018, which fail the rule and are not included in the counts; LEAD endpoints are not part of this analysis.

[S3] McInnes L, Healy J, Melville J. UMAP: Uniform Manifold Approximation and Projection for Dimension Reduction; 2018. arXiv preprint; arXiv:1802.03426.

[S4] Rousseeuw PJ. Silhouettes: A graphical aid to the interpretation and validation of cluster analysis. Journal of Computational and Applied Mathematics. 1987;20:53-65.

[S5] Goswami M, Szafer K, Choudhry A, Cai Y, Li S, Dubrawski A. MOMENT: A Family of Open Time-series Foundation Models. In: International Conference on Machine Learning; 2024. arXiv:2402.03885.

[S6] Ke G, Meng Q, Finley T, Wang T, Chen W, Ma W, et al. LightGBM: a highly efficient gradient boosting decision tree. In: Neural Information Processing Systems; 2017. p. 3146-54.

[S7] Smilkov D, Thorat N, Kim B, Viégas F, Wattenberg M. SmoothGrad: removing noise by adding noise; 2017. arXiv preprint; arXiv:1706.03825.

[S8] Li J, Aguirre AD, Junior VM, Jin J, Liu C, Zhong L, et al. An electrocardiogram foundation model built on over 10 million recordings with external evaluation across multiple domains. NEJM AI. 2025;2(7).

[S9] Lunelli R, Nicolson A, Pröll S, Reinstadler S, Bauer A, Dlaska C. BenchECG and xECG: a benchmark and baseline for ECG foundation models. npj Digital Medicine. 2026.

[S10] McKeen K, Oliva L, Masood S, Toma A, Rubin B, Wang B. ECG-FM: an open electrocardiogram foundation model. JAMIA Open. 2025;8(5):ooaf122.

[S11] Nolin-Lapalme A, Sowa A, Delfrate J, Tastet O, Corbin D, Kulbay M, et al. Foundation models for electrocardiogram interpretation: clinical implications. European Heart Journal. 2026;47(18):2174-86.

[S12] Auer A, Klotz D, Bock S, Hochreiter S. Pre-trained Forecasting Models: Strong Zero-Shot Feature Extractors for Time Series Classification; 2025. arXiv preprint, NX-AI (TiRex-2 feature extractor); arXiv:2510.26777.

[S13] Koscova Z, Li Q, Robichaux C, Moura VF, Ghanta M, Gupta A, et al. The Harvard-Emory ECG Database. Scientific Data. 2026;13(1):516.

[S14] Gow B, Pollard T, Nathanson LA, Johnson A, Moody B, Fernandes C, et al. MIMIC-IV-ECG: Diagnostic Electrocardiogram Matched Subset. PhysioNet. 2023 Sep. Version 1.0. Available from: https://doi.org/10.13026/4nqg-sb35.

[S15] Pollard T, Moody BE, Lehman LwH, Gow BJ, Fernandes C, Xie C, et al. PhysioNet as a global platform for biomedical research. Nature Health. 2026 Aug;1(8):792-5. Available from: https://doi.org/10.1038/s44360-026-00096-z.

[S16] Poterucha TJ, Jing L, Ricart RP, Adjei-Mosi M, Finer J, Hartzel D, et al. Detecting structural heart disease from electrocardiograms using AI. Nature. 2025;644(8075):221-30. Dataset described in Poterucha TJ, Jing L, Ricart RP, et al. Detecting structural heart disease from electrocardiograms using AI. Nature 2025; 644: 221–30.

[S17] Ribeiro AH, Ribeiro MH, Paixão G, Oliveira D, Gomes PR, Canazart JA, et al. Automatic diagnosis of the 12-lead ECG using a deep neural network. Nature Communications. 2020;11(1):1760.

[S18] Ribeiro AH, Paixão G, Lima EM, Ribeiro MH, Filho MMP, Gomes PR, et al. CODE-15%: a large scale annotated dataset of 12-lead ECGs (version 1.0.0). Zenodo. 2021.

[S19] Wagner P, Strodthoff N, Bousseljot RD, Kreiseler D, Lunze FI, Samek W, et al. PTB-XL, a large publicly available electrocardiography dataset. Scientific Data. 2020;7(1):154.

[S20] Zheng J, Zhang J, Danioko S, Yao H, Guo H, Rakovski C. A 12-lead electrocardiogram database for arrhythmia research covering more than 10,000 patients. Scientific Data. 2020;7(1):48.

[S21] Liu FF, Liu CY, Zhao LN, Zhang X, Wu X, Xu X, et al. An open access database for evaluating the algorithms of electrocardiogram rhythm and morphology abnormality detection. Journal of Medical Imaging and Health Informatics. 2018;8(7):1368-73.

[S22] Liu H, Chen D, Chen D, Zhang X, Li H, Bian L, et al. A large-scale multi-label 12-lead electrocardiogram database with standardized diagnostic statements. Scientific Data. 2022;9(1):272.

[S23] Breyer-Kohansal R, Hartl S, Burghuber OC, Urban M, Schrott A, Agusti A, et al. The LEAD (Lung, Heart, Social, Body) study: objectives, methodology, and external validity of the population-based cohort study. Journal of Epidemiology. 2019;29(8):315-24.

[S24] Seják M, Sido J, Zahour D. ElectroCardioGuard: preventing patient misidentification in electrocardiogram databases through neural networks. Knowledge-Based Systems. 2023;280:111014. IKEM dataset v1.0.0: Zenodo, doi:10.5281/zenodo.8393007 (CC BY 4.0).

[S25] Collins GS, Moons KGM, Dhiman P, Riley RD, Beam AL, Van Calster B, et al. TRIPOD+AI statement: updated guidance for reporting clinical prediction models that use regression or machine learning methods. BMJ. 2024;385:e078378.

[S26] Beck M, Pöppel K, Spanring M, Auer A, Prudnikova O, Kopp MK, et al. xLSTM: Extended Long Short-Term Memory. In: Neural Information Processing Systems. Neural Information Processing Systems Foundation, Inc. (NeurIPS); 2024. p. 107547-603. arXiv:2405.04517.
